# HeAlth System StrEngThening in four sub_Saharan African countries (ASSET) to achieve high-quality, evidence-informed surgical, maternal and newborn, and primary care: protocol for pre-implementation phase studies

**DOI:** 10.1101/2021.01.06.20248468

**Authors:** Nadine Seward, Charlotte Hanlon, Ahmed Abdulahi, Zulfa Abrams, Atalay Alem, Ricardo Araya, Max Bachmann, Birke Bogale, Nataliya Brima, Dixon Chibanda, Robyn Curran, Justine Davis, Andualem Deneke, Lara Fairall, Souci Frissa, Jennifer Gallagher, Wei Gao, Richard Harding, Muralikrishnan R. Kartha, Andrew Leather, Crick Lund, Maggie Marx, Kennedy Nkhoma, Jamie Murdoch, Inge Petersen, Ruwayda Petrus, Jane Sandall, Andrew Sheenan, Amezene Tadesse, Graham Thornicroft, André van Rensburg, Nick Sevdalis, Ruth Verhey, Chris Willot, Martin Prince

## Abstract

**Objectives:** To achieve universal health coverage, health systems need to be strengthened to support the consistent delivery of high-quality, evidence-informed care at scale. The aim of the National Institute for Health Research (NIHR) Global Research Unit on He**<u>A</u>**lth **<u>S</u>**ystem **<u>S</u>**tr**<u>E</u>**ng**<u>T</u>**hening in Sub-Saharan Africa (ASSET) is to address this need in a four-year programme spanning three healthcare platforms (primary health care for the integrated treatment of chronic conditions in adults, maternal and newborn, surgical care) involving eight work packages. This paper describes the pre-implementation phase research protocols that assess: (1) barriers to accessing care; (2) health system bottlenecks in care process and pathways; (3) quality of care, and; (4) people centredness. Findings from this research are used to engage stakeholders and to inform the selection of a set of health system strengthening interventions (HSSIs) and subsequent methodology for evaluation.

**Settings:** Publicly funded health systems in rural and urban areas in Ethiopia, Sierra Leone, South Africa, and Zimbabwe.

**Population:** Stakeholders including patients and their caregivers, community representatives, clinicians, managers, administrators, and policymakers.

**Study methodologies and delivery:** In each work package, we apply a mixed-methods approach, including: literature reviews; situation analyses; cohort studies; cross-sectional surveys; ethnographic observations; semi-structured interviews, and; focus group discussions. At the end of the pre-implementation phase, findings are fed back to stakeholders in participatory theory of change workshops that are used to select/adapt an initial set of contextually relevant HSSIs. To ensure a theory-informed approach across ASSET, implementation science determinant frameworks are also applied, to help identify any additional contextual barriers and enablers and complementary HSSIs. Outputs from these activities are used to finalise underlying assumptions, potential unintended consequences, process indicators and implementation and clinical outcomes.

**Conclusions:** ASSET places a strong emphasis of the pre-implementation phase of the programme in order to provide an in-depth and systematic diagnosis of the existing heath system functioning, needs for strengthening and active stakeholder engagement. This approach will inform the design and evaluation of the HSSIs to increase effectiveness across work packages and contexts, to better understand what works, for whom, and how.

**Strengths and limitations of this study:**

- The National Health Institute of Research (NIHR) Global Research Unit on Health System Strengthening in sub-Saharan Africa (ASSET) is a four-year programme (2017-2021) that is closely aligned with the SDG goal of UHC, and the recommendations of the Lancet Commission for High Quality Health Systems.
- The aim of ASSET is to develop and evaluate effective and sustainable HSSIs, promoting consistent delivery of high-quality, people-centred care.
- The ASSET programme is being conducted in two phases including the diagnostic pre-implementation and piloting/rolling implementation phase.
- The purpose of this paper is to describe the methodology for the pre-implementation phase, which has the core aim of mapping comprehensive care pathways of a patient’s journey though the health system including the community, different providers), and health facilities, documenting what care is provided at what level of the health system and the associated health system bottlenecks.
- At the end of the pre-implementation phase of ASSET, it is hoped the common approach taken across different countries, care platforms and health conditions will facilitate cross platform learning and understanding of how differences in health systems and broader contextual influences shaped the development of the interventions.
- The overarching expectation is that by using an in-depth participatory process to engage with the stakeholders and map care pathways to and through the health system, we develop a HSS programme that can be implemented at scale that meets the needs and priorities of the local community.

## Introduction

Substantial gains in survival have been made in low -and middle-income countries (LMICs), mainly through vertical programmes targeting infectious diseases including malaria, HIV and tuberculosis (TB), maternal, newborn and child conditions, and vaccine-preventable deaths.(1) However, siloed approaches to care are inefficient and undermine the aspiration of integrated people-centred care.(2) Furthermore, the epidemiological transition to greater disease burden from chronic and often multi-morbid disorders, driven by increased life expectancy and globalisation of behaviours associated with unhealthy lifestyles, brings new challenges to the provision of high-quality care.(1) The coronavirus disease 2019 (COVID-19) pandemic has intensified these issues, resulting in health systems being unable to cope with the increased use in services.(3)

The accelerated demand and recently exposed fragility of health systems, presents challenges to the United Nations Sustainable Development Goals (SDGs) launched in 2015 which includes a call for Universal Health Coverage (UHC), implicit within which is access to quality healthcare with financial risk protection.(4-6) The 2017 Lancet Commission for High-Quality Health Systems, emphasises that resilient, high-quality health systems are required to meet the escalating demands and prevent against health crisis such as Ebola epidemic in west Africa between 2014-2016.(7) The Commission describes high-quality health systems as “consistently delivering care that improves or maintains health, being valued and trusted by all people and responding to changing population needs”. Estimates are provided that suggest such high-quality health systems can save 8.6 million lives a year in LMICs. Of these potentially avertable deaths, estimates suggest that five million lives can be saved a year with high-quality care and 3.6 million lives can be saved by improving access to care.(8) The Commission urges that health systems strengthening to ensure high-quality healthcare should be a core component of UHC.(7)

Strengthening health systems is not only about saving lives, but also adapting a more comprehensive and systems view of multiple co-morbid conditions including chronic communicable and non-communicable diseases (NCDs) with an agenda of improving outcomes throughout the life course. Surgical care is an integral component of any health system that is required to treat many of these chronic conditions.(9) Scaling up basic surgical care in LMICs to treat acute and chronic conditions can help manage longer-term disability that also has the potential to prevent an estimated 1.4 million deaths per year.(10) An often neglected area in health systems is palliative care, that UHC identifies as an essential health service for patients and families facing progressive disease.(11) There is also growing burden of oral disease in low income countries (12, 13), and a very weak dental system in the region (14) Strengthening health systems to manage the epidemiologic shift to chronic multi-morbid conditions and anticipate shocks such as the Covid-19 pandemic and the Ebola crisis, will need to target multiple components of the health system at different levels, and not just focus on improving avertable deaths.

Unsurprisingly, achieving UHC with high-quality care has been identified as an urgent priority for health systems strengthening (HSS) in LMICs that requires the translation of knowledge (evidence-based care) into policy and routine practice (evidence-informed care).(7, 15) HSS involves comprehensive changes to policies and regulations, organisational structures, and relationships across the health system building blocks that motivate changes in behaviour among providers and patients, allowing more effective use of resources to improve healthcare across the board.(16, 17) Interventions to strengthen health systems, by their very nature, improve health outcomes by providing components that influence several mechanisms, both simultaneously and at various time points and levels of the health system.(16)

Implementation research, which applies a multidisciplinary approach to understand which interventions and implementation strategies work for whom, and how, can be usefully applied to HSS by identifying and addressing barriers and opportunities to the delivery of high-quality quality care and testing potential solutions.(18) Of particular importance is the pre-implementation phase of research that involves careful assessment of context to understand and address barriers to implementation of high-quality evidence-based health care.(19) This approach can be used to inform the development of a set of health system strengthening interventions (HSSIs) that can deliver high-quality evidence-informed care to support the specific needs of a community and health system.(20)

### The ASSET research programme

The National Health Institute of Research (NIHR) Global Research Unit on Health System Strengthening in sub-Saharan Africa (ASSET) is a four-year programme (2017-2021) that is closely aligned with the SDG goal of UHC, and the recommendations of the Lancet Commission for High Quality Health Systems. The aim of ASSET is to develop and evaluate effective and sustainable HSSIs, promoting consistent delivery of high-quality, people-centred care.(21) ASSET is working on three healthcare platforms: (1) primary care for the integrated treatment of chronic conditions in adults; (2) maternal and newborn care; and (3) surgical and dental care, across four diverse sub-Saharan African countries: Ethiopia, Sierra Leone, South Africa, and Zimbabwe. Eight work packages use a common approach to develop a set of context-specific HSSIs that address locally relevant and platform-specific challenges, while bringing wider system benefits. Summaries of the care platforms and associated work packages can be found in Table 1.

**Table 1:**
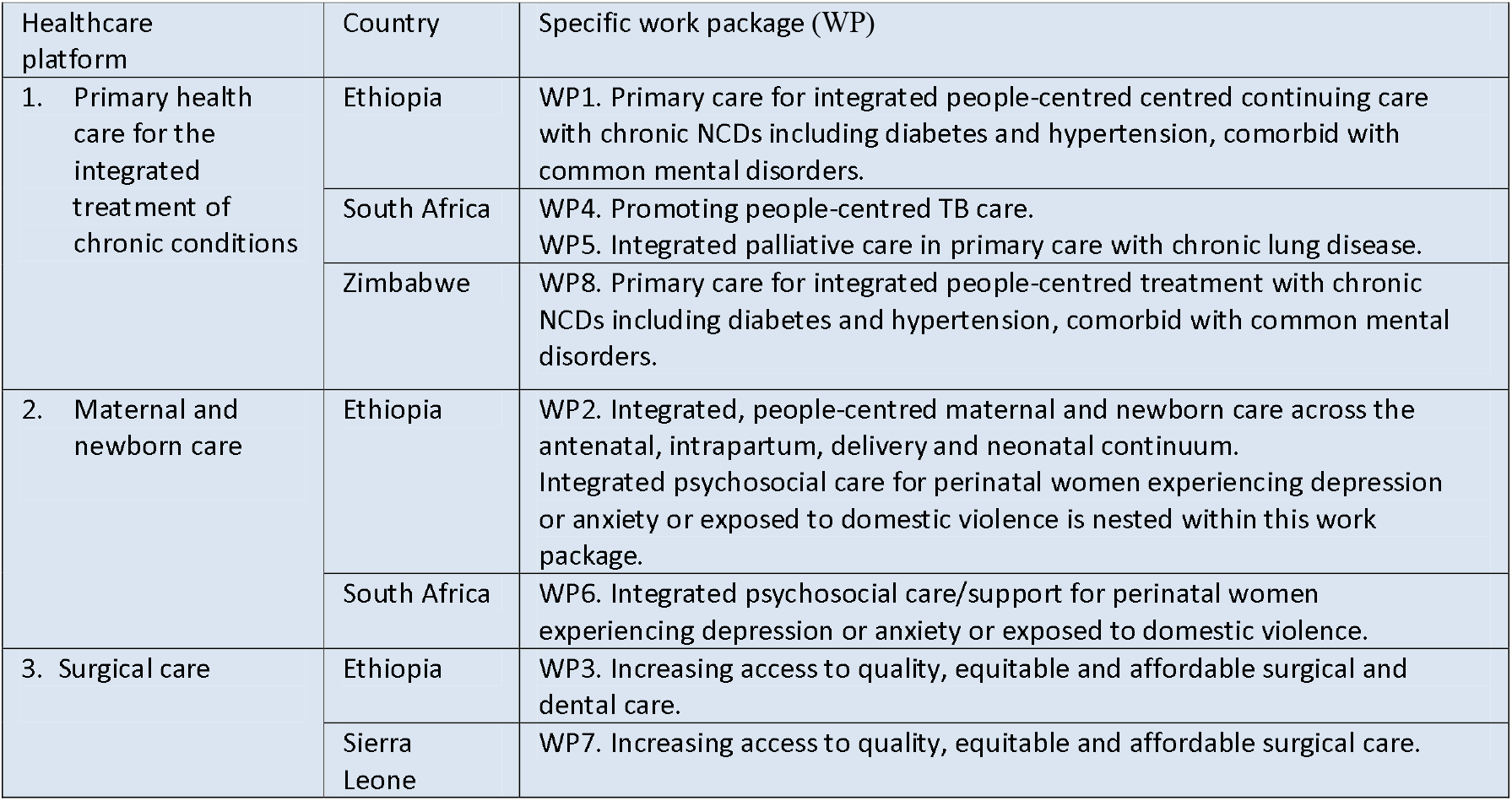
Description of the ASSET work packages for the different healthcare platforms.

The ASSET programme is being conducted in two phases including the diagnostic pre-implementation and piloting/rolling implementation phase. The purpose of this paper is to describe the methodology for the pre-implementation phase, which has the core aim of mapping comprehensive care pathways of a patient’s journey though the health system including the community, different providers (e.g. private sector and non-governmental organisations), and health facilities, documenting what care is provided at what level of the health system and the associated health system bottlenecks. The emphasis on mapping care pathways helps to anchor the work within a people-centred framework and also engage stakeholders in the co-production of HSSI. Participatory Theory of Change (ToC) workshops involving all relevant stakeholders are used throughout ASSET to develop and refine the programme theories for the selected HSSIs.(22) ToC methodology is another important component of ASSET that is a participatory approach involving stakeholders that allows the articulation of the ‘theory’ of how a complex interventional programme will work in reality, describing the necessary interventions to bring about the change, as well as the assumptions inherent to the programme and importantly the context of implementation.(23) After adjusting the initial programme theories, pilot studies using quasi-experimental designs combined with implementation methodologies are used to test the effectiveness of the set of HSSIs for each work package and to identify factors that may influence the implementation of the proposed interventions.(21)

ASSET requires an extensive pre-implementation phase occurring between March 2018 until March 2021. A combination of different methods are used to effectively account for gaps in the provision of high-quality healthcare. The importance of taking time to engage with stakeholders who are part of the public health system cannot be underestimated, as this helps to ensure their needs and priorities are addressed and a set of HSSIs are selected to address local barriers identified for people in need of care. Another factor shaping the pre-implementation phase, is the lack/absence of routinely available data that is of sufficient quality to provide insight into key issues that need addressing (i.e. disease burden, quality of care).

ASSET is one of the first implementation research programmes for HSS that involves diverse care platforms, across different contexts, that also applies a common implementation science approach to the design and evaluation of HSSIs, thus allowing for comparability across settings and potential generalisability. In this paper we describe the methods being used in the pre-implementation phase of the ASSET programme, to understand the requirements of the health system to deliver high-quality people-centred care and describe how the findings are used to inform the selection and adaptation of contextually relevant HSSIs.

The cross-cutting objectives for the pre-implementation phase (phase 1) of ASSET are:

1. To generate engagement and build relationships with stakeholders from the outset to ensure co-production and ownership of HSS that will survive the programme and help to generate both capacity building and sustainability.
2. To apply a mixed-methods approach (qualitative and quantitative) to evaluate the following for each of the work packages:
  I. Barriers to accessing care;
  II. Bottlenecks (critical shortage of a particular resource that results in care being blocked) in the care processes and pathways and associated outcomes;
  III. Quality and care (detection, diagnosis and treatment); and
  IV. People-centred care and its determinants.
3. For each work package, the outputs from the different studies are used to inform the following:
  I. ToC workshops to develop a programme theory Illustrating how and why the package of HSSIs are hypothesised to deliver valued outcomes in practice;(24)
  II. A set of HSSIs to overcome the contextual determinants of problems identified for the different care pathways at the micro, meso and macro levels; and
  III. Selection and development of process, clinical and implementation outcome measures for the different HSSIs.

## Methods

### Study location

The eight work packages cover a range of demographic populations (i.e. with respect to gender, age, socioeconomic status, medical and social needs), located in rural, peri-urban and urban geographical settings.(21) Table 2 describes the different study sites and types of publicly-funded health facilities for each work package.

**Table 2.** Study locations and relevant health facilities of each Work Package (WP)

| Work package | Location | Public Health facilities |
| --- | --- | --- |
| **Person-centred primary healthcare for chronic conditions** |  |  |
| Ethiopia (WP1) | Three districts of the Gurage Zone, Southern Nations, Nationalities and Peoples' Region of Ethiopia. | One general hospital - staffing includes key specialists, a surgeon, an obstetrician/gynaecologist, a radiologist in addition to the staff available in primary hospitals<br>one Primary hospital – staffed by non-specialised doctors, integrated emergency surgical officers, health officers, midwives and nurses.<br>18 health centres staffed by health officers, midwives and nurses.<br>125 health posts staffed by community-based health extension workers |
| South Africa (WP4) | Amajuba District Municipality in the province of KwaZulu-Natal. Predominately African (isiZulu) population. | Four primary healthcare facilities, staffed by nurses, and one public sector hospital, staffed by doctors and nurses. Tuberculosis treatment is limited to the public sector in South Africa and is provided free at point-of-care. |
| South Africa (WP5) | Cape Town Metropolitan area. | Three primary care district hospitals staffed by doctors, nurses, nursing assistants, a social worker, HIV counsellors, pharmacists and pharmacy assistants, physiotherapists, radiographers, dieticians, dentist and part-time occupational therapists.<br>Two 24-hour community health centres with emergency units staffed by doctors, nurses, nursing assistants, a social worker, HIV counsellors, pharmacists, pharmacy assistants, a physiotherapist, part-time or full-time dietician, a part time occupational therapist, health promoters, dentists and dental assistants at one only.<br>Three eight-hour community health centres with emergency rooms doctors, nurses, nursing assistants, a part-time social worker, HIV counsellors, pharmacists, pharmacy assistants, health promoters in two, part time dieticians for all, physiotherapist in one, dentist, dental assistant and oral hygienist in one, part time rehab nurse in one and part time psychiatric nurse in one. |
| Zimbabwe (WP8) | The cities of Harare, Chitungwiza and Gweru. | Nine poly clinics (i.e. primary health care clinics that offered more services in terms of maternal, newborn and childcare, and HIV). The nurse in charge oversees all activities and leads the support staff consisting of community nurses, mental health nurses, midwives, HIV counselors, lay health workers, nurse aids, and pharmacy technicians.<br>Medical doctors are not permanently present and will hold clinics on specific days in the poly clinics which also influence the composition of the clinic user population on these particular clinic days (for example HIV clinic day). |
| **Maternal and newborn care** |  |  |
| Ethiopia (WP2) | Three districts of the Gurage Zone, Southern Nations, Nationalities and Peoples' Region of Ethiopia. | One general hospital - staffing includes key specialists, a surgeon, an obstetrician/gynaecologist, a radiologist in addition to the staff available in primary hospitals<br>one Primary hospital – staffed by non-specialised doctors, integrated emergency surgical officers, health officers, midwives and nurses.<br>18 health centres staffed by health officers, midwives and nurses.<br>125 health posts staffed by community-based health extension workers |
| South Africa (WP6) | Cape Town Metropolitan area. | Four Midwife Obstetric Units (MOUs) in the Cape Town Metropolitan area staffed by antenatal care nurses, midwives, health promoters and breast feeding counsellors. |
| **Surgical care** |  |  |
| Ethiopia (WP3) | Gurage and Silte Zones, Southern Nations, Nationalities, and Peoples' Region. | The Ethiopian Health Alliance for Quality cluster (7 hospitals) co-led by a general hospital in the study site. |
| Sierra Leone (WP7) | Western Area of Sierra Leone including Freetown and surrounding districts. | One tertiary level government facility providing surgical care in the Western Area (Freetown and surrounding districts) that is staffed by consultant surgeons and Consultant anaesthetists and the full range of staff that you would expect at a large tertiary site.<br>Two district hospitals - with limited surgical and anaesthetic staff – a medical officer would do the operations (general doctor with no specialist training)<br>Six Peripheral Primary Health Units staffed by nurses only. |

### Participants

The participants in the pre-implementation phase of ASSET are a combination of stakeholders, including patients and their caregivers, clinicians, managers, administrators and policy makers. ASSET also engages with important community/multi sectorial stakeholders such as non-governmental organisations and private healthcare providers. Broad, in-depth and sustained stakeholder involvement from the outset is a core component of ASSET and fundamental to ensuring that the needs of the health system and communities they serve are appropriately addressed with contextually appropriate HSSIs.

### Research studies within the pre-implementation phase

Across the work packages, a combination of qualitative and quantitative methods and ToC workshops are used to identify a set of HSSIs to overcome the contextual barriers at the micro, meso and macro levels identified for the different care pathways. The following research methods will be applied: literature/scoping reviews; situation analyses; cross-sectional surveys involving patients identified in healthcare facilities; follow-up surveys in the communities involving patients who received surgical interventions in participating healthcare facilities; cross-sectional community surveys to identify unmet need for surgical treatment; ethnographic observations of provision of healthcare; semi-structured interviews and focus group discussions, and ToC workshops. Triangulation of the findings is used to help substantiate and add validity to the overall findings, illuminating where important differences exist, for example, differences between what we observe and what is reported in semi-structured interviews, or differences between perspectives among stakeholder groups. The protocols for the pre-implementation of ASSET for each of the work packages can be found in Appendix 1. Table 3 describes the objectives of for the different studies that are applied in the pre-implementation phase of ASSET.

**Table 3.** Rational/Objectives of pre-implementation phase studies.

| **Method**<br>**(work**<br>**package(s))** | **Objectives**<br>**(not all objectives apply to each work package)** |
| --- | --- |
| Literature review (1-8) | Collate evidence for cross-cutting issues relevant to all work packages (i.e. people-centred care, skills required for HSS, integrated primary care) to better address ASSET's main objectives.<br>Identify evidence-base for effective HSSs that have potential relevance to the local context. |
| Situation analysis (1-8) | Appraise the national and local area level health systems context including demographic characteristics, epidemiology, policies and plans, guidelines, patient's care pathways based on both local and national guidelines, stakeholders and community resources.<br>Examine routinely collected data to understand the numbers of treated patients and the outcomes of care.<br>Use routinely available data to identify the requirements of the intervention to address the gaps in the quality of care for the different recommended care pathways.<br>Evaluate routinely collected data including information available in Healthcare Management Information Systems (HMIS) to understand coverage, completeness, quality of process and clinical outcome indicators. This will provide insight as to the availability of data to capture process and outcome indicators to contribute to the research evaluation of the intervention.<br>Identify requirements of local facility resources and infrastructure in meeting the current disease burden including:<br>Healthcare workers, roles, skills and competencies; availability of equipment, medications and other treatments, and investigations; policies, guidelines and procedures, and organisation of care.<br>Identify structural barriers to delivering evidence-based care that can occur both within a health facility (e.g. waiting times, out-of-pocket costs, availability of services) as well as externally (e.g. distance to facility, transportation, childcare). |
| Cross-sectional surveys of people attending at healthcare facilities (1, 2, 5, 6, 8) | Evaluate the prevalence of morbidity and comorbidity to demonstrate the added value of an integrated care approach to address existing disease burden.<br>Document care processes and outcomes including the proportion of cases that appropriately detected and treated to identify components of the intervention that can be used to improve quality of care.<br>Evaluate patient knowledge and awareness of the condition/s including self-care management, help-seeking behaviour and participation in decisions around their care to identify requirements to support patients to take a more informed and active role in their care. |
| Cohort study of patients identified at tertiary health care facility as having a condition requiring surgical intervention and subsequently followed up in the community (3,7) | Evaluate patient reported outcomes, satisfaction with care and associated determinants to identify requirements of the intervention to address quality of care as experienced and reported by patients (wp3, wp7).<br>Establish prevalence of peri- and post-operative complications (wp3).<br>Evaluate level of satisfaction with inpatient surgical care (wp3, wp7).<br>Assess disability relating to surgical care (wp3, wp7). |
|  | Identify the requirements of the intervention to address the affordability of surgical care by evaluating direct and indirect out-of-pocket costs of care for patients (wp3, wp7).<br>Examine the burden and impact of oral/ dental conditions, and their determinants as reported by the community and oral examination (wp3).<br>Identify the unmet dental treatment needs and investigate access to care in the community (wp3).<br>Explore the determinants of the oral/ dental conditions and barriers to dental care attendance (wp3). |
| Population based community survey to identify unmet need for surgical (3, 7) and dental care (3) | Evaluate the prevalence of surgical conditions (defined as those in need of assessment and/ or care), and unmet needs for surgical intervention as reported by patients (wp3).<br>Use verbal autopsy data to determine what proportion of deaths are potentially avertable with surgical intervention (wp3).<br>Determine the burden of life-limiting surgical conditions in the community (wp3).<br>Evaluate for the difference in level of impairment, pain, overall functioning and days out of work between those receiving and not receiving surgical intervention (wp3).<br>Identify nodes for improving timely access to surgical care by describing help-seeking behaviours and pathways to and through care (wp3).<br>Compare costs for those people with surgical conditions who did or did not receive surgical intervention. Findings can be used to gain insight into how addressing structural barriers to access might impact on residual burden related to not accessing interventions (wp3).<br>Evaluate the willingness to pay for surgical care (wp3, wp7) to inform policy development for financing, particularly equitable co-payment structures that mitigate risk of catastrophic or impoverishing health expenditure.<br>Examine the burden and impact of oral/ dental conditions, and their determinants as reported by the community and oral examination (wp3) and the level of unmet need for care (wp3).<br>Explore patterns of access to dental care and barriers to care of people in the community (wp3). |
| Documentary analysis involving review/analysis of local guidelines, policies, Health Management Information Systems (HMIS), and clinical records and case notes (1, 2, 3, 4, 7, 8) | Compare care processes and pathways, and their variation among defined patient groups, to the ideal standards described in locally applicable guidelines/ standards or what is known based on evidence-based care. Findings will be used to identify components of the intervention to improve the quality and continuity of care by enhancing adherence to evidence-based/ guideline-based care.<br>Identify necessary improvements to the quality and frequency of documentation for routinely available information (i.e. hard copy data, HMIS) by describing current clinical record keeping practices including quality of clinical records and compliance with current guidelines and record management.<br>Assess any routine procedures for aggregating clinical information into HMIS data (process and outcome indicators).<br>The above outputs will ensure information for relevant process and clinical and |
|  | implementation outcome indicators are captured. In turn, this information will be used to evaluate the impact of the intervention (context, quality of care and its determinants). |
| Ethnographic observations of health care practices<br>(1, 2, 3, 4, 6) | Document the ecology of care (i.e. the physical and social/ interpersonal environment) to identify behavioural change opportunities. Ecology is a broad construct which encompasses aspects such as respect for privacy, interactions between healthcare professionals (different cadres and hierarchies), and between staff and patients.<br>Identify factors that influence provision of person-centred, compassionate and respectful care.<br>Map out care pathways to understand bottlenecks.<br>Identify training needs and other resources required to ensure quality of care, including adherence to evidence-based guidelines and patient safety. |
| Semi-structured interviews and focus group discussions with patients and/or healthcare workers<br>(1-8) | Identify barriers and enablers to correctly detect and treat relevant conditions to inform the selection of HSSIs.<br>Explore the concept of person-centred care and identify barriers and enablers to providing both person-centred care and treating certain conditions associated with stigma including violence against women and TB.<br>Understand perceptions of staff and patients regarding quality of care to inform the selection of HSSIs.<br>Understand how care pathways that were characterised in the pre-implementation phase (i.e. review of clinical documentation) are working in practice.<br>Identify potential issues around implementation of HSSIs including barriers associated with: acceptability and feasibility of proposed intervention; knowledge and beliefs about implementation process; implementation readiness.<br>Investigate patient's willingness to pay for care as well as staff's acceptability of this measure.<br>Explore potential health system strengthening interventions that are acceptable to all stakeholders and based on findings from other studies in the pre-implementation phase of ASSET. |
| Participatory Theory of Change workshop with stakeholders<br>(1-8) | Identify components of possible HSSIs that are acceptable and feasible, necessary and sufficient to effect change towards achievement of the long-term goal.<br>Determine appropriate sequential pathways for the above interventions to achieve valued goals, and their interactions, with a logical sequence of causality between the different components. |
|  | Identify relevant process indicators and outcomes and associated contextual barriers and enablers, that in turn creates a draft framework for evaluation of the HSSIs.<br>Enable stakeholders to come to a common understanding of what the interventions will achieve.<br>Foster buy-in from all stakeholders with agency for implementation.<br>Identify relevant knowledge gaps that need further attention in pre-implementation phase of ASSET. |

### Studies conducted within the pre-implementation phase of ASSET

#### Literature review (WPs 1-8)

Unpublished literature reviews are used to inform ASSET’s cross-cutting objectives for health systems strengthening including: the role of non-technical skills required for HSS (e.g. clinical communication skills, leadership skills, quality improvement skills), integrated primary health care, mHealth, and person-centred care. Individual work package teams also use literature reviews to establish HSSIs that are most effective to address the relevant public health issues in the local context. As an example, work package four is publishing a scoping review of tuberculosis and mental health disorders and person-centred care. (25) Work package eight is conducting a systematic review of HSSI to improve the quality of surgical and anaesthesia care at the hospital level in sub-Saharan Africa. The type of literature review used is dependent on the existing level of evidence for the issue in question.

#### Situation analyses (WPs 1-8)

According to the World Health Organization (WHO), a situation analysis is not only a collection of facts describing the epidemiology, demography and health system status of the population, but also a comprehensive assessment of the full range of current and potential future health issues and their determinants.(26)

Work packages 1-3, 6 and 7 conduct situation analyses of primary health facilities using an adapted version of the situation analysis tool developed by Programme for Improving Mental Health Care (PRIME).(27) The PRIME situation analysis tool was originally developed to appraise district and sub-district mental health systems and services in LMICs for primary care but has broader applicability to chronic care. The ASSET programme uses the adapted PRIME tool to assess publicly available information such as existing policies and guidelines and data to determine the location and nature of the gaps between what services intend to provide compared to what is achieved in practice. The tool is also used to identify some of the critical shortages (staff, skills, knowledge) that contribute to these gaps. Of particular relevance to ASSET is using the tool to assess the availability and quality of HMIS data.(27)

Work Packages 3 and 7 also use the Hospital Assessment Tool, developed by the Ethiopian Federal Ministry of Health in collaboration with the WHO and Programme in Global Surgery and Social Change, to assess secondary and tertiary care facilities for surgical care.(28) (29)

### Cross-sectional patient surveys in health facilities (WPs 1, 2, 5, 6, 8)

To establish the extent of the care gaps and to describe parameters that can influence local HSSI development, work package teams conduct cross-sectional surveys using both validated and bespoke assessment tools, of patients at primary and secondary health facilities. Patients presenting at the different health facilities are recruited consecutively when they attend for care. Following consultation, research measures are administered, including clinical measures of selected chronic conditions. The clinical notes are also reviewed to identify diagnoses and management plans which are compared to the research clinical measures. Patients identified as having a condition of interest, are asked additional questions including about their awareness about self-management and their involvement in decision-making and care planning.

Although in many instances similar questionnaires are used to measure the same outcome (i.e. PHQ-9) for different work packages, in some cases work packages use questionnaires with specific relevance to their local context. The assessment tools are described in Table 4.

**Table 4.**
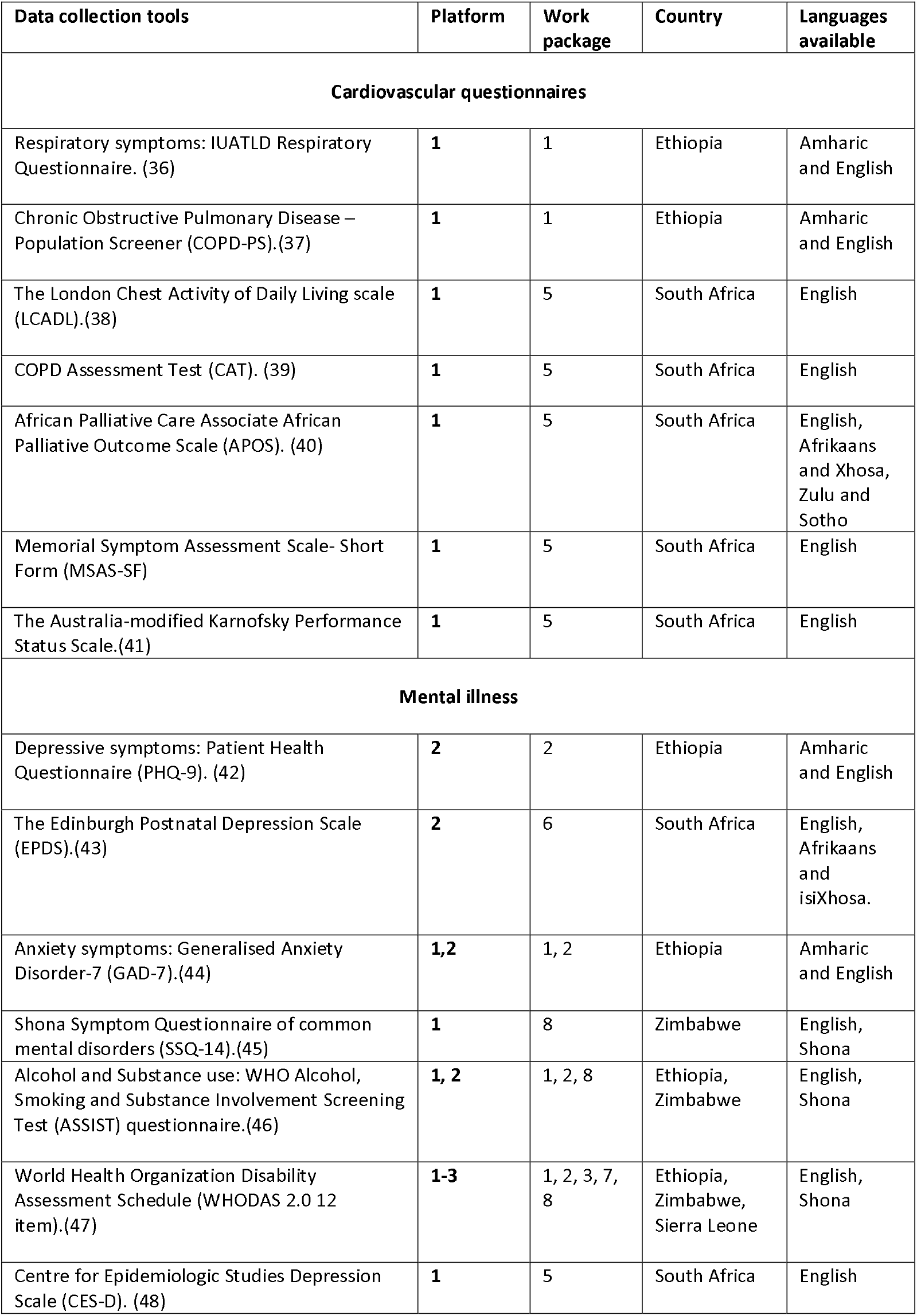
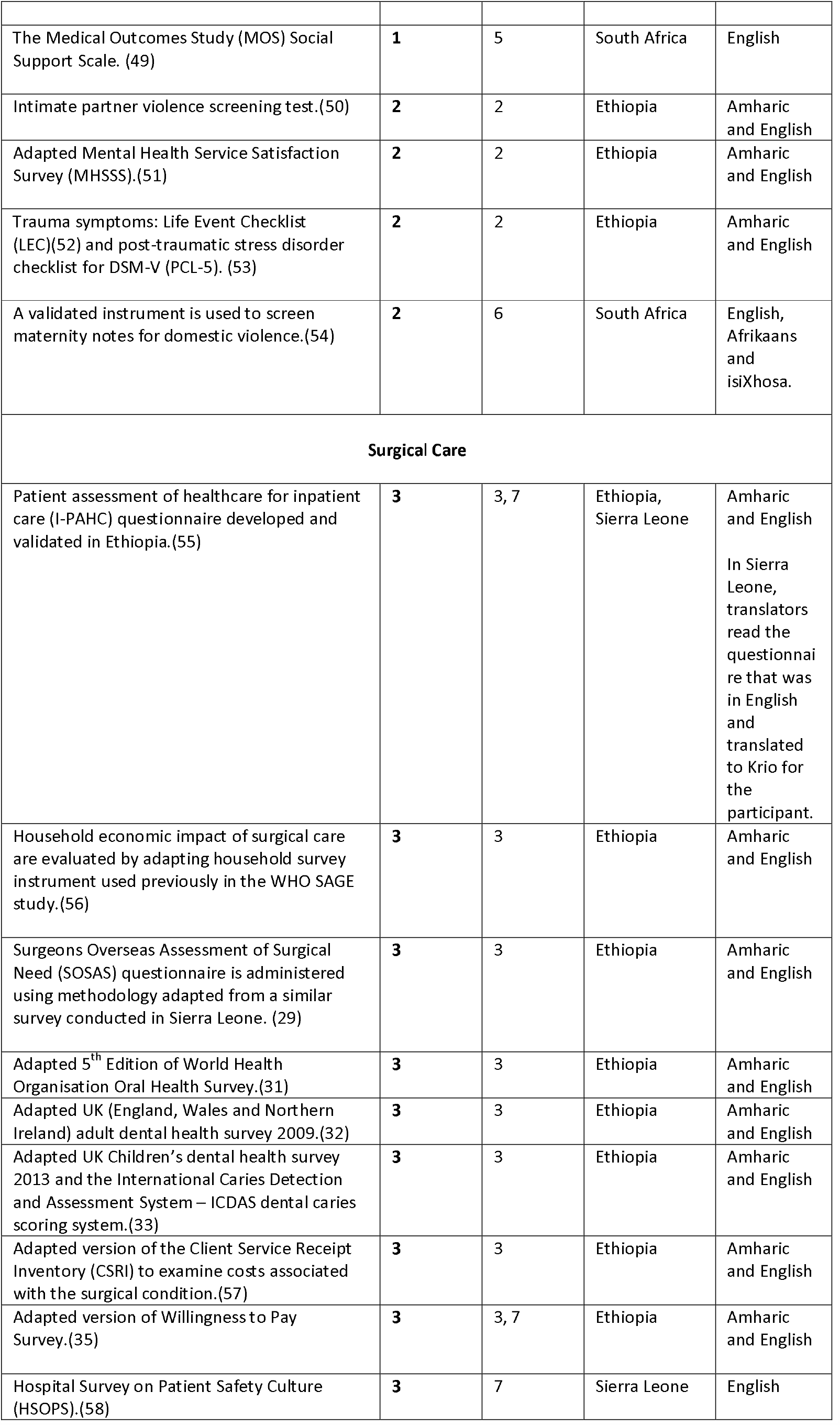
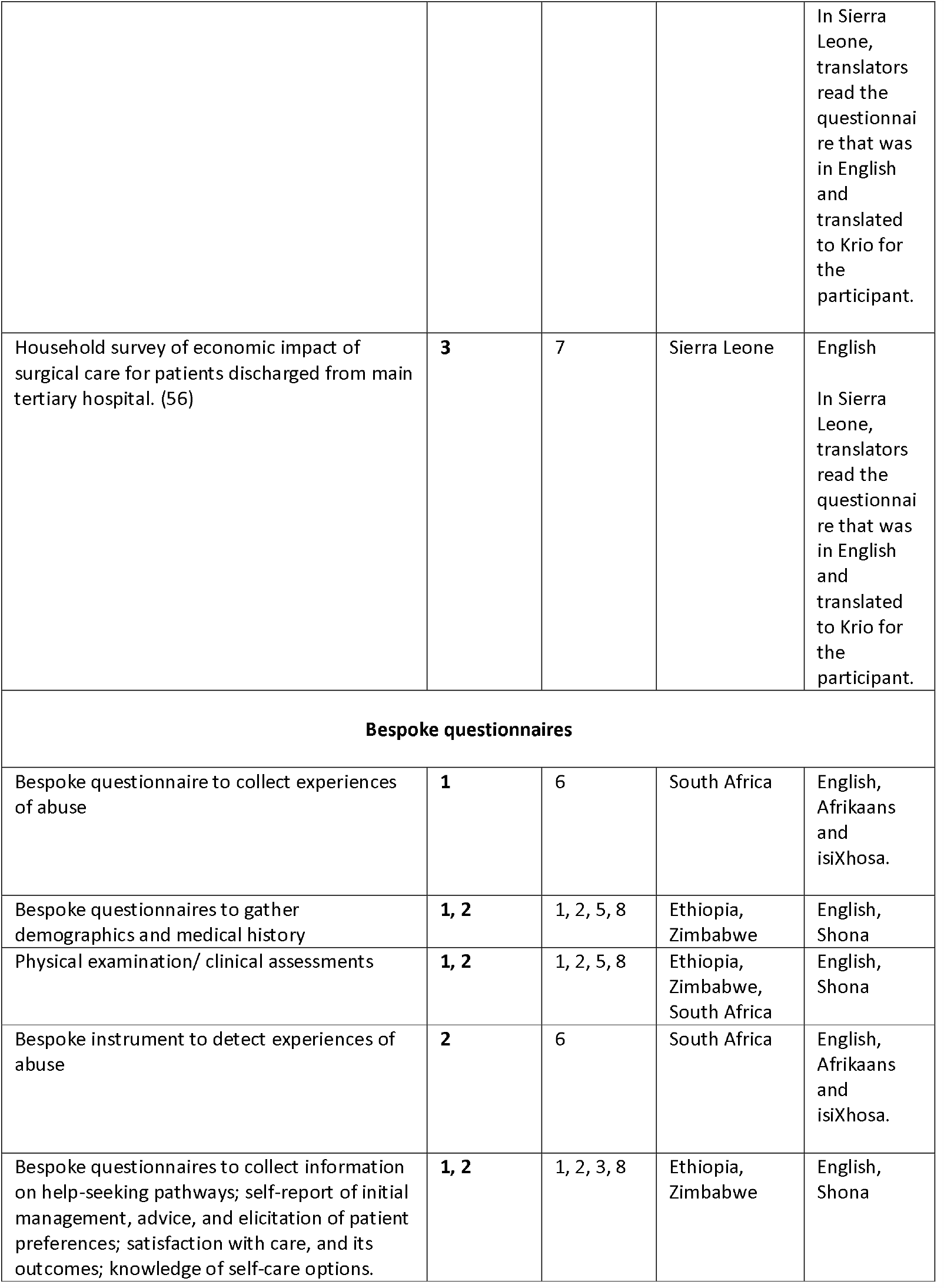
Data collection tools and instruments used in quantitative patient surveys.

| Data collection tools | Platform | Work package | Country | Languages available |
| --- | --- | --- | --- | --- |
| **Cardiovascular questionnaires** |  |  |  |  |
| Respiratory symptoms: IUATLD Respiratory Questionnaire. (36) | 1 | 1 | Ethiopia | Amharic and English |
| Chronic Obstructive Pulmonary Disease – Population Screener (COPD-PS).(37) | 1 | 1 | Ethiopia | Amharic and English |
| The London Chest Activity of Daily Living scale (LCADL).(38) | 1 | 5 | South Africa | English |
| COPD Assessment Test (CAT). (39) | 1 | 5 | South Africa | English |
| African Palliative Care Associate African Palliative Outcome Scale (APOS). (40) | 1 | 5 | South Africa | English, Afrikaans and Xhosa, Zulu and Sotho |
| Memorial Symptom Assessment Scale- Short Form (MSAS-SF) | 1 | 5 | South Africa | English |
| The Australia-modified Karnofsky Performance Status Scale.(41) | 1 | 5 | South Africa | English |
| **Mental illness** |  |  |  |  |
| Depressive symptoms: Patient Health Questionnaire (PHQ-9). (42) | 2 | 2 | Ethiopia | Amharic and English |
| The Edinburgh Postnatal Depression Scale (EPDS).(43) | 2 | 6 | South Africa | English, Afrikaans and isiXhosa. |
| Anxiety symptoms: Generalised Anxiety Disorder-7 (GAD-7).(44) | 1,2 | 1, 2 | Ethiopia | Amharic and English |
| Shona Symptom Questionnaire of common mental disorders (SSQ-14).(45) | 1 | 8 | Zimbabwe | English, Shona |
| Alcohol and Substance use: WHO Alcohol, Smoking and Substance Involvement Screening Test (ASSIST) questionnaire.(46) | 1, 2 | 1, 2, 8 | Ethiopia, Zimbabwe | English, Shona |
| World Health Organization Disability Assessment Schedule (WHODAS 2.0 12 item).(47) | 1-3 | 1, 2, 3, 7, 8 | Ethiopia, Zimbabwe, Sierra Leone | English, Shona |
| Centre for Epidemiologic Studies Depression Scale (CES-D). (48) | 1 | 5 | South Africa | English |
| The Medical Outcomes Study (MOS) Social Support Scale. (49) | 1 | 5 | South Africa | English |
| Intimate partner violence screening test.(50) | 2 | 2 | Ethiopia | Amharic and English |
| Adapted Mental Health Service Satisfaction Survey (MHSSS).(51) | 2 | 2 | Ethiopia | Amharic and English |
| Trauma symptoms: Life Event Checklist (LEC)(52) and post-traumatic stress disorder checklist for DSM-V (PCL-5). (53) | 2 | 2 | Ethiopia | Amharic and English |
| A validated instrument is used to screen maternity notes for domestic violence.(54) | 2 | 6 | South Africa | English, Afrikaans and isiXhosa. |
| **Surgical Care** |  |  |  |  |
| Patient assessment of healthcare for inpatient care (I-PAHC) questionnaire developed and validated in Ethiopia.(55) | 3 | 3, 7 | Ethiopia, Sierra Leone | Amharic and English<br>In Sierra Leone, translators read the questionnaire that was in English and translated to Krio for the participant. |
| Household economic impact of surgical care are evaluated by adapting household survey instrument used previously in the WHO SAGE study.(56) | 3 | 3 | Ethiopia | Amharic and English |
| Surgeons Overseas Assessment of Surgical Need (SOSAS) questionnaire is administered using methodology adapted from a similar survey conducted in Sierra Leone. (29) | 3 | 3 | Ethiopia | Amharic and English |
| Adapted 5<sup>th</sup> Edition of World Health Organisation Oral Health Survey.(31) | 3 | 3 | Ethiopia | Amharic and English |
| Adapted UK (England, Wales and Northern Ireland) adult dental health survey 2009.(32) | 3 | 3 | Ethiopia | Amharic and English |
| Adapted UK Children's dental health survey 2013 and the International Caries Detection and Assessment System – ICDAS dental caries scoring system.(33) | 3 | 3 | Ethiopia | Amharic and English |
| Adapted version of the Client Service Receipt Inventory (CSRI) to examine costs associated with the surgical condition.(57) | 3 | 3 | Ethiopia | Amharic and English |
| Adapted version of Willingness to Pay Survey.(35) | 3 | 3, 7 | Ethiopia | Amharic and English |
| Hospital Survey on Patient Safety Culture (HSOPS).(58) | 3 | 7 | Sierra Leone | English |
|  |  |  |  | In Sierra Leone, translators read the questionnaire that was in English and translated to Krio for the participant. |
| Household survey of economic impact of surgical care for patients discharged from main tertiary hospital. (56) | 3 | 7 | Sierra Leone | English<br>In Sierra Leone, translators read the questionnaire that was in English and translated to Krio for the participant. |
| **Bespoke questionnaires** |  |  |  |  |
| Bespoke questionnaire to collect experiences of abuse | 1 | 6 | South Africa | English, Afrikaans and isiXhosa. |
| Bespoke questionnaires to gather demographics and medical history | 1, 2 | 1, 2, 5, 8 | Ethiopia, Zimbabwe | English, Shona |
| Physical examination/ clinical assessments | 1, 2 | 1, 2, 5, 8 | Ethiopia, Zimbabwe, South Africa | English, Shona |
| Bespoke instrument to detect experiences of abuse | 2 | 6 | South Africa | English, Afrikaans and isiXhosa. |
| Bespoke questionnaires to collect information on help-seeking pathways; self-report of initial management, advice, and elicitation of patient preferences; satisfaction with care, and its outcomes; knowledge of self-care options. | 1, 2 | 1, 2, 3, 8 | Ethiopia, Zimbabwe | English, Shona |

### Cohort study of surgical patients recruited in health care facilities in Ethiopia and Sierra Leone

Work packages that are a part of the surgical care platform, recruit patients who are identified in participating health facilities for follow-up assessment in the community post discharge. Patients are administered questionnaires to identify peri and post-operative infection rates, disability, household economic impact of surgery, and healthcare satisfaction (Table 4). Outcomes are linked to clinical processes for the hospital admission, documented on a daily basis.

### Community-based surveys involving patients in need of dental and surgical care in Ethiopia and surgical care in Sierra Leone

#### Ethiopia

To understand the prevalence and associated burden of unmet need for surgical and/or dental care in Ethiopia, three community-based surveys are conducted by randomly sampling households using the sampling frame of the Butajira Health and Demographic Surveillance Site,(30) then randomly selecting two people within each household to complete the survey.

To identify people in need of surgical and dental conditions, the Surgeons Overseas Assessment of Surgical Need (SOSAS) questionnaire is administered using methodology adapted from a similar survey conducted in Sierra Leone.(29) People identified as having a current/recent surgical problem (in the past two years) are also administered a fully structured questionnaire to investigate help-seeking behaviour, delays/barriers to obtaining surgical care and costs of help-seeking. The dental survey involves a dental examination to assess for dental caries experiences, periodontal diseases and oral mucosal lesions. Additionally a questionnaire survey (linked to the surgical survey) explored their perceived oral health, dental health behaviours including oral hygiene, diet, tobacco and alcohol consumption. The surveys draw on standard instruments (questionnaire and oral examination forms) based on the 5^th^ edition of WHO Oral Health Survey Methods, drawing on: the methodology of the UK (England, Wales and Northern Ireland) adult dental health survey 2009, the UK Children’s dental health survey 2013 and the International Caries Detection and Assessment System – ICDAS dental caries scoring system (31-34), and adapted for use in Ethiopia.

A willingness to pay survey used in Nigeria is adapted for surgical services in Ethiopia. The adapted surveys assess willingness to pay for two common surgical problems, one emergency and one elective (appendicitis and hernia).(35)

#### Sierra Leone

A cohort of patients identified in the hospital who are in need of surgical interventions are followed up in the community and administered questionnaires similar to WP 3. A willingness to pay survey, similar to Ethiopia, is also applied in Sierra Leone.

Table 4 describes the data collection tools. All scales are available from the corresponding author upon request.

#### Documentary analyses (WPs 1-4, 7-8)

A documentary analysis is used to identify the extent and quality of clinical documentation, through review of clinical records, guidelines, policies, and Health Management Information Systems (HMIS). Clinical records are compared to policy guidelines using methods such as process mapping, checklists, observations of patient flow, review of HMIS data, and review of clinical records using proformas. Findings are used to assess for adherence to guideline-based care or evidence-based care. Table 5 describes the relevant guidelines.

**Table 5.** Data sources and data collection instruments for documentary analysis.

| Guidelines | Description | Platform (work package) |
| --- | --- | --- |
| Ethiopian Primary Healthcare Clinical Guideline (59). | The Ethiopian Primary Healthcare Clinical Guidelines have been contextualised from the Practical Approach to Care Kit.(59) The PHCG integrates care for all common presentations to primary health care, based on the best available evidence. PHCG promotes holistic care through integrated treatment algorithms that consider multimorbidity. | 1, 2 (1, 3) |
| Essential drugs list and standard treatment guidelines for Zimbabwe (EDLIZ).(60) | The essential medicines list and standard treatment guidelines covers the most common health conditions in Zimbabwe and is based on the essential medicines concept. It is endorsed by the National Medicine & Therapeutics Policy Advisory Committee (NMTPAC) and was collaboratively created health care workers of all levels of the health care system. It is continuously revised and updated. | 1 (8) |
| Ideal Clinic Policy PACK Guidelines(61) | Ideal Clinic policy promotes integrated clinical services for all patients with a view that patients receive all care by one clinician.<br>PACK guidelines are clinical decision support tools, providing an evidence-based, comprehensive clinical approach to support the treatment of common symptoms, syndromes, diagnoses and conditions. The guides are designed for use in each consultation and starts with screening and a symptom-based approach, guides the diagnosis of common conditions, including priority chronic conditions and facilitates the routine care of the patient with one or several chronic conditions. PACK guides include PACK Child, Adolescent, Adult, Community and Home, thereby covering the life course and supporting all health workers in the primary care team.(62) | 1 (4) |
| Adult Primary Care (APC) Guidelines.(63) | APC guidelines are a comprehensive clinical tool for primary care of adults 18 years or older. The guidelines were developed using approved clinical policies and guidelines issued by the National Department of Health and is intended for use by health care practitioners. APC is being implemented as part of the Integrated Clinical services Management, a key focus within the Ideal Clinic. | 1 (4) |
| National Tuberculosis Management Guidelines.(64) | The National Tuberculosis Management Guidelines provide South African department of Health's guidance for management of TB, guidance on the management of adverse drug events and anti-retroviral initiation for patients co-infected with HIV. | 1 (4) |
| National Infection Prevention Control Guideline for TB, MDR-TB and XDR-TB(65) | The National Infection Prevention Control guidelines for TB, MDR-TB and XDR-TB provide guidance for staff to minimise the risk of TB transmission in health settings. Infection control measures should be established to reduce risk of TB transmission to both the general population and health care personnel. | 1 (4) |
| WHO surgical checklist.(66) | The WHO Surgical Safety Checklist was developed to decrease errors and adverse events and increase teamwork and communication in surgery. The 19-item checklist has demonstrated a significant reduction in both morbidity and mortality and is now used by a majority of surgical providers around the world. | 3 (3, 7) |
| Amajuba Mortality Report.(67) (68) | Summary of TB mortality trends from routine data systems in the Amajuba District Municipality. | 1 (4) |
| Sierra Leone Early Warning Score (SLEWS))(69) | Sierra Leone Early Warning Scoring system (SLEWS) helps to identify deteriorating patients based on a numerical scoring system given to abnormal physiological parameters. | 3 (7) |

### Ethnographic and structured observations of healthcare practices and context (WPs 1-4, 6)

ASSET applies both unstructured and structured ethnographic observations of clinician-patient encounters to complement quantitative methods. Unstructured observations are used extensively to understand quality of care and the broader context of patient interactions. Unstructured ethnographic observations are the best approach for exploring stigmatised conditions like TB, mental illness and domestic violence, that also captures the quality of clinician-patient interactions from a non-clinical perspective. In particular, these methods are useful for looking at issues like respect, compassion, and quality of listening. Communication of healthcare workers amongst themselves and with patients are observed to understand adherence to guideline-based care and the extent to which care is respectful and people-centred. Structured observation of clinician-patient encounters is conducted using observational checklists, including the enhancing assessment of common therapeutic factors (ENACT) rating scale for competence in elements of person-centred care.(70) Checklists are also used to help to determine the extent to which clinicians are adhering to guideline based care.

### Semi-structured interviews and focus group discussions (WPs 1-8)

Qualitative semi-structured interviews and focus-group discussions are used by all work packages to triangulate findings with data from the quantitative surveys and observational data, and to explore perspectives of various stakeholders. Interviews are held with different groups of participants separately, allowing for frank expression of what people experience when probing around sensitive topics. Such an approach provides the human narrative component to complement the quantitative methods and understand trends in data. Table 6 describes objectives, processes and participants for the interviews and group discussions.

**Table 6.**
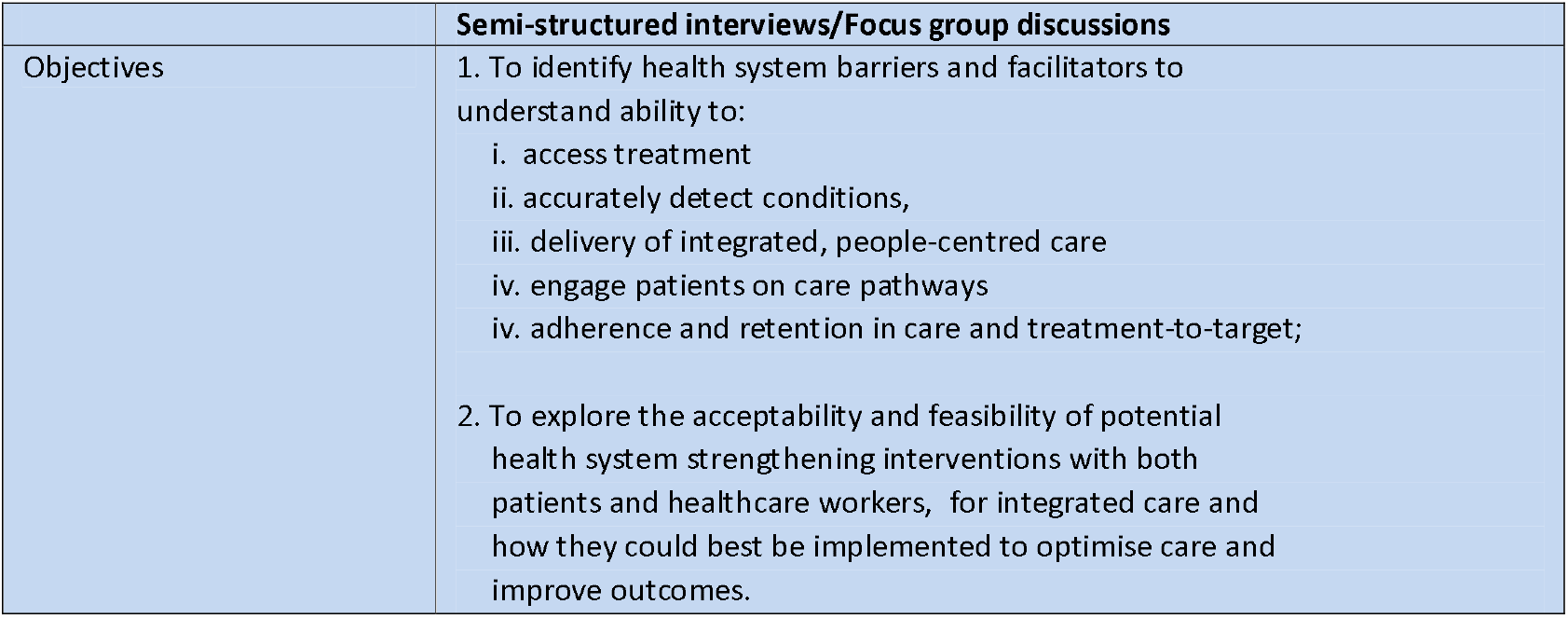
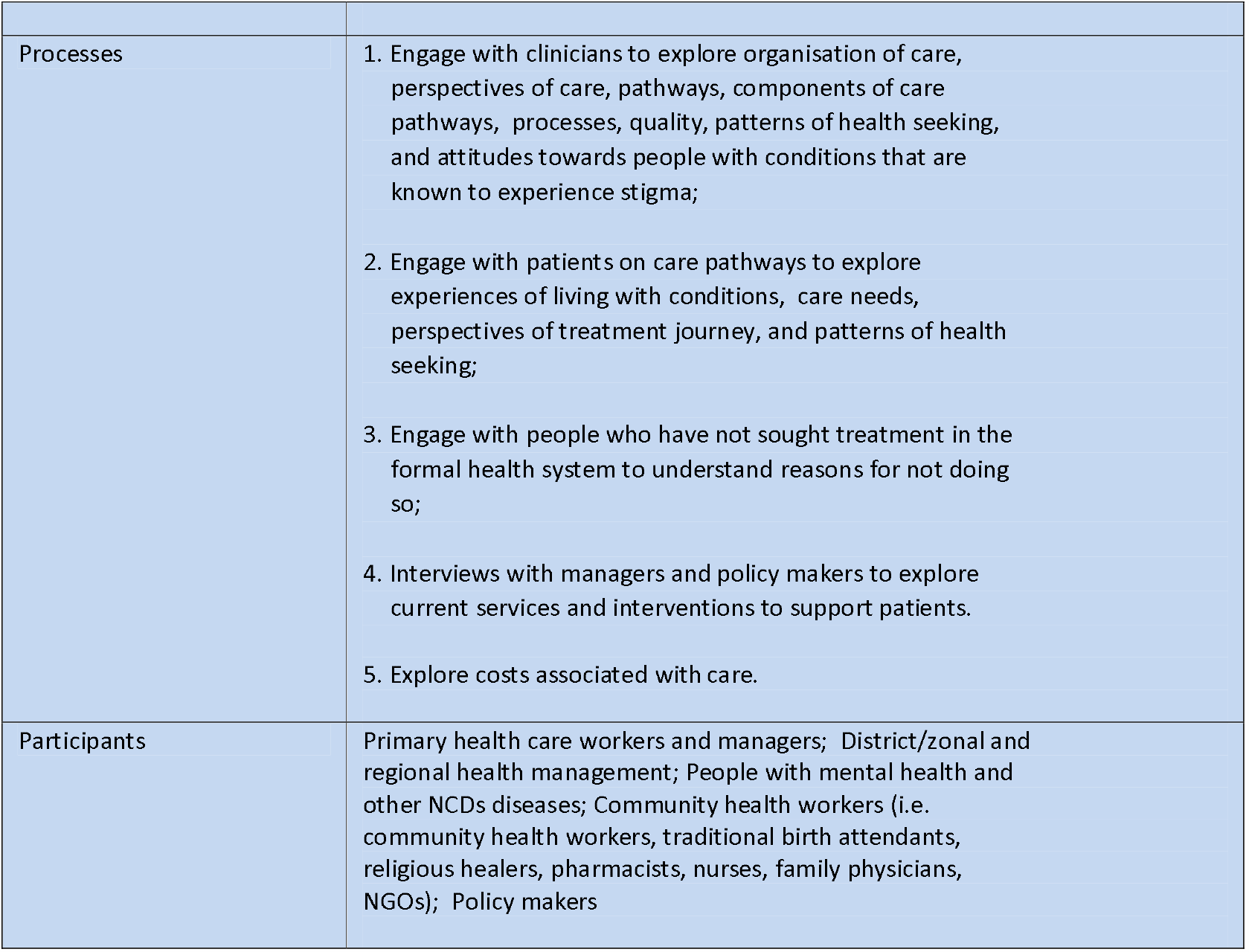
Summary of qualitative data collection methods and samples.

### Methods to inform the piloting and evaluation phases of ASSET

#### Theory of change workshops to develop a programme theory

Findings from the pre-implementation phase of ASSET are shared with the stakeholders during ToC workshops in order to elicit their ideas and priorities for quality improvement. Findings which reflect negatively on quality of care provided within a service are conveyed in such a way so that they can be shared with staff to engage in quality improvement. Confidentiality is key to this process, as is the use of patient narratives (including patient quotes). This approach helps to engage staff in constructive ways conveying how they could work differently. This shifts the quality improvement process from a culture of inspection and punishment to one of true reflection and change.

In the pre-implementation phase of ASSET, work package teams oversee between one and three ToC workshops to develop an initial programme theory (define a programme theory). In subsequent phases of the ASSET programme, work package teams use a series of ToC workshops to adapt and refine the initial programme theory as the implementation process progresses. The result is a final programme theory that articulates pathways to change, intermediate outcomes, clinical and implementation outcomes, and underlying assumptions including contextual barriers and enablers. Each work package team invites different cadres of stakeholders to relevant meetings and workshops. Table 7 describes the ToC workshops used in each work package.

**Table 7.** ToC workshops conducted for each work package in the pre-implementation phase of ASSET.

| Work package | Number of workshops | Timing of workshop and relevant stakeholders involved |
| --- | --- | --- |
| **Integrated primary healthcare care for chronic conditions** |  |  |
| Ethiopia (WP1)<br>Integrated care for persons with NCDs diseases, including common mental disorders | 3 | Workshops held at the beginning of the pre-implementation phase that include stakeholders (Community representatives, health extension workers, primary care clinicians, secondary care clinicians, mental health professionals, and managers).<br>One workshop is held at the end of the pre-implementation phase: national/regional level stakeholders including district, regional and national level administrators and policymakers, service user association representatives, mental health and NCDs disease clinicians and primary care clinicians. |
| Zimbabwe (WP8)<br>Integrated care for persons with NCDs diseases, including common mental disorders | 1 | Held at the end of the diagnostic phase involving community health workers, primary health care nurses, mental health professionals, diabetes association representatives, traditional healers, patients, health service managers and policy makers. |
| South Africa, Cape Town (WP4) Promoting | 3 | ToC with initial findings from the pre-implementation phase that involved the District TB Programme Coordinator, Hospital CEO, clinical and nursing |
| person-centred TB care |  | management, facility managers, and Primary HealthCare (PHC) manager.<br>ToC including reporting of additional research requested at the first workshop that included TB District Manager, Hospital Manager, Facility Managers, PHC manager, clinicians.<br>Co-development of intervention that included Community Health Worker Manager, Operational Managers of facilities, District Director, Nursing Managers, Ward-Based Outreach Team leaders (who supervise teams of community health workers), and PHC Supervisors. |
| South Africa, Cape Town (WP5) Integrated palliative care with chronic obstructive pulmonary disease | 1 | Held at the end of the diagnostic phase involving patients, family members, primary care physicians, palliative care physicians, respiratory physician, one representative from the department of health. |
| **Maternal and newborn care** |  |  |
| Ethiopia (WP2)<br>1. maternal and newborn care across the antenatal, intrapartum, delivery and neonatal continuum; | 2 | ToC at the beginning of the pre-implementation phase that includes: community representatives, health extension workers, primary care clinicians, secondary care clinicians, and managers). Results of this workshop are shared with the surgical ToC, given the overlap in stakeholders.<br>ToC to be held at the end of the pre-implementation phase with national/regional level stakeholders including district, regional and national level administrators and policymakers, service user association representatives, clinicians. |
| 2. Integration of psychosocial care for perinatal women experiencing mental health problems or exposed to domestic violence | 3 | Two ToCs held at the beginning of pre-implementation phase: district-level participants including NGO representative and women with experience of IPV, community health extension workers and primary health care clinicians. At the first workshop there is also an expert group including mental health researchers, mental health clinicians, social workers, a psychologist with experience in adapting/delivering mental health interventions in the study site.<br>Intervention adaptation workshop: perinatal women with experience of depression and primary healthcare workers. |
| South Africa, Cape Town (WP6) Integration of psychosocial care for perinatal women experiencing mental | 1 | End of the pre-implementation phase involving health service managers in the Western Cape Department of Health responsible for PHC, maternal health and mental health in the City of Cape Town and its sub-districts.<br>Key to finalising the programme theory is the continued |
| health problems or exposed to domestic violence |  | engagement through feedback sessions at the health care facilities with stakeholders. |
| **Surgical care** |  |  |
| Ethiopia (WP3) Surgical and dental care | 3 | ToC held at the beginning of the pre-implementation phase includes: community representatives, health extension workers, primary care clinicians, secondary care clinicians, and managers. Results of this workshop are also shared with maternal obstetric care work package.<br>ToC held at end of pre-implementation phase with national/regional level stakeholders including district, regional and national level administrators and policymakers, service user association representatives, clinicians. |
| Sierra Leone (WP7) Surgical care | 2 | The first ToC was held at the end of the baseline assessment of the health system, and attended by patient representatives, senior hospital managers, senior surgeons, junior doctors, anaesthetists, nurses including matrons, Primary Health Unit leads, representatives from Ministry of Health and Sanitation, local Non-Governmental Organisations. The second was held a few months later and attended by senior hospital managers, senior surgeons, junior doctors, anaesthetists, nurses including matrons. |

#### Implementation Science Methods

Implementation science theory-based determinant frameworks provide a systematic approach to the identification and description of contextual factors that are known to influence implementation outcomes, as well as key factors to consider in the design, implementation and evaluation of the HSSIs.(71) Implementation science frameworks are applied to findings from the pre-implementation phase of ASSET to help interpret the findings, identify commonalities and differences across platforms and countries and ultimately help identify how context at the micro, meso and macro levels may influence the implementation of evidence-informed care.

There are numerous frameworks and theories that could potentially be applied within ASSET. A formative approach is taken, in which the implementation science cross-cutting theme within ASSET initially shortlists a few relevant frameworks and subsequently critically present, review and select those most suitable jointly with the ASSET work-packages reported in this paper. This approach allows an informed yet flexible approach to choosing a number of useful and heuristic tools for the purpose of ASSET implementation theories. The corollary of the approach will be to enhance implementation science understanding and capabilities across ASSET work-packages. The implementation science element of the ASSET programme is reported in a separate protocol, due to its breadth and complexity, as the ASSET diagnostic phase gets underway. Ultimately, the implementation science component of ASSET will help determine the following: (1) whether any additional health systems strengthening interventions are required, (2) finalise process indicators and outcomes of interest in the programme theory developed in the ToC workshops, and (3) inform the design of the piloting and evaluation phases in terms of contextual factors that may influence the effectiveness of the HSSI in delivering evidence-based and people-centred care.

### Data analyses

A combination of mixed methods are used to analyse the data collected as part of the pre-implementation phase. Quantitative outcomes of interest are reported as means and proportions, accounting for clustering where appropriate. Regression analysis (logistic, negative binomial) accounting for clustering where appropriate, is used to determine predictors of quality of care (i.e. accurately detecting conditions), satisfaction with care, recovery, and risk factors for the condition in question.

Qualitative analyses will use simple descriptive summaries for the outcomes of interest. Thematic framework analysis is used for in-depth interviews and focus group discussions. An inductive approach is used to analyse unstructured ethnographic observations.

### Patient public involvement

Patients and the public were not involved in the designing/writing protocol for pre-implementation phase of ASSET. However, extensive participatory methods that involve both the patients and public will be used in this phase of research to design, select and evaluate the HSSI for ASSET.

### Ethical considerations

All work packages have received separate ethics approval from the Research Ethics Committee at Kings College London (KCL) as well as the relevant country institutional and local government ethics review committees. See Appendix 2 for details for the different work packages.

## Discussion

HSS for universal health coverage with high-quality care requires the critical engagement with policy makers, researchers, service providers, and patients from the onset, to co-design interventions using high-quality, routinely available data that is responsive to the changing requirements of the users and health systems.(72) However, the current approach to strengthen heath systems in LMICs, is failing to meet these demands and is demonstrated by vertical programmes and academic research initiatives having little impact on broader health systems.(72)

ASSET is a health system strengthening programme that involves the participatory design and evaluation of a set of contextually appropriate HSSIs across three healthcare platforms, within are eight work packages, in four countries in sub-Saharan Africa. Each work package addresses complex public health issues that are relevant to local requirements and contexts. Such a diverse programme requires a flexible approach to develop a set of HSSIs tailored to the local context.

This protocol describes how robust and extensive formative research methodologies are applied to identify limitations in the delivery of, and access to, quality care. A strong emphasis is placed on engagement of relevant stakeholders and embedding ASSET within the health systems from the onset, including people with health conditions, their carers, communities, clinicians and policy makers. It is anticipated that the use of participatory methods through group and individual consultations, and ToC workshops at various stages of the pre-implementation phase, helps to foster partnership and local ownership for the different interventions that are acceptable and feasible to implement, responding to the patient needs, that be sustained in the longer term. The COVID-19 pandemic demonstrates how ASSET has embedded itself within the health system whereby ministries have engaged with the different work packages to help manage the crisis.

A critical component of the pre-implementation work is mapping the care pathway into and through health services that allows the work we do to be people-centred, facilitating more compassionate conversations about how and why health systems fail people. We emphasise health systems as opposed to health providers because health systems are also failing people who provide the care, making it extraordinarily difficult to deliver care, let alone a respectful, people–centred approach. Mapping care pathways also helps to engage stakeholders, facilitating the co-production of HSSIs. Couching problems in systems language and using patient narratives to humanise them helps to ensure the health systems strengthening is inherently people-centred.

ASSET is also investing heavily in capacity building for HSS. Extensive training is provided in implementation science and other methodologies that invites a wide range of stakeholders both from the ASSET programme as well as the wider community. Training on the different methodologies for HSS not only ensures comparability of findings across different work packages and platforms with hopeful generalisability, but importantly increased capacity for research led HSSI within these countries.

The extensive process ASSET is undertaking in the pre-implementation phase, is in part due to the absence of high-quality data available in the HMIS that can be used to inform the requirements for HSS relevant to the local context. However, this process may have negative bearing on short-term outcomes as it puts delivery of the entire programme of work at risk where completed evaluations with follow-up of adequate duration to influence policy/ practice may not be delivered. Nevertheless, engaging in these activities is critical if HSS interventions seek to bring evidence-informed care to scale in a sustainable manner.(72)

At the end of the pre-implementation phase of ASSET, it is hoped the common approach taken across different countries, care platforms and health conditions will facilitate cross platform learning and understanding of how differences in health systems and broader contextual influences shaped the development of the interventions. The overarching expectation is that by using an in-depth participatory process to engage with the stakeholders and map care pathways to and through the health system, we develop a HSS programme that can be implemented at scale that meets the needs and priorities of the local community. Ultimately it is hoped that this approach will provide people– centred high quality care that is resilient to the changing dynamic of an aging population that can also prevent future shocks like Ebola and COVID-19.

## Supporting information

Appendix 1 individual work package protocols for pre-implementation phase of research

Appendix 2 ethics approvals

## Data Availability

Data available upon request

## Abbreviations

ASSET: Global Research Unit on Health System Strengthening in sub-Saharan Africa
(ENACT): enhancing assessment of common therapeutic factors
HMIS: Health Management Information System
HSS: Health systems strengthening
HSSI: Health systems strengthening interventions
LMICs: Low- and Middle-Income Countries
NIHR: National Health Institute of Research
NCDs: Non-communicable diseases
PRIME: Programme for Improving Mental Health Care
SDG: Sustainable Development Goals
SOSAS: Surgeons Overseas Assessment of Surgical Need
TB: Tuberculosis
ToC: Theory of Change
UHC: Universal Health Coverage
WHO: World Health Organization
WP: Work package

## Declarations

### Patient consent

Protocol paper so consent not applicable

### Data sharing statement

No additional data available as a protocol paper

### Ethics approval

See appendix 2

### Authors contributions

NSeward wrote the first and subsequent drafts of the paper

MP, CH and NSeward conceptualised the idea for the paper

CH, and NSeward reviewed and edited all drafts of the paper

MP offered overall guidance

All other authors (CL, AAbulahi, ZA, AAlem, RA, MB, BB, NB, DC, RC, JD, AD, F, SF, JG, WG, RH, MK, AL, CL, KN, JM, IP, RP, JS, AT, GT, AR, NSevdalis, RV, CW) edited and offered input to various drafts of the paper

## Supplementary material

**Appendix 1: Protocols for individual work packages**

**Appendix 2: Ethics approval numbers from relevant local ethics review committees as well and Kings College London.**

