## Appendix 1 individual work package protocols for pre-implementation phase of research for "HeAlth System StrEngThening in four sub_Saharan African countries (ASSET) to achieve high-quality, evidence-informed surgical, maternal and newborn, and primary care: protocol for pre-implementation phase studies"

### Health system strengthening in sub-Saharan Africa (ASSET) Work Package Research Protocols

### ETHIOPIA: WORK PACKAGES 1 – 3

**Study Design:** Implementation research, health systems research, mixed qualitative and quantitative methods

#### Investigators

**Overall Principal investigator & Lead PI for (1) Integrated NCD/mental health, and (2) Violence against women**

Dr Charlotte Hanlon, CDT-Africa, College of Health Sciences, Addis Ababa University (AAU), Ethiopia;

**Lead co-PI (cross-project)**

Professor Martin Prince, Global Health Unit for Health System Strengthening, King's College London, UK.

**Co-investigator (cross-project, policy)**

Dr Desalegn Tegabu, Director, Policy and Planning Directorate, Federal Ministry of Health of Ethiopia


**Co-investigator (cross-project: statistics)**

Dr Girmay Medhin, Aklilu-Lemma Institute of Pathobiology, Addis Ababa University


**Co-investigator (integrated primary care)**

Dr Abebaw Fekadu, Director, CDT-Africa, College of Health Sciences, Addis Ababa University (AAU), Ethiopia;

**Co-investigator (integrated primary care)**

Dr Helen Yifter, Department of Internal Medicine, School of Medicine, College of Health Sciences, AAU, Ethiopia;

**Co-PI (NCD/mental health)**

Professor Atalay Alem, Department of Psychiatry, School of Medicine, College of Health Sciences, AAU, Ethiopia;

**Co-investigator (NCD/mental health)**

Dr Tewodros Haile, Division of Pulmonary and Critical Care, Department of Internal Medicine, School of Medicine, College of Health Sciences, AAU, Ethiopia;

**Co-investigator (NCD/mental health)**

Dr Lara Fairall, Knowledge Translation Unit, University of Cape Town, South Africa


**Co-investigator (maternal healthcare)**

Dr Solomon Shiferaw, School of Public Health, College of Health Sciences, AAU, Ethiopia.


**Lead PI (maternal healthcare)**

Dr Ahmed Abdella, Department of Obstetrics and Gynaecology, School of Medicine, College of Health Sciences, AAU, Ethiopia;

**Co-investigator (maternal healthcare)**

Professor Andy Shennan, Department of Women's Health, King's College London, UK.


**Co-investigator (maternal healthcare)**

Professor Jane Sandall, Department of Women's Health, King's College London, UK.


**Co-investigator (violence against women)**

Dr Negussie Deyessa, School of Public Health, College of Health Sciences, AAU, Ethiopia.


**Co-investigator (violence against women)**

Professor Louise Howard, Section of Women's Mental Health, Institute of Psychiatry, Psychology and Neuroscience, King's College London.


**Co-investigator (violence against women)**

Dr Roxanne Keynejad, Section of Women's Mental Health, Institute of Psychiatry, Psychology and Neuroscience, King's College London.


**Co-investigator (maternal healthcare)**

Dr Tanya Robbins, Department of Women's Health, King's College London, UK.


**Lead PI (surgery)**

Dr Andualem Deneke, Department of Surgery, School of Medicine, College of Health Sciences, Addis Ababa University;

**Co-PI (surgery)**

Dr Amezene Tadesse, Consultant General Pediatric Surgeon, Tikur Anbessa Hospital, Addis Ababa University


**Co-investigator (surgery)**

Dr Abebe Bekele, Department of Surgery, School of Medicine, College of Health Sciences, AAU, Ethiopia;

**Co-investigator (surgery)**

Dr Andy Leather, Director, King's Centre for Global Health, King's College London


**Co-investigator (surgery)**

Professor Justine Davies, King's Centre for Global Health, King's College London


**Co-investigator (Surgery/dental)**

Dr Wondwossen Fantaye, Head, Department of Dentistry, School of Medicine, College of Health Sciences, Addis Ababa University,

**Co-investigator (Surgery)**

Professor Jennifer E Gallagher, King's College London Dental Institute, King's College London


### Executive summary

#### Background

Health system strengthening is needed in order to achieve quality, integrated continuing healthcare and improved patient outcomes. In Ethiopia, the Federal Ministry of Health is introducing evidence-based guidelines for common conditions encountered in primary care (the Ethiopia primary health care clinical guidelines). The guidelines have a particular focus on non-communicable diseases (NCDs), mental health and women's health. TheMinistry of Health has also launched a programme (Saving Lives Through Safe Surgery; SaLTS) focused on improving access to quality surgical care. To fully realise the potential of these important initiatives, we need high quality evidence on effective interventions to strengthen the health system around these programmes.

#### Overall aims

The overall aim of ASSET (health system strengthening in sub-Saharan Africa) is to develop effective health system strengthening interventions to support the translation of clinical evidence into delivery of integrated continuing care at scale across healthcare platforms for NCDs, mental and substance use problems, surgical and dental care, and maternal healthcare in Ethiopia.

#### Specific aims

For NCDs/mental healthcare, maternal healthcare and surgical/dental care:

(1) To identify the health system bottlenecks to delivery of integrated, quality continuing care
(2) To adapt, pilot and refine health system strengthening interventions based on (A) defining care pathways, (B) electronic information systems, (C) quality improvement, (D) person-centred care), and (E) workforce strengthening.
(3) To implement and evaluate the impact of these health system strengthening interventions upon patient outcomes.

#### Methods

The diagnostic phase of ASSET will include the following:

(1) Theory of Change workshops with key stakeholders
(2) Situation analysis based on data available in the public sector
(3) Process mapping facility-based studies to understand pathways through care
(4) Observational studies of person-centred care and fidelity to treatment guidelines
(5) Surveys of patient knowledge and experience of person-centred care
(6) Cross-sectional facility-based studies of health worker detection of focus conditions
(7) Community survey of surgical conditions: burden, help-seeking and barriers to care

(8) Qualitative studies with key stakeholders

During the piloting phase, health system strengthening interventions will be adapted, piloted and assessed using process data and qualitative studies.

In the implementation phase, the impact of adapted health system strengthening interventions upon patient level outcomes will be evaluated.

#### Expected results

The diagnostic phase will identify health system bottlenecks to the implementation of integrated primary and maternal healthcareand increased access to quality surgical care. The output will be a comprehensive integrated district level plan.

The overall ASSET study will provide rigorous and relevant evidence on effective and feasible health system strengthening interventions to support integrated primary care and quality surgical care.

#### Funding

Funded through a grant from the UK National Institute of Health Research (NIHR)

#### Keywords

Health system strengthening; implementation research; quality improvement research; mhealth; global surgery; mental health; substance use; violence against women; non-communicable diseases; maternal health

### Overall background

ASSET-Ethiopia is a collaborative project between Addis Ababa University, King's College London, University of Cape Town and the University of KwaZulu-Natal. The focus of ASSET-Ethiopia is on the development and evaluation of interventions to strengthen the Ethiopian health system to support quality, integrated, continuing care in the areas of integrated primary care for non-communicable diseases (NCDs), mental health problems and substance use disorders, maternal health, health consequences of violence against women (VAW) and surgical and dental health.

#### Health system challenges and opportunities in Ethiopia

The Ethiopian health system is in transition. In the Federal Ministry of Health (FMoH) 20-year vision for the health system (1), primary care is placed at the centre of efforts to shift the health system from an acute care focus to better respond to the increasing burden of chronic disorders. In the Health Sector Transformation Plan (for the years 2015/16 to 2019/20), new emphasis is given to achieving quality, person-centred, respectful and equitable health care(2). Health system strengthening is required to achieve these changes, including employing electronic data capture to allow better use of information flows through the system and equipping health workers to deliver compassionate and respectful care (CRC).

A major new initiative of FMoH to support integrated and quality care is the introduction of evidence-based, primary care treatment guidelines. This is being done using the Practical Approach to Care Kit (PACK)(3), adapted to become the Ethiopia Primary Health Care Clinical guideline (Ethiopia PHC Guideline). The Ethiopia PHC guidelineis a fully integrated set of treatment guidelines for chronic disorders (including chronic infectious conditions, such as HIV and TB, as well as non-communicable diseases, mental health and substance use disorders) and women's health to be delivered by primary care-based staff. PACK was developed by the

Knowledge Translation Unit at University of Cape Town and includes the treatment guidelines, as well as a programme of facility-based learning for primary care providers, mentoring from dedicated PACK trainers and a focus on health system strengthening to support sustainability. The guidelines are continuously updated by 'best evidence' obtained in collaboration with the BMJ Best Practice unit. They are then contextualised for low- and middle-income countries at UCT. FMoH is working with UCT and BMJ to further contextualise the guidelines for Ethiopia, with a view to scaling up across the whole country.

PACK has been implemented in South Africa, Botswana, Nigeria and Mexico, and has been shown to improve treatment outcomes for people with HIV and TB (4)(5). In a recently published pragmatic cluster randomised controlled trial, PACK did not lead to significantly improved treatment practices by primary care workers for diabetes, hypertension, chronic respiratory illness and detection of depression (6). Nonetheless, there was no evidence of harm arising from this task-shifting model. This recent evidence for PACK highlights the challenge of effecting change through implementation of treatment guidelines without associated health system change.

The Ethiopia mental health research group has expertise in implementation research and health systems research in relation to scaling up mental health care through integration within primary care(7)(8). The key lesson learned from mental healthcare scale-upaccords with the experience implementing PACK: short-term training of primary healthcare workers has limited success unless supported by health system strengthening interventions (HSSIs) (7).

#### Ethiopia Primary Healthcare Clinical Guideline for integrated care

The Ethiopia primary health care clinical (PHC) guidelineis structured according to common symptomatic presentations in primary care, e.g. headache, fatigue, sleeping difficulty, and then follow algorithms through possible diagnoses, starting with those requiring an emergency response. Such an approach has the potential to promote an integrated approach to care, for example, prompting healthcare workers to consider mental health and substance use problems even when patients present with physical symptoms (a common presentation of depression in Ethiopia). The Ethiopia PHC guideline has been contextualised and the training of country-level Master trainers will take place in January 2018. Training will then be cascaded down, first to regional level Master trainers, and then to district and facility-level trainers. Working with the Federal Ministry of Health, the facility-based training will commence in the ASSET study districts in July 2018. The facility-based training emphasises on-the-job learning. Every week, the health centre-based healthcare workers will meet together for two hours to work through case-based training materials which are linked to the Ethiopia PCC guidelines. In the ASSET districts, the facility-based training sessions will first cover the case-based training related to NCDs, mental health, substance use and maternal healthcare.

#### ASSET health system strengthening approach

In the ASSET project, therefore, we will use implementation and quality improvement (QI) research to address barriers to translating clinical evidence within the Ethiopia PHC guideline) into delivery of integrated continuing care at scale. Our thesis is that a) task-shifted care, a key universal health coverage strategy, needs effective ongoing training, mentoring, and support, and that b) guideline-based care is essential, but insufficient to improve effective coverage. Hence, in ASSET, we will work across healthcare platforms for surgery, maternal healthcare and NCDs/mental health and substance use problems to develop, adapt and evaluate health system strengthening interventions (HSSI) to enhance delivery of guideline based care.

Core domains of HSSI and quality improvement for continuing care are well established, from research, and consensus(9). Substantial evidence supports effectiveness of domains of the Chronic Care Model (Delivery System Design, Self-Management Support, Decision Support, Clinical Information Systems and Healthcare organization)(10). For resource-poor settings in sub-Saharan Africa, we operationalise these into 5 HSSI elements:

1. evidence-based care pathways for integrated primary care for NCDs, mental health and substance use problems, maternal care, and high volume surgical procedures, with practice-based training, ongoing mentoring and support for implementation
2. Health Management Information System (HMIS) data to monitor and improve the quality of continuing care and patient outcomes, with specific focus on measuring detection, engagement on care pathways, adherence, retention and ‘treatment to target’
3. a peer-driven QI culture, with motivated healthcare workers taking responsibility for improving care processes and outcomes (a ‘learning health systems approach’)
4. person-centred care reflecting patient values and preferences, with agreed care plans and patient empowerment for self-management
5. workforce strengthening to prepare for necessary organisational change, and foster non-technical skills (e.g. leadership, teamwork, communication, counselling, and stress management).

There is evidence that these approaches can work, but, in sub-Saharan Africa, limited mainly to demonstrationprojects of single elements and rarely documented and evaluated with rigour. In ASSET, HSSI will be adapted to local contexts, piloted and refined, and then applied as a package of measures to improve care delivery.

#### ASSET projects within Ethiopia

The ASSET team within Ethiopia brings together researchers and the Federal Ministry of Health with the goal of carrying out high quality implementation research which can contribute directly to improved patient outcomes. The rationale for the selection of the three focus areas (integrated primary care for NCDs/mental health/substance use problems, maternal healthcare and surgical/dental care) is that each requires a continuing care approach in order to optimise patient outcomes. The projects will be linked by (1) the methodological approach (a diagnostic phase, piloting and patient-level evaluation of impact), (2) the types of HSSI that we will implement (as described above), and (3) the geographical area (the Gurage Zone of the Southern Nations, Nationalities and Peoples’ region of Ethiopia). Dr Hanlon will provide overall direction across all projects to ensure methodological consistency, and will be PI for the integrated primary care and violence against women studies. Drs Abebe Bekele and Andualem Deneke are theco-PIs for the surgical and dentalstudy and Drs Solomon Shiferaw and Ahmed Abdella are co-PIs for the maternal healthcare study.

#### Significance of the study

Stand-alone training of health workers is unlikely to achieve sustainable benefits to patient care. Attention needs to also be paid to the health system strengthening needed to support programme implementation. In the ASSET diagnostic phase, we will use rigorous methods to identify existing health system bottlenecks to the delivery of programmes of care for integrated PHC for NCDs/mental health/substance use problems, maternal healthcare and surgical/dental care. The evidence we generate will be of direct relevance to work being conducted by the Federal Ministry of Health of Ethiopia for scale-up of Ethiopia PHCG, and implementation of the Saving Lives Through Safe Surgery (SaLTS) initiative and programmes to reduce maternal mortality. The evidence generated will also inform the development of health system strengthening interventions to overcome the bottlenecks, which will then be evaluated in the next phase of ASSET. As there have been few studies in Ethiopia examining health system interventions with rigorous methods, our study has the potential to contribute to the knowledge base, as well as build capacity for health systems research in a large multi-disciplinary team of Ethiopian clinicians. Our involvement of key stakeholders through a participatory process will ensure the relevance of our findings and increase their likely impact.

### Overall aim of ASSET

The overall aim of ASSET is to develop effective health system strengthening interventions to support the translation of clinical evidence into delivery of integrated continuing care at scale across healthcare platforms for NCDs, mental and substance use problems, surgical and dental care, and maternal healthcare in Ethiopia.

### Overview of methodological approach

The general methodological approach will involve (1) a diagnostic phase to identify care pathways, the main health system bottlenecks, acceptability and feasibility of potential system strengthening interventions and development of a district level integrated healthcare plan, (2) a piloting phase, and (3) an implementation and scale-up phase.

In this application, we are seeking ethical approval for the diagnostic and piloting phases of ASSET.

Each of the ASSET studies will now be presented in turn.

### Study 1: Integrated primary care for NCDs, mental health and substance use problems

#### <u>**Study 1: Background**</u>

##### *Links between NCDs and mental health and substance use problems*

There are substantial inter-relationships between the non-communicable diseases of hypertension, diabetes mellitus, cardiovascular disease and chronic respiratory disease, and mental health and substance use problems (11). These conditions share many social, cognitive and environmental determinants, and are risk factors for one another. For example, depression is a risk factor for development of hypertension, but can also be a consequence of hypertension (and the medications used to treat hypertension, e.g. propranolol), and common risk factors (e.g. poverty, physical inactivity) may predispose to both conditions. There is thus high co-morbidity between these conditions, and even multi-morbidity (where more than two conditions are present in the same individual). The consequences of undetected mental health and substance use problems in people with NCDsinclude low quality of life and increased disability(12), poorer adherence to medications, worse self-care and lower uptake of beneficial lifestyle changes, culminating in worse prognosis and poorer outcomes e.g., increased risk of blindness, amputation and mortality in persons with diabetes mellitus(11).

##### *Integrated care for people with NCDs, mental health and substance use disorders*

Despite the high degree of overlap of these conditions, the health system response in most low- and middle-income countries is typically focused on providing care for each disorder in isolation. Indeed, mental health and substance use co-morbidity with NCDs is often overlooked altogether. In the last five years, the Federal Ministry of Health of Ethiopia has published a National Mental Health Strategy (2012/2016, currently under revision) (2)and a National Strategic Action Plan for prevention and control of Non-Communicable Diseases (13) in Ethiopia. At present these two policy documentsboth focus on integration of care within primary care to expand access to services, but have been implemented vertically, with almost no cross-talk between the two.

There is strong evidence forbenefits of health service configurations that support integrated and co-ordinated care for people with chronic health conditions. The Chronic Care Model was developed in the USA and has been shown to be associated with improved quality of care and better patient outcomes (10). The Chronic Care Modelseeks to achieve *“productive interactions between informed activated patients and prepared proactive practice*”. This is achieved through the following:

strengthening patient self-management, decision support for healthcare workers, delivery system redesign (e.g. team-based working, case management and task sharing) and clinical information systems. Underpinning this approach is a commitment to person-centred care. The Chronic Care Model was adapted for LMICs to form the ‘Innovative Care for Chronic Conditions (ICCC) framework’(14).Rather than focusing on the patient-healthcare worker dyad, the ICCC framework expands the focus to include family and community partners. There is greater emphasis on the need to ‘prepare’ healthcare workers for the new roles, a shift from decision support to ‘organised and well-equipped’ healthcare teams and more focus on the need for co-ordinated and continuing care.

In ASSET, we have operationalisedthe key health system strengthening interventions needed to support care within the ICCCF as follows: (1) establishment of evidence-based care pathways, (2) utilisation of Health Management Information System (HMIS) data for detection, engagement on care pathways, adherence, retention and ‘treatment to target’, (3) embedding a peer-driven culture of quality improvement, with involvement of service users to develop a learning health system, (4) person centred care and strengthened self-management, and (5) workforce strengthening to prepare for necessary organisational change and foster non-technical skills.

##### *Existing work on health systems to support chronic care in Ethiopia*

In response to the plan to scale up mental health services across Ethiopia, there have been several initiatives which inform the need for HSSIs to improve integrated care. In the Programme for Improving Mental healthcarE (PRIME), detection of depression by health centre-based nurses and health officers was extremely low at baseline and showed little improvement after stand-alone training of healthcare workers (15). Exploratory work indicates that high levels of stigma, unmet emotional needs of healthcare workers themselves and feeling ill-equipped to shift from a more paternalistic clinician-patient relationship are important factors contributing to reluctance to diagnose depression(16). In PRIME, as well as the TaSCS trial (task-sharing for care of severe mental disorders), integrated primary care-based services for people with mental disorders have been characterised by high levels of initial engagement followed by high levels of drop-out from care or a pattern of intermittent engagement. Identified reasons include inadequate understanding of the need for ongoing care in patients (deficits in equipping patients for self-management) and a health system which does not support proactive planning of care and co-ordination with health extension workers to provide community outreach (17).Low quality care and a failure to ‘treat-to-target’ i.e. adapt care according to response with a goal of achieving recovery, are also emerging as important concerns. As part of the Emerald project (emerging mental health systems in low- and middle-income countries, we identified a lack of chronic care orientation in both patients and healthcare workers, with few components of good practice in operation in the study health centres. In Emerald, we also identified a low baseline level of service user involvement in service development and improvement, but a willingness and enthusiasm to pursue this approach (9).

#### Study 1: Diagnostic phase

##### *Study 1: Diagnostic phase aims*

Following the introduction of the Ethiopia primary health care guidelines (PHCG) at health centre level for people with NCDs and depression/substance use disorders:

(1) Map out existing care pathways
(2) Identify health system bottlenecks for

    a. the delivery of person-centred care, and for
    b. detection, engagement on care pathways, adherence, retention in care and treatment to target

(3) Explore acceptability and feasibility of potential health system strengthening interventions (HSSIs) to address the barriers.
(4) Participatory development of a district-level integrated primary healthcare plan incorporating HSSIs.

The diagnostic phase comprises the following activities: theory of change workshops, situation analysis, facility process mapping and quality of care study, facility-based cross-sectional study and a qualitative study.

##### *Study 1: Setting*

The ASSET maternal healthcare study will take place in Meskan, Mareko and Sodo districts of the Gurage Zone, Southern Nations, Nationalities and Peoples' Region of Ethiopia. These districts are located around 100 to 130km south of the capital city, Addis Ababa. The Butajira Health and Demographic Surveillance Site (HDSS) is nested within the Meskan and Mareko districts, comprising 10 sub-districts (*kebeles*) where the population has been enumerated since 1987, established as part of the Butajira Rural Health Programme (18). There is one general hospital (Butajira hospital), one primary hospital (Buee hospital), 18 health centres (8 in Sodo, 7 in Meskan, 3 in Mareko) and 125 health posts (58 in Sodo, 41 in Meskan and 26 in Mareko) serving the population of the three districts.

##### *Study 1: Theory of Change workshops*

Theory of Change (ToC) is a participatory approach that is employed to develop a contextually valid roadmap to achieve a shared programme outcome(19). Stakeholders are brought together in workshops. The first task is to build consensus on the most important outcome of the programme. The group of stakeholders then work backwards to map out the intermediate outcomes that need to be achieved on the way to the final outcome. The interventions needed at each step in the pathway are determined by reviews of the existing evidence base combined with local experience. Assumptions underlying the steps on the pathway are made explicit and investigated using research studies. Indicators of success are also specified. The output of ToC is a map which specifies the necessary elements of a complex interventions, the approach to implementation and the indicators that need to be evaluated. In this way, the ToC process enables stakeholders to come to a common understanding of what the interventions will achieve, and how they will achieve their goal. The ToC map also provides a framework for evaluation of the complex interventions.

The ToC approach has been used previously in one of the study districts in Ethiopia to guide development, implementation and evaluation of a district level plan to integrate mental health into primary care(8).

For the integrated primary are study, we will conduct ToC workshops at both the district level/zonal level and at the regional/national level in order to capture the relationship between central policies/plans and local implementation. The purpose of the workshops, their timing and planned participants are detailed in Table 1. The sample size will be approximately 50 persons. Theory of Change workshops need to involve key stakeholders but also need to be manageable in size to allow full participation (not more than 25 persons per workshop).

Inclusion criteria:

- Key stakeholders for the topic under discussion
- Able to converse in Amharic, the official language in the study districts
- Providing informed consent

Exclusion criteria: none.

Informed consent will be sought from ToC participants (See Appendix 5 for information and consent forms) and the workshops will be audio-recorded. The outputs of the ToC workshops will be a ToC map and a report of how key decisions were made in order to agree on the ToC map.

*Table 1: ToC approach for integrated care for NCDs, mental health and substance use problems*

| Level | District/zonal level | Regional/national level |
|---|---|---|
| Participants | *Organisational:* District and zonal health office heads/focal persons for NCD/mental health care (Mareko, Meskan, Sodo), CEOs/medical directors of Butajira/Buee hospitals<br>*Facilities:*Health centre-based managers and clinicians (health officers, nurses)<br>*Community:* Community leaders, community representatives from hospital boards, health extension workers, religious leaders, relevant local NGO leads | *Policy:* FMoH NCD unit head and mental health focal person, Clinical Services Directorate, Health Extension Programme directorate, Quality directorate, health information systems.<br>*HSSI implementers:* organisations working in mHealth, quality improvement, leadership training (Institute of Primary Care)<br>*Medical:* specialists in NCD and mental health/substance use care,<br>*Civil society groups:* mental health society of Ethiopia, patient representatives from NCD-linked organisations. |
| Timing | Workshop 1: following district situation analysis<br>Workshop 2: following completion of diagnostic phase studies | |
| Purpose | Workshop 1: develop preliminary ToC map to identify gaps in understanding which need to be addressed in the diagnostic phase studies<br>Workshop 2: finalise district level perinatal health care plan, with process indicators and outcomes | |

#### *Study 1: District level situation analysis*

We will collect data available in the public domain to inform our understanding of the current district level situation (community, primary care facility and organisational level) with respect to integrated NCD, mental health and substance use care. We will adapt a situation analysis used previously by the study team in the ASSET study sites in relation to integrated mental healthcare(20). The focus of the situation analysis will be on identifying structural system barriers to the integrated care packages and pathways specified in the Ethiopia PHC guidelines, e.g. in terms of available personnel, equipment, medications, information systems, service structure and policies and plans. See section **Error! Reference source not found.**.

#### *Study 1: Facility cross-sectional study*

*Research Questions*

In people seeking routine out-patient care at primary care health centres where healthcare workers have been trained in the Ethiopia PHC clinical guidelines:

- What is the prevalence, co-morbidity/multi-morbidity and level of treatment control for the focus NCDs (hypertension, diabetes mellitus, cardiovascular disease, chronic respiratory disorder), depression and substance use disorders?
- What is the level of health worker detection of NCDs, depression and substance use disorders?
- What is the level of knowledge, awareness, preferred help-seeking and person-centred care of people with detected NCDs, depression or substance use disorder about the conditions?
- What are the unmet needs for family planning in women?

*Study design*

We will conduct a two-phase facility-based cross-sectional survey.

*Sampling and sample size*

Sample 1 (from phase 1): detection of NCDs in out-patient

We will seek to recruit consecutive adult attendees at (1) the general out-patient clinicsat primary care health centres in Sodo, Meskan and Mareko districts, Gurage Zone. We will select health centres with a volume of out-patient attendees greater than 25 per month for feasibility.

For the out-patient clinic study, the parameters to guide calculation of sample size are not known. Using varying estimates of detection and prevalence, with a power of 80%, and alpha of 0.05, and the sample divided equally between 8 health centres, the precision around the detection estimate would vary between 6 and 8% for a sample size around n=2000.

*Table 2: Sample size estimations for out-patient NCD detection study*

| Estimated prevalence of hypertension | Estimated detection | Precision | Sample size for people with hypertension | Screened sample | Design effect | Final sample size |
|---|---|---|---|---|---|---|
| 10% | 10% | +/-6% | 96.0 | 960 | 2.19 | 2102 |
| | 20% | +/-8% | 96.0 | 960 | 2.19 | 2102 |
| 15% | 10% | +/-6% | 96.0 | 640 | 1.79 | 1145 |
| | 20% | +/-7% | 125.4 | 836 | 2.04 | 1701 |
| 20% | 10% | +/-6% | 96.0 | 480 | 1.59 | 763 |
| | 20% | +/-6% | 170.7 | 855 | 2.06 | 1760 |

For the sample size, the following formula was used: $N = (z^2 \times p\,(1-p))/\delta^2$.

There are no published studies from Ethiopia to guide us in our estimation of the intra-cluster correlation (ICC) for detection by health workers at a specific health centre. We therefore estimate the ICC to be 0.01, the median ICC from a review of 31 cluster-based interventions in primary health care (Adams et al. 2004).

Patient flow in the out-patient clinics at the 8 largest health centres in the study area varies from around 400 to 1000 patients per month. We estimate, therefore that recruitment will take two to three months.

Sample 2 (from phase 2): patient knowledge about NCDs and experience of NCD/mental health care

People who are identified in the OPD and ANC surveys as having an NCD will be invited to attend a clinic on a specified day in order to carry out patient surveys of knowledge and experience of NCDs/mental health care and observational studies (as described in the next section). This will include (1) people with established NCD diagnoses, (2) people with a new PHC worker diagnosis on an NCD on the day of the OPD survey, and (3) people detected as having an NCD who are not detected by the PHC workers.

*Inclusion criteria*

- Willingness to provide informed consent
- Adult: aged 18 years or above
- Fluent in the language of the interview (Amharic)
- Able to understand the interview (for example, excluding people with severe intellectual disability or dementia)

*Exclusion criteria*

- Requiring treatment for medical emergency. Health centre clinicians will be asked whether or not any of the potentially eligible attendees requires treatment for a medical emergency.
- Pregnant, as reported by the woman.

*Recruitment and study procedures*

Participants will be informed about the study and invited to participate by project data collectors while they are waiting for their appointment. See appendix 5 for information and consent forms. If they consent to participate, they will first see the clinician and then participate in an interview with the data collector using fully structured questionnaires, followed by a clinical assessment by project nurses. If the person is found to have abnormal clinical findings, this information will be provided to the healthcare provider after completion of the project assessment to inform further assessment.

*Measures*

Fully structured lay interviewer administered measures for all participants. See Appendix 1.

- **Sociodemographic characteristics:** age, gender, educational level, place of residence (rural/urban), exposure to indoor air pollution
- **Presenting complaint:** the reason that the person attended the facility
- **Number of contacts with health services:** Health centres; healthcare extension workers
- **Medical history:** any existing diagnosis of hypertension, diabetes, asthma, chronic obstructive pulmonary disease, cardiovascular disease (heart attack, chest pain or stroke).
- **Family history:** History of cardiovascular disease.
- **Obstetric history:** No of pregnancies and outcome for each; currently on family planning and what modality and where they receive this.
- **Habits:** current smoking and number of cigarettes per day, number of cups of coffee per day
- **Respiratory symptoms**: the IUATLD Respiratory Questionnaire(21).
- **Depression symptoms**: these will be measured using the Patient Health Questionnaire, 9 item version. The PHQ-9 has been validated previously in out-patient attendees to primary care health centres in the Butajira area, with a cut-off of 5 and above giving the best sensitivity (83.3%) and specificity (74.7%) for detecting major depressive disorder.
- **Substance use:** this will be measured using the World Health Organisation ASSIST questionnaire (22). The ASSIST has been used previously in the Ethiopian setting following rigorous translation procedures and adaptation of references for a 'typical drink' (23). The ASSIST allows classification of level of risk for a substance use disorder (low, medium and high) based on the total score. The ASSIST will be used to detect alcohol and khat use disorders.
- **Disability:** this will be measured using the World Health Organisation Disability Assessment Schedule, version 2.0, 12-item (24). This measure has been validated for use in the study area and found to be sensitive to change (25).

Project clinical assessments on all study participants:

- **Blood pressure:** this will be measured using the CRADLE device, which allows automated measures of blood pressure. Three blood pressure readings will be carried out, each at least two minutes apart while the person is seated, following standard practice.
- **Fasting glucose or HbA1c:** this will be measured using a glucometer to measure fasting glucose with a sample of capillary blood or a point-of-care measure of HbA1C if the person is not fasting.
- **Spirometry:** this will be carried out on all OPD study participants who have not been diagnosed as having TB or who have symptoms of TB (cough/difficulty breathing for > 2 weeks, weight loss and

night sweats). Disposable cardboard mouth-pieces with one-way valves will be used to ensure there is no infection risk.

- **Anthropometry:** Height/ weight/ waist circumference.

*Clinical records*

After the person has been seen by the clinician, the following data will be extracted from the routinely recorded clinical notes: presenting complaint, diagnoses, investigations requested, medication prescribed, other treatments, advice given, timing of next follow-up visit.

*Data collection and management*

Data will be collected using mobile devices with the Open Data Kit (ODK-collect) software and uploaded on the AAU server owned by the School of Public Health. The data which will be available for immediate aggregation and analysis and feedback before data collectors leave the field. The data will be kept on a virtual Server dedicated for this purpose which will be password protected to ensure safety of the data. Only study team members and individuals/stakeholders whose request is approved will have access to the data.

The lay data collectors will have a minimum of diploma level education and will be trained for 3 days in the questionnaires, with a further two days of observed field practice.

The clinical assessors will be nurses or health officers, with a minimum qualification level of diploma.

A supervisor will be present in each of the facilities, to oversee data collection and ensure that sampling procedures are implemented as per the protocol. Electronic data entered onto the tablets will be transferred to password-protected computers of a secure server which can be accessed by data managers based in Addis Ababa. The data managers will check the data for consistency and outliers at least once per week.

We will pay 50 Birr ($2) as time compensation for participants to compensate for their time and any additional expenses incurred by spending extra time at the facility.

*Data analysis*

The presence of a clinical abnormality indicative of a focus NCD condition will be determined using the criteria presented in Table 3, on the basis of the clinical assessments. The electronic data entry device will then instruct the assessor to carry out further assessment, depending on the condition present.

*Table 3: Cut-offs for defining NCDs, NCD risk factors and mental health problems*

| | |
|---|---|
| Body mass index | Overweight: 25 to 29.9 kg/m$^2$<br>Obese: ≥ 30 kg/m$^2$ |
| Depressive symptoms | Scoring 5 or more on the PHQ-9 and reporting impaired functioning or diagnosis of depression from health professional |
| Risk of harmful alcohol use | On the ASSIST: moderate risk 4-26 and high risk > 26 |
| Risk of harmful khat use | On the ASSIST: moderate risk 4-26 and high risk > 26 |
| Hypertension | Systolic blood pressure ≥ 140 mmHg and diastolic blood pressure ≥ 90 mmHg or ever received a diagnosis of hypertension from a health professional |
| Cardiovascular disease | Ever been diagnosed by a health professional with a heart attack or stroke. |
| Diabetes mellitus | Random blood glucose of ≥ 7.0mmol/l (126mg/dl) or diagnosis of diabetes mellitus from a health professional |
| Chronic respiratory disorder | Abnormal Forced Expiratory Volume in one second or diagnosis of asthma or bronchitis or emphysema or chronic respiratory problems from a health professional |

Using the above definitions, we will calculate the prevalence, co-morbidity and multi-morbidity for hypertension, cardiovascular disease, diabetes, chronic respiratory disorder, depression and harmful substance use. We will calculate the % of people with an NCD, depression or substance use disorder whose condition is detected and controlled. We will conduct multi-variable analyses to identify factors associated with NCD/mental health or substance use problems, awareness of the condition and treatment control.

*Specific ethical considerations*

If the person is found to have an abnormal finding in terms of physical or mental health, the information will be given to the health worker and the person will be re-assessed where necessary.

#### *Study 1: Facility process mapping*

*Research Questions*

In health facilities where the Ethiopia PHC clinical guidelines have been implemented:

(1) What are the care processes and pathways for integrated care for people with NCDs, mental health and substance use problems?
(2) What is the quality of clinical care in terms of:
    a. Compassionate, respectful and person-centred care?
    b. Adherence to the Ethiopia PCC evidence-based guidelines?

*Study design*

Documentary analysis and observational studies.

These studies will be conducted with the permission of the health centre head and with the involvement of health centre staff.

*Setting*

Six health centres will be selected purposively, one urban and one rural from each of the three districts.

*Process mapping and documentary analysis of quality of care*

We will identify the processes of care for people with NCDs, mental and substance use problems at the health centre by discussing with the facility head and Health Management Information System (HMIS) focal person and reviewing relevant documents and reports. We will map out (1) the type of routinely recorded data available (including the extent it allows for tracking detection, engagement in care pathways, adherence, retention in care and treatment-to-target), (2) the completeness of routinely recorded data, and (3) the pathways through care experienced by people with these conditions.

In addition, we will identify clinical records from the study 1 cross-sectional survey (see below) where the person is documented to have an NCD and/or mental health and/or substance use disorder. We will then review these charts in detail to extract information on the extent to which the evidence-based care protocols in the Ethiopian PHC clinical guidelines were followed and documented. See section **Error! Reference source not found.**for the data extraction template.

*Observational study*

The observational study will examine the communication between the healthcare worker and the people attending routine out-patient care with NCDs.

With the permission of the health worker, project psychiatric nurses or clinically-qualified research workers will complete an observational assessment of the clinician's clinical communications skills (including respectful care) and holistic care of the person (including the detection and assessment of psychosocial problems). The project workers will use an adapted version of the ENACT scale (Enhancing Assessment of Common Therapeutic factors) (26), which has been adapted for the Ethiopian context and shown to be reliably administered by trained clinicians. See Appendix 1 (8.3). We will assess inter-rater reliability of the project workers before proceeding with the study.

We will purposively select 6 to 8 health centres for the quality of care study, to include both urban and rural facilities. We will aim to assess 5 consultations each for a total of 60 health care workers.

Inclusion criteria:

- Primary care worker providing out-patient care in study clinic
- Providing informed consent.

Exclusion criteria: None

#### *Study 1: Health worker survey*

In order to assess the organisational readiness of health centres for implementation of the Ethiopia PHC Clinical Guidelines, we will conduct a cross-sectional survey of health centre clinicians, managers and woreda health administrators.

*Measure*

We will use the Organisational Readiness to Implement Change (ORIC) survey. The ORIC questionnaire has not been used in Ethiopia previously. We will translate into Amharic, back-translate, produce a consensus version and then carry out cognitive interviewing with health centre workers not from the study site.

*Sample*

The survey will be carried out in all health facilities and district health offices of the three study districts (approximate sample size of n=200).

Inclusion criteria:

- Health worker/manager working in study districts
- Providing informed consent.

Exclusion criteria:

- Worked in health facility for less than 3 months. We will obtain this information from the health worker by self-report.

*Procedures*

We will provide the questionnaires in envelopes with a unique identifier and code to indicate the health facility. Respondents will be invited to complete the survey independently and return to the data collector in a sealed envelope. The data will be used descriptively, with a particular focus on examining patterns of potential barriers to change and differences across the health facilities.

There will be no payment offered to participants.

#### *Study 1: Qualitative study*

In primary care attendees with NCDs, mental health and substance use disorders:

- What are the health system barriers and facilitators to:
    - detection, engagement on care pathways, adherence, retention in care and treatment-to-target?
    - delivery of integrated, person-centred care?
- What is the acceptability and feasibility of potential health system strengthening interventions for integrated care? How could these HSSIs be best implemented to optimise care and improve outcomes?

*Study design*

Qualitative study using a framework approach and in-depth interviews with key stakeholders.

*Setting*

The Meskan, Mareko and Sodo districts of the Gurage zone.

*Sample*

We will sample a total of around 40 to 50key stakeholders purposively, focusing on the following groups of respondents:

- Primary care attendees who have been diagnosed as having an NCD, mental health or substance use disorder.
- Primary care attendees who have an undiagnosed NCD, mental health or substance use disorder
- Health care providers: health extension workers, health centre clinicians involved in general out-patient care, health centre managers, health facility staff involved in collection, synthesis and reporting of HMIS data, hospital and facility-based trainers for the Ethiopia PCC guideline implementation, hospital-based clinicians involved in NCD, mental health and substance use service delivery
- District/zonal health office leads for NCDs, mental health and substance use disorders, district, zonal and regional health office staff involved in Ethiopia PHC guideline implementation, relevant community leaders and representatives from NGOs, faith-based organisations and traditional and religious healers.

Sampling will continue until theoretical saturation is attained  or until all relevant key informants have been interviewed (e.g. in the case of district managers where numbers may be limited). The sample size is an estimate, based on the need to have enough interviews in the differing categories of respondent.

Inclusion criteria:

- Able to converse in Amharic
- Aged 18 years of above
- Able to understand the interview.

Exclusion criteria

- Not acutely unwell as judged by the qualitative interviewer or reported by the individual.

*Procedures*

Potential participants will be approached and informed about the study. They will be invited to provide informed consent by the research assistant who will conduct the in-depth interviews. The research assistant will be qualified at Masters level and will have experience in qualitative data collection. The location of the interview will be determined according to the convenience of the participant, and may be conducted in the person's home, the Butajira mental health research office, health facilities or in the outside area of a centrally-located hotel. Privacy will be ensured. With the consent of the participant, the interview will be audio-recorded. Care will be taken to convey to the participant that we are interested in hearing their perspectives, that there are not right or wrong answers and that the interview transcripts will be anonymised.

The interview will begin with collection of relevant sociodemographic data and then proceed to follow a topic guide. The purpose of the topic guide is to provide a framework for discussion, but the research assistant will conduct the interview in a conversational manner, encouraging the respondent to elaborate on their responses. The order of the topic guide sections will be flexibly administered according to the direction of the discussion. The topic guides are provided in Appendix 3, but will be developed iteratively as data collection proceeds.

The topic guides will be piloted and reviewed to ensure appropriateness before proceeding to the main study data collection. The language of the interview will be Amharic, the official language of Ethiopia. Amharic is spoken by the majority of people in the study area, although for many rural residents it is a second language. The audio files will be downloaded onto a password-protected computer. We will employ transcribers to transcribe the audio files in Amharic. The Amharic transcripts will then be translated into English. The translators will make note of any conceptually difficult translation for further discussion. Selected transcripts will also undergo independent translations to ensure the accuracy of the translation process.

Participants will be reimbursed for their time and transport (100 Birr; $4) if they are required to travel from their residence or place of work in order to participate in the interview.

*Data analysis*

We will follow a framework approach to data collection and analysis (27). The initial coding framework will be derived from the topic guide. At the time that interviews are conducted, we will keep field diaries of the interview contexts and any observations or reflections on the interview content. The English transcripts will be read as soon as they are available to inform iterative development of the topic guide and inform ongoing sampling. Two researchers (including the research assistant who conducted the interviews) will independently read through three transcripts and code text, adapting the pre-existing coding framework as needed and allowing new codes to emerge from the data. The remaining interviews will then be coded by one researchers using the coding framework, with further modification and re-coding as needed. At the mid-point of coding, there will be a researcher meeting to carry out an interim analysis and review the coding framework. Once all interviews are coded, an Excel table will be created to chart summaries of the coded text (columns) in relation to participants (rows). The chart will then be used to allow comparative analysis by type of informant (e.g. health worker vs. primary care attendee) for the major themes. The syntheses of findings for the major themes will be checked back against the original data.

*Ethical considerations*

Please see the general ethical consideration of the overall ASSET project. For this specific qualitative study, we will follow these general principles, and pay particular attention to the following potential issues. In all cases we will take care that people do not feel pressurised to participate. All participants will be remunerated for time and transport costs, where relevant. We will ensure anonymity of the interview transcripts and any quotations used in publications or reports.

#### Study 1: Pilot phase

On the basis of the diagnostic study findings, the proposed health system strengthening interventions will be adapted for the Ethiopian context. They will then be piloted in one urban and one rural health centre. The pilot evaluation will combine process evaluation and qualitative methods (observational, focus group discussions and in-depth interviews).

| | | |
|---|---|---|
| 1 | Change management/ clinical communication skills | |
| | Activities | Evaluation |
| | Training of managers | Numbers trained<br>Number of meetings with staff to facilitate change |
| | Training of facility staff | Numbers of facilitated training sessions<br>Number of staff attending training<br>Tracking of integrated care (with mHealth)<br>Service user experience of care (qualitative)<br>Health worker experience of delivering care (qualitative) |
| 2 | mhealth for chronic NCD/mental health care | |
| | Activities | Evaluation |
| | Training of key personnel | Numbers of health workers trained<br>Pre-post testing |
| | Introduction of mHealth system | Adoption by health workers (how many using mHealth)<br>Feasibility: tracking functionality of devices, need for technical support<br>Acceptability: in-depth interviews. |
| 3 | Learning health system | |
| | Activities | Evaluation |
| | Training in quality improvement | Number of health workers trained<br>Number of QI meetings |
| | Training workshops for health workers and service users | Number of health workers trained<br>Number of service users trained |
| | Establishing service user involvement in QI | Number of meetings between health workers and service users<br>FGDs and interviews |
| | Integration of mHealth into QI procedures | Number of QI meetings where mHealth data used |

The pilot phase will also involve validation of measures of person-centred care.

### Study 2: Integrated maternal healthcare

#### Study 2: Background

##### *Maternal healthcare in Ethiopia*

Ethiopia has made great strides in reducing maternal mortality, but the maternal mortality ratio is still estimated to be 412 per 100,000 live births (EDHS 2016) (28). Reduction of maternal mortality is an important target of the Sustainable Development Goals. Three conditions contribute to over half of maternal mortality: pre-eclampsia/eclampsia, sepsis and haemorrhage. Earlier detection, timely care escalation and evidence-based treatments for these conditions have the potential to reduce maternal mortality. In Ethiopia, uptake of maternal healthcare is low. An estimated 28% of women had skilled birth attendance in the latest Demographic and Health Survey 2016 (29). The percentage of women attending for four antenatal care assessments (as per current Ministry of Health guidance) was 32% (29). In addition to low autonomy of women and non-availability of transportation, disrespectful and abusive treatment from healthcare providers may be an important factor contributing to low uptake of services (30). The lack of timely uptake of emergency maternal healthcare also appears to reflect low awareness in the community. In community studies of women in Ethiopia, there are low levels of birth preparedness (31)(32), a lack of recognition of danger signs in pregnancy, childbirth and the postnatal period (33) and limited awareness of what to do in response to danger signs (complication readiness) (31).

Health system bottlenecks are likely to make an important contribution to high maternal mortality at each step in the care pathway, from the moment of detection of danger signs, through prompt referral and communication of risk, the primary care response, the timely escalation of care and the quality of definitive care available in secondary healthcare facilities. Most studies from Ethiopia have focused on the quality of routine maternal healthcare and present evidence of poor quality of care (34). There has been less focus on the quality of the health system response to emergency maternal conditions, and little is known of the key system barriers and how they may be overcome.

##### *The unmet need for integrated psychosocial care*

The psychosocial aspect of maternal healthcare has been neglected in Ethiopia and many low- and middle-income countries (LMICs) (35). This is despite evidence of higher levels of maternal depression and anxiety (36) and exposure to intimate partner violence (37) in LMICs when compared to high income countries. In Ethiopia, intimate partner violence is more frequent in pregnant women(38), with 77% of pregnant women reporting physical violence in their current pregnancy (39). Intimate partner violence is strongly linked to poorer maternal mental health, which has also been shown in studies from Ethiopia (40). The consequences of undetected and untreated maternal mental health problems and psychosocial risks (in particular exposure to violence) are substantial. Studies from Ethiopia have shown antenatal depression to be associated with increased emergency presentations in pregnancy (41), increased perinatal complications (42), prolonged labour (43) and increased use of emergency delivery care (44). Maternal depression has also been shown to have consequences for the child, with Ethiopian studies showing associations with low birth weight (45) (although some negative studies (46), delayed initiation of breast-feeding, increased risk of infant diarrhoea and child accidental injuries (46). In Butajira, south central Ethiopia, maternal depression when present in addition to intimate partner violence was associated with increased child mortality (47).

Independent of its influence on mental health, intimate partner violence (IPV) is acknowledged by the World Health Organization to be a key social determinant of physical health (48). Women experiencing IPV are 1.5 times more likely to acquire HIV, syphilis, chlamydia and gonorrhoea infection and 16% more likely to have a low birthweight baby. 42% of women subject to intimate partner violence are injured, and 38% of homicides against women are committed by their intimate partner. A recent systematic review has highlighted the impact on birth outcomes with increased risk of premature birth, small for gestational age babies and low birthweight (49). WHO guidance highlights the importance of identifying IPV in maternity settings internationally (50).

Despite the evidence for adverse impacts of maternal mental health problems, alone or in conjunction with intimate partner violence, in the Ethiopian setting, there is currently no service provision at the primary care level (51). As part of the Programme for Improving Mental health carE (PRIME) (52), mental healthcare is being integrated into the general health care settings of primary care and maternal healthcare in Ethiopia. Midwives and primary care clinicians have been trained to deliver mental health care using the World Health Organisation's mental health Gap Action Programme intervention guide (53). However, detection of maternal depression has been extremely low. Through PRIME, a brief psychosocial intervention is being developed and piloted for women with perinatal depression and anxiety.

##### *Interventions for integrated maternal healthcare in Ethiopia*

The Ethiopian Federal Ministry of Health is introducing the Ethiopia Primary Health Care Clinical Guidelines (Ethiopia PHC Guidelines) to promote integrated and evidence-based care in health centres. The Ethiopia PHC guidelines include clinical guidelines on (1) management of obstetric emergencies and (2) routine antenatal and postnatal care, including integrated screening for maternal mental health problems, substance use and experience of violence. The Ethiopia PHC Guidelines will be implemented in the ASSET study districts in early 2018, using the Federal Ministry of Health model for training of Master trainers, facility trainers and delivering on-the-job adult-based learning for health centre-based clinicians.

In ASSET our aim is to support implementation of the Ethiopia PHC guidelines with health system strengthening activities. In the context of these health system strengthening interventions, we will also (1) implement a blood pressure monitoring device (CRADLE Vital Signs Alert) which is able to detect shock and pre-eclampsia with high accuracy, and (2) implement the PRIME psychosocial intervention for common mental health problems and augment with an integrated intervention for intimate partner violence.

#### **Study 2: Diagnostic phase**

##### *Aims*

Following the introduction of the Ethiopia PHCguidelines at health centre level:

(1) Identification of health system bottlenecks for:
    a. the timely detection, care escalation and evidence-based management of life-threatening maternal conditions (pre-eclampsia, sepsis and haemorrhage)
    b. the integration of psychosocial care for perinatal women experiencing mental health problems or exposed to intimate partner violence.
(2) Identifying potential health system strengthening interventions (HSSIs) to address the barriers.
(3) Participatory development of a district-level maternal healthcare plan incorporating HSSIs.

##### *Study 2: Diagnostic phase approach*

The diagnostic phase comprises the following activities: theory of change workshops, situation analysis, antenatal care study, facility process mapping and quality of care study, and a qualitative study.

##### *Study 2: Setting*

The setting for the ASSET integrated maternal healthcare study will be the same as for study 1 (integrated NCDs and mental health care).

#### *Study 2: Theory of Change Workshop*

For the maternal care study, we will conduct Theory of Change workshops at the district level, but also at higher levels in the health system in order to capture the interface with secondary care. The purpose of the workshops, their timing and planned participants are detailed in Table 4.

*Table 4: Theory of Change approach for integrated maternal health care*

| Level | District | Secondary care |
|---|---|---|
| Participants | *Organisational:* District health office heads/focal persons for surgical care (Mareko, Meskan, Sodo), Women's Affairs office representative,<br>*Facilities:* Midwives and health centre-based clinicians (health officers, nurses)<br>*Community:* Community leaders, community representatives from hospital boards, health extension workers, religious leaders, relevant local NGO leads (e.g. Progynists, supporting women who experience intimate partner violence) and beneficiaries/patient representatives, if appropriate. | CEOs/medical directors of Butajira/Buee/Mercy Project hospitals, integrated emergency surgical officers, obstetrician/gynaecologists; midwives, and health officers<br>Zonal health office representatives |
| Timing | Workshop 1: following district situation analysis<br>Workshop 2: following completion of diagnostic phase studies | |
| Purpose | Workshop 1: develop preliminary ToC map to identify gaps in understanding which need to be addressed in the diagnostic phase studies<br>Workshop 2: finalise district level perinatal health care plan, with process indicators and outcomes | |

We estimate that we will include approximately 50 people. Theory of Change workshops need to involve key stakeholders but also need to be manageable in size to allow full participation (not more than 25 persons per workshop).

Inclusion criteria:

- Key stakeholders for the topic under discussion
- Able to converse in Amharic, the official language in the study districts
- Providing informed consent

Exclusion criteria: none.

Informed consent will be sought from the district level Theory of Change workshop participants (See Appendix 5 for information and consent forms) and the workshops will be audio-recorded. The outputs of the Theory of Change workshops will be a detailedroadmap and a report of how key decisions were made.

#### *Study 2: Situation analysis*

<u>*District level situation analysis*</u>

We will collect data available in the public domain to inform our understanding of the current district level situation (community, primary care facility and organisational level) with respect to integrated maternal care. The focus will be on identifying structural barriers to integrated maternal care, e.g. in terms of available personnel, equipment, medications. See Section **Error! Reference source not found.**.

*Secondary care situation analysis*

Information about secondary care level operative interventions will be linked to the surgical study situation analysis using the Ethiopia Hospital Assessment Tool (see Appendix 1). In addition to examining the availability of personnel, equipment and medications to support emergency obstetric intervention, we will examine the structural requirements for optimal non-surgical management of pre-eclampsia/eclampsia, perinatal sepsis and haemorrhage. See study 1 for details of sampling.

#### *Study 2: Facility process mapping and quality of care study*

*Research questions*

(1) What are the processes and bottlenecks for early detection, care escalation and appropriate management for perinatal women with pre-eclampsia, haemorrhage or sepsis?
(2) What is the quality of routine antenatal care in terms of:
    a. Screening for signs of pre-eclampsia and care instituted in response to an abnormal blood pressure reading
    b. Compassionate and respectful care from health workers and consideration of psychosocial aspects of the woman's presentation

*Study design*

Facility-based documentary analysis and observational study.

*Setting*

Six health centres will be selected purposively, one urban and one rural from each of the three districts. The study will also be carried out in the primary/district hospital (Buee hospital) and general/secondary hospital (Butajira).

*Processes of care study*

We will carry out documentary analysis and an observational study with the permission of the health centre/hospital head and with the involvement of health centre/hospital staff working alongside the project workers.

##### Documentary analysis of processes of care

The clinical documentation of aspects of care for pregnant or postnatal women who present with symptoms indicative of pre-eclampsia/eclampsia, sepsis and haemorrhage will be investigated, including initial triage forms, clinical records, delivery records and any additional forms or registrations books that are completed. Using the clinical records, and in discussion with health facility staff, we will map out (1) the type of routinely recorded data available (including the extent it allows for tracking observations in the woman over time e.g. blood pressure), space for documenting domestic violence, (2) the completeness of routinely recorded data, and (3) the pathways through care experienced by these women. For (3), we will develop a proforma to extract data on the following areas:

- What happens when a perinatal woman with danger signs presents to the health facility? (at different times of day/night/weekend/holidays). What triage occurs? How are the danger signs detected? Who detects them? What happens between detection of the danger signs and the initiation of treatment? Where in the health facility is the woman treated? Who is involved in emergency treatment of a perinatal woman? Once treatment is initiated, what monitoring takes place? How and when is a decision to refer made? How is the referral information communicated to the hospital? How and when is a decision to operate made (if indicated)? Who accompanies the woman to the referral institution?

##### Observational study of processes of care

In the hospitals we will conduct an observational study ofprocesses of care for women with danger signs of pre-eclampsia/eclampsia, sepsis or haemorrhage in pregnancy. Two observers will be present in the hospital, one observer with a clinical background and the second observer from the research team. Both will be trained in behavioural observation and in recording the processes that occur in response to the perinatal woman with danger signs. This will include the processes described above, as well as observations of the team interactions and broader contextual factors that appear to be relevant to the woman's care pathway. The observational component will seek to track at least 10 women from presentation at the hospital gates through the care pathway and up to the point of discharge back to the community or referral to a higher-level hospital. The hospital observational study for maternal healthcare will link with the observational study of surgical care processes. See observation checklists in Section **Error! Reference source not found.**.

*<u>Quality of care study</u>*

We will conduct both documentary analysis and an observational study.

##### Documentary analysis of quality of care

We will identify around 20 records from the ANC cross-sectional survey (see below) where there is documentation of abnormal blood pressure in pregnancy. We will then review these charts in detail to extract information on the extent to which the evidence-based care protocols in the Ethiopian PCC guidelines were followed.See section **Error! Reference source not found.**.

##### Observational study of quality of care

The observational study will examine (1) the communication between the healthcare worker and the woman attending for antenatal care, and (2) the extent to which health workers adhere to the Ethiopia PHC guidelines for routine antenatal care.

With the permission of the health worker carrying out antenatal care, project psychiatric nurses or clinically-qualified research workers will complete an observational assessment of the clinician's clinical communications skills (including respectful care) and holistic care of the woman (including the detection and assessment of psychosocial problems). The project workers will use an adapted version of the ENACT scale (Enhancing Assessment of Common Therapeutic factors) (26), which has been adapted for the Ethiopian context and shown to be reliably administered by trained clinicians. We will assess inter-rater reliability of the project workers before proceeding with the study.

The project health workers will also complete an observational checklist developed for the purposes of this study to assess adherence to the Ethiopia PHC guideline for antenatal care and adequacy of assessment in the routine care setting (see Section **Error! Reference source not found.**).

We will purposively select 6 to 8 health centres for the quality of care study, to include both urban and rural facilities. We will aim to assess 5 consultations each for a total of 60 health care workers.

Inclusion criteria:

- Primary care worker providing out-patient care in study clinic.
- Providing informed consent.
- For each observed consultation, patient provides informed consent.

Exclusion criteria: None.

#### *Study 2: Antenatal care study*

*Study design*

Cross-sectional, facility-based survey.

*Research questions*

(1) In women who attend health-centre based antenatal care,
    a. What is their knowledge of what to do about life-threatening pregnancy, childbirth and postnatal complications?
    b. To what extent is antenatal care person-centred in terms of information provision, communication of risk and supported decision-making?
    c. What care do they receive and what is their level of satisfaction?

(2) What % of psychosocial problems (depressive symptoms, substance use and experience of violence) are detected and addressed in women attending antenatal care?

(3) What % of NCDs (hypertension and diabetes) are detected and addressed in women attending antenatal care?

*Sampling and sample size*

Consecutive women attending selected health centres for antenatal care in the health centres in the Meskan districts will be invited to participate.

The sample size calculation is based on the detection of antenatal depression symptoms or violence exposure by health workers in women attending the health centre for antenatal care. To measure 10% detection by health workers with a precision of +/-5%, with alpha = 0.05 and power of 80%, the sample size is given by the formula:

$N = (z^2 \times p\ (1-p))/\delta^2$

$= 1.97 \times (0.1 \times 0.9)/0.0025$

= 139 women with depressive symptoms/violence. Assuming a 20% prevalence of high depressive symptoms and/or exposure to violence in antenatal women (40)(43), a total sample of 695 is required. Accounting for the design effect, the final sample size will be 1 + (n-1)*ICC (where n= number of subjects per cluster) Assuming an intra-cluster correlation of 0.02 and n=100 if the study is conducted in 7 health centres in Meskan district, the design effect will be 1 + (100-1)x0.02 = 2.98 and the total sample will be 2.98 x 695 = 2071.

*Recruitment*

Women who attend for antenatal care at the health centre will be approached by the project data collectors, provided with information about the study and invited to participate. For women who give informed consent, they will participate in the study assessment after they have undergone their antenatal care appointment. The assessment will take place in a private place.

Inclusion criteria:

- Attending the antenatal clinic during the study period
- Able to converse in Amharic
- Providing informed consent

Exclusion criteria:

- Requiring treatment for medical emergency. The health centre clinicians will inform the research staff if the woman is too unwell to speak to the research staff due to a medical emergency.

*Measures*

(1) Data from clinical records

The data collectors will use a structured form (see Section **Error! Reference source not found.**) to extract data from the current antenatal care visit. This will include: parity, gravida, whether the woman meets criteria to be referral, documented mental health or substance use problems and their management, documented blood pressure, documented violence exposure and any intervention.

(2) Interviewer-administered questionnaires

The following fully structured measures will be administered in an interview format.

(1) *Sociodemographic characteristics:* including age, educational level, marital status, place of residence, religion and ethnicity.
(2) Structured questions in relation to extent of person-centred antenatal care, hypothetical actions in response to perinatal danger signs and potential barriers to accessing timely emergency care.
(3) *Depression symptoms and care.* Depression symptoms will be measured using the Patient Health Questionnaire (PHQ-9) (54). This is a nine-item questionnaire asking about the presence of depressive symptoms in the preceding two weeks. Each item is rated according to persistence of the symptom (0 = not at all, 1 = several days; 2 = more than half of the days; 3 = nearly every day). The PHQ-9 has been validated in the study area in both primary care attendees (55) and antenatal care attendees (area under the receiver operating characteristic (ROC) curve 0.91 (95% confidence interval 0.86, 0.96) (56). For primary care attendees a score of five or more was the optimal cut-off for identifying possible depressive disorder, for antenatal women, a cut-off of four or more was optimal. However, at the optimal cut-offs the positive predictive value was less than 50%. In this study, therefore, we will include women who score above the cut-off combined with reported difficulty in their day-to-day activities (measured using the $10^{th}$ item of the PHQ) in order to be a more robust criterion against which to measure the detection of depression by health centre clinicians. Structured questions about the help-seeking for depressive symptoms and acceptability of integrated depression care will also be administered.
(4) *Trauma symptoms.* We will use the Life-Event Checklist (LEC) (57) and Post-Traumatic Stress Disorder checklist for DSM-V (PCL-5) (58) which have been translated into Amharic and are being adapted for a rural Ethiopia context. We will further adapt the questionnaires for the perinatal period.
(5) *Anxiety symptoms.* We will use the PHQ-anxiety measure(59). This has not been used previously in Ethiopia and will undergo translation into Amharic, back-translation and consensus translation, as per WHO recommendations.
(6) *Substance use.* This will be measured using the World Health Organisation ASSIST questionnaire (22). The ASSIST has been used previously in the Ethiopian setting following rigorous translation procedures and adaptation of references for a 'typical drink' (23). The ASSIST allows classification of level of risk for a substance use disorder (low, medium and high) based on the total score. The ASSIST will be used to detect alcohol and khat use disorders. We will ask about changes in substance use in relation to pregnancy.
(7) *Revised Conflict Tactic Questionnaire:* this five-item screening test has been used in other LMIC contexts and found to be a valid measure of intimate partner violence (60). The scale has used previously in the study site and found to have convergent validity with measures of depressive symptoms. We will ask whether the health worker asked about exposure to violence and the acceptability of these questions.
(8) *Health service satisfaction scale (HSSS):* This measure of service satisfaction was adapted from existing scales and validated in the study site for people accessing out-patient mental health care (61). We will

remove the three items that are specific to mental health care and adapt the wording to refer to antenatal care where relevant.

Research clinician-administered assessments:

(1) Blood pressure and fasting blood glucose or HbA1c, as described in Study 1.

*Data collection and quality assurance*

The data collectors will be women (minimum 10th grade completed) who have been trained in administration of the measures using interactive training techniques, including role plays and observation of practice interviews. Data will be collected using electronic tablets. A supervisor will be present in each health centre to oversee data collection, review the questionnaires for incompleteness or inconsistency at the time of data collection and to ensure ethical procedures are followed. We will pay 50 Birr ($2) as time compensation for participants to compensate for their time and any additional expenses incurred by spending extra time at the facility.

*Data management and analysis*

Health worker detection of psychosocial problems (mental health, substance use and violence exposure) will be assessed as follows:

- Mental health problems: health worker documentation of mental health problems in the clinical record compared to scoring above the cut-offs for depressive, anxiety or trauma symptoms.
- Substance use: health worker documentation of substance use in the clinical record compared to positive scores on the FAST or khat use questions.
- Exposure to violence: health worker documentation of violence exposure/problem relationship with partner compared to a positive score on item 5 of the WAST.

*Ethical considerations*

For women who are identified as having severe depression (PHQ-9 score of 10 or above) or who endorse the item indicating suicidal ideation, the information will be given to the antenatal care provider (who has been trained in integrated mental health care) to allow for consideration of the need for clinical intervention. For women who disclose exposure to intimate partner violence during pregnancy, research staff will be trained to listen non-judgementally, offer privacy and confidentiality and information about local agencies who can provide assistance(48), in keeping with World Health Organization guidelines.

##### *Study 2: Qualitative study*

*Research Questions*

In pregnant women with pre-eclampsia, sepsis and haemorrhage:

- What are the community and health system barriers and facilitators to accessing health care in a timely fashion?
- How is the existing care pathway working in practice?
- What are the barriers and facilitators to timely detection and escalation of care along the care pathway?
- What is the acceptability and feasibility of potential health system strengthening interventions? How could these HSSIs be best implemented to optimise care and improve outcomes?

For maternal psychosocial study:

- What affects a pregnant women's decision to disclose emotional difficulties and/or experience of violence in the home to health centre workers?
- What is the acceptability and feasibility of health centre workers asking women about emotional difficulties and experiences of violence?
- What are the perspectives of health workers and pregnant women on the most appropriate interventions for antenatal depression and violence?
- How can interventions to improve detection and management of antenatal common mental disorders and/or violence be integrated into routine practice? What barriers do health workers and pregnant women foresee to uptake and implementation of such interventions?
- What would be health worker and women's priority outcomes for such interventions?

*Study design*

Qualitative study using a framework approach and in-depth interviews with key stakeholders.

*Setting*

The Meskan, Mareko and Sodo districts of the Gurage zone, as described previously.

*Sample*

We will sample a total of around 60 to 70key stakeholders purposively, focusing on the following groups of respondents:

- Family members/the birth companions of women who are identified through the Butajira Health and Demographic Surveillance (HDS) site as having died during pregnancy and were reported as having symptoms of pre-eclampsia, sepsis or haemorrhage in responses to the World Health Organisation Verbal Autopsy questionnaire (63).
- Women who are identified through the Butajira HDS site as having had a child who died in the neonatal period and who reported having symptoms of pre-eclampsia, sepsis or haemorrhage in that pregnancy.
- Women who experience 'near misses' for maternal mortality as defined by the World Health Organisation: "a woman who nearly died but survived a complication that occurred during pregnancy, childbirth or within 42 days of termination of pregnancy", with a particular focus on severe pregnancy-related complications e.g. severe post-partum haemorrhage, severe pre-eclampsia/eclampsia, sepsis or ruptured uterus) (64). The maternal mortality near misses will be identified through health facility and hospital records in the districts. We will also seek to identify maternal mental health 'near misses' for suicide or homicide through health facility or police records.
- Pregnant women who are not attending primary care, identified via health extension workers
- Pregnant women who are attending routine antenatal care
- Health care providers: health extension workers, health centre midwives, health centre clinicians involved in perinatal care, hospital-based midwives and clinicians involved in perinatal care
- District health office leads for maternal health care, Women's affairs office representative, representatives from NGOs engaged in supporting women (e.g. Progynists in Butajira)
- Community leaders, religious leaders, traditional healers and traditional birth attendants.

Sampling will continue until theoretical saturation is attained.

*Inclusion criteria:*

- Able to converse in Amharic
- Aged 18 years of above

- Able to understand the interview.

*Exclusion criteria*

- Not acutely unwell as judged by the qualitative interviewer or reported by the individual.

*Procedures and analysis*

The procedures and analysis for the qualitative study will be the same as those described for study 1.

The location of the interview will be determined according to the convenience of the participant, and may be conducted in the person's home (for pregnant women or their family members), the Butajira mental health research office, health facilities or in the outside area of a centrally-located hotel. If a person is required to travel from their place of residence or work to attend an interview, we will pay them 100 Birr ($4) as compensation.

*Ethical considerations*

Please see the general ethical consideration of the overall ASSET project. For this specific qualitative study, we will follow these general principles, and pay particular attention to the following potential issues. When approaching the families of deceased women or children, great sensitivity will be exercised. In all cases we will take care that people do not feel pressurised to participate. All participants will be remunerated for time and transport costs, where relevant. We will ensure anonymity of the interview transcripts and any quotations used in publications or reports.

#### Study 2: Pilot phase

The output from the diagnostic phase will be the district level plan for integrated maternal healthcare which specifies key system bottlenecks to achieving improved integrated maternal healthcare. The potential health system strengthening interventions (HSSIs) presented earlier will then be adapted to target these key bottlenecks, with new HSSIs developed as needed.

The HSSIs will be piloted in the district hospitals and health centres, with evaluation that includes the following study designs: quality improvement studies, process indicators (pre-post training evaluations, competence evaluations, qualitative studies.

### Study 3: Improving access to quality surgical and dental care

#### Study 3: Background

The global burden of surgical disease is considerable and considered to be one of the most neglected areas in global health. In a recent Lancet Commission, an estimated 32.9% of all lives lost in the year 2010 were estimated to have been due to health conditions needing surgical care (1), resulting from a 90% treatment gap for basic surgical care in lower-income countries. Inequity is high, with the biggest unmet need for surgical care being borne by the poorest sector of society. The Lancet Commission identified indicators and targets (to be achieved by 2030) for surgical care in all countries. These are summarised in Table 5.

*Table 5: Lancet Commission on Global Surgery Indicators and Targets for 2013*

| **Indicator** | **Target by 2030** |
|---|---|
| % of population able to access, within 2 hours, a facility that can do: Caesarean Section, emergency laparotomy and treatment of open fracture (Bellwether Procedures) | 80% coverage of essential surgical and anaesthetic care |
| Number of surgical and anaesthesia specialists per 100,000 population | At least 20 surgical, anaesthetic, and obstetric physicians per 100 000 population |
| Surgical volume: number of surgical procedures carried out per 100,000 population | 5000 surgical procedures/100,000 population |
| Perioperative mortality: all-cause mortality before discharge from hospital per number of people having an operative procedure | Perioperative mortality tracked in 100% of facilities |
| Protection against impoverishing healthcare expenditure due to need for surgical care | 100% protection against impoverishment from out-of-pocket payments for surgical and anaesthesia care by 2030 |
| Protection against catastrophic healthcare expenditure due to need for surgical care | 100% protection against catastrophic expenditure from out-of-pocket payments for surgical and anaesthesia care by 2030 |

Disorders that could be managed by surgery constitute a significant portion of the global burden of disease[1]. Injuries kill nearly 5 million people and about 270000 women die from complications of pregnancy annually. Many of these injury-related and obstetric-related deaths, as well as deaths from other causes (e.g. abdominal emergencies and congenital anomalies), could be prevented by improved access to surgical care. Beyond mortality, untreated surgical diseases are among the top 15 causes of physical disability worldwide. More than 11% of the world's burden of disease stems from conditions that can be treated successfully with surgery. In the case of disability, surgery is not merely curative, but also prevents the social and economic disparities that accompany untreated disabilities. Like other public health solutions, surgery improves the economic output due to improved population health.

Access to surgical care in LMICs has been long neglected and poorly understood in the global public health framework. Recent estimates have shown that there are as many as 5 billion people with little access to essential surgical care. As a result, common birth defects and minor injuries become life-threatening or result in life long disabilities, and easily treatable conditions cause death. The neglected person with a surgical condition is one who suffers from the lifelong consequences of inadequate treatment and management of common conditions such as obstructed labor, cataracts, hernias, cancers, injuries, and untreated congenital and infectious diseases.

Despite this large burden, surgical services are not being delivered to many of the individuals who need them most. An estimated 2 billion people lack access to even the most basic of surgical care. This need has not been widely acknowledged, and priorities for investment in health systems' surgical capacities have therefore only recently been investigated. Indeed, until the 1990s, health policy in resource-constrained settings focused sharply on infectious diseases and under-nutrition, especially in children. Surgical capacity was developing in urban areas but was often viewed as a secondary priority that mainly served socioeconomically advantaged people.

The need to improve the quality of surgical care in Ethiopia is not disputed, but there may be challenges in implementing quality improvement initiatives in this setting (2). In particular, in an Ethiopian study, the use of clinical audit was perceived as a process for apportioning blame rather than resolving clinical management problems. The lack of a tradition of multi-disciplinary working, strong hierarchies and high turnover of staff undermined efforts to achieve quality improvement in surgical care.

The current professional profile of Ethiopia showed that there are approximately 250 Surgeons, 300 Gynecologists, 50 orthopedic surgeons, and 100 ophthalmologists in Ethiopia. The number of surgeries done in Ethiopia is not more than 200000 a year with the unmet need of 5000000 surgeries per year. The waiting time for surgery extends up to four years, especially in referral hospitals. Surgical safety is a concern as most institutions do not implement the standard WHO surgical check list(65)(66).

Ethiopia has emerged as a leader among low-income countries working to improve access to surgical and anaesthesia care for its population of 100 million people. Recognizing Ethiopia's limitations in providing safe and essential surgery, the Federal Ministry of Health (FMOH) launched the Saving Lives through Safe Surgery (SaLTS) initiative in 2015. SaLTS is a five-year (2015-2020) strategy that aims to improve access to safe surgical and anaesthesia care across all levels of the health system. The initiative is designed to address quality and equitable health system reform, one of four transformation agendas of the FMOH's national Health Sector Transformation Plan (2015-2020). SaLTS is the one of the first national surgical plans written and is by far the most expansive implementation of a national surgical reform program in both sub-Saharan Africa and any low-income country. SaLTS uses a forward-thinking health systems approach through the Ethiopian Hospitals Alliance for Quality (EHAQ) platform to simultaneously tackle activities across eight 'pillars of excellence' and avoids the pitfalls of vertical programming.

#### Study 3: Diagnostic phase

The goal of the diagnostic phase is to develop a surgical and dental health care plan at the level of the district and hospital quality cluster which will lead to expanded access to quality essential and emergency care.

The Ethiopian Health Alliance for Quality established clusters of hospitals in an effort to allow better-resourced hospitals to support less well-resourced hospitals and improve the access to quality surgical care. In discussion with the Regional Health Bureau representative for SaLTS at the Southern Nations Nationalities and Peoples' region (SNNPR), ASSET will work with a specific quality cluster led by Butajira hospital (a general hospital), co-led by Wurabe hospital (general hospital), and including Durame (general hospital), Buee (primary hospital), Tora (primary hospital), Kibet (primary hospital), Gunchire (primary hospital) and Saja (primary hospital). Not all interventions will be implemented in all hospitals, and more in-depth evaluation of the impact of evaluation will be reserved for Butajira and Buee hospitals (both in the Gurage Zone).

Sub-studies in the diagnostic phase for surgical care will comprise Theory of Change workshops with key stakeholders, a situation analysis, facility studies, a community survey, and a qualitative study. Each sub-study is described in detail below.

#### *Study 3: Theory of Change workshops*

The ToC approach has been described above. For the surgical care study, we will conduct ToC workshops at the national/regional and quality cluster/district levels. The purpose of each workshop and planned participants are described below inTable 6.

*Table 6: Participants for Theory of Change workshops*

| **Level** | **National/regional** | **Quality cluster/district** |
|---|---|---|
| **Participants** | Federal Ministry of Health: Director of the quality directorate, SaLTS team, clinical services directorate, Surgical Society of Ethiopia, Ethiopian Anesthesiologists' Association, Anesthesia Professionals Association.<br>Regional Health Bureau: SaLTS lead, quality lead.<br>CEOs/medical directors and salts focal persons of all quality cluster hospitals<br>Representatives from other surgery improvement initiatives in Ethiopia; relevant NGO leads; patient organisation representatives | Zonal health office head/focal for surgical care, woreda health office heads/focal persons for surgical care (Mareko, Meskan, Sodo, Silti), CEOs/medical directors of Butajira/Buee/Mercy Project hospitals, surgeons, integrated emergency surgical officers, obstetrician/gynaecologists, anaesthetists/anaesthetic assistants, recovery room nurses, dentists, dental assistants, community leaders, community representatives from hospital boards, religious leaders, relevant local NGO leads and patient representatives.<br><br>Also, key individuals from the national/ regional ToC to facilitate cross-over learning. |
| **Timing** | Workshop 1: after rapid situation analysis completed<br>Workshop 2: at the end of the diagnostic phase | |
| **Purpose** | Both levels of workshop (national/regional and cluster/district) will be focusing on elucidating the roadmap to improved access to quality surgical, anaesthetic and dental care at the cluster/district level, within the framework of the national SaLTS programme.<br>**Workshop 1:** develop preliminary ToC map to identify gaps in understanding which need to be addressed in the diagnostic phase studies.<br>**Workshop 2:** finalise district/quality cluster level surgical, anaesthesia and dental health care plan, with process indicators and outcomes. | |

We estimate that we will include approximately 50 people. Theory of Change workshops need to involve key stakeholders but also need to be manageable in size to allow full participation (not more than 25 persons per workshop).

Inclusion criteria:

- Key stakeholders for the topic under discussion
- Able to converse in Amharic, the official language in the study districts
- Providing informed consent

Exclusion criteria: none.

Informed consent will be sought from district/quality cluster level ToC participants (See Appendix 5 for information and consent forms) and the workshops will be audio-recorded. The outputs of the ToC workshops will be a full ToC map and a report of how key decisions were made in order to agree on the ToC map.

#### *Study 3: Situation Analysis*

A situation analysis will be undertaken in order to provide a baseline understanding of (1) the potential structural bottlenecks to improving access to high quality surgical, anaesthetic and dental care, and (2) the adequacy of routinely available information about surgical, anaesthetic and dental care.

The situation analysis will collate data using the standardised and nationally endorsed Hospital Assessment Tool and the ASSET situation analysis tool.

##### *Hospital Assessment Tool*

The Ethiopian Federal Ministry of Health, in collaboration with the World Health Organisation and the Programme in Global Surgery and Social Change (PGSSC) has developed a Hospital Assessment Tool to indicate the preparedness of facilities to deliver surgical care. The Hospital Assessment Tool is administered to the hospital CEO, surgeon/integrated emergency surgical officers (IESOs), obstetrician/ gynaecologist and anaesthetist/operating theatre nurse in order to collect data about physical structures, staffing, information systems and documentation, equipment and medication, volume and type of surgical procedures, costs and surgical outcomes and complications. See Section **Error! Reference source not found.**. The Hospital Assessment Tool has been modified for ASSET to also assess dental care.

With permission from the Regional Health Bureau, we will administer the Hospital Assessment Tool to all eight hospitals in the quality cluster described above. The data will be collected by trained surgical nurses. The data will be analysed descriptively and presented in a report form to be shared with the Regional Health Bureau and hospital CEOs. For ASSET we will circulate the report to participants in the ToC workshops to inform discussions about structural bottlenecks to be overcome to improve surgical and dental care. We anticipate consulting with around 25 hospital workers across the eight hospitals in order to complete the hospital assessment tool. These workers will be selected on the basis of their key role in the delivery and management of surgical and dental care services, with permission obtained from the hospital CEO.

Inclusion criteria

- Professional engaged in part of the surgical pathway
- Providing informed consent

Exclusion criteria: none.

##### *District level situation analysis*

To complement the hospital level situation analysis, we will also collect data available in the public domain to inform our understanding of the current district level situation (community, primary care facility and organisational level) with respect to the surgical care pathway.

#### *Study 3: Facility process mapping study*

##### *Research questions*

At the hospital level, what are the processes from the time a person presents with a surgical condition until they are discharged from the hospital?

What processes of surgical care are documented and to what extent is information acted upon appropriately?

*Study design*

We will conduct a facility-based documentary analysis and observational study.

*Setting*

The study will be conducted in Butajira general hospital and Buei primary hospital.

*Documentary analysis procedures*

We will carry out the study in collaboration with the hospital management, with the assistance of designated clinicians.

The clinical documentation of aspects of care for a person presenting with a surgical condition will be investigated, including initial triage forms, clinical records, nursing records (if separate), surgical checklists and any additional forms or registrations books that are completed. The documentation will be identified from all relevant clinical settings; for example, triage room, emergency department, obstetrics ward, surgical ward, operating theatre.

We will map out (1) the type of routinely recorded data, and (2) the completeness of routinely recorded data. The completeness of data recording will be investigated on the clinical documents linked to around 20 patients who underwent a surgical procedure or who presented to hospital with a condition requiring a surgical intervention.

*Observational study*

We will work with the hospital management to identify clinical staff who will work with research staff to map out and observe the processes of surgical care, from presentation to the hospital until discharge. This will be carried out over a one-month period, including observation on weekend days. In addition to indicators of surgical quality, ethnographic observations will explore the social dynamics, styles of communication and organizational norms involved in the delivery of care. Examples of the kinds of processes, exchanges and indicators of interest are presented in Table 7.

*Table 7: Processes and indicators on surgical care pathway*

| | |
|---|---|
| Hospital main gate | Any restrictions or delays on entering the hospital compound?<br>Where do ambulances get sent? |
| Triage room/ emergency department | Are there any delays between presentation to hospital and initial triage? What are the reasons for delays (e.g. needing to purchase a card, presenting during lunchtime, etc).<br>Who carries out the triage? Do they measure vital signs? Are they documented? What happens next if the person is considered to have a surgical condition? To whom and how is this information communicated? |
| Surgical assessment | At what stage would the patient be reviewed by a surgical health worker (health officer level, general practitioner, surgeon or obstetrician/ gynaecologist)? Who else is involved in the diagnostic process (patient, family members?) and what information is provided by the patient that might facilitate referral? E.g. about connections or payments.What are the reasons for delays before surgical review? When the decision to operate is made, what happens next? What observations are conducted while the person is waiting for surgery? What else needs to happen before the surgery can take place? |

| | |
|---|---|
| | What happens if the person needs surgical referral? How long do they wait in the hospital before transfer? |
| Operating theatre | Observe the use (or not) of the WHO safe surgery checklist, and the adequacy of adherence.<br>Apply the Cleancut checklist (also pre- and post-operative): process of skin antisepsis, availability and use of antiseptic agents, how hands are washed and scrubbed, how gowns and drapes are washed, dried and sterilised, how gowns and drapes are mended, how sterile gloves are procured, how sterilisers work and are maintained, how instruments are cleaned, how instruments are sterilised, how sterilisation is confirmed, what antibiotics are available, how the antibiotics are obtained, how and when antibiotics are given, who administers antibiotics, how long are antibiotics continued, protocols that exist for peri- and post-operative administration of antibiotics, how swabs are tracked during surgery, how often swabs are counted, what happens when there is a discrepancy in swab numbers, how surgical wound infections are documented, how is information on wound infection collected, how surgeons can know their wound infection rates, where patients go if they develop wound infection after hospital discharge.<br>What are the interactions between staff involved implementing the checklist? Who is in charge and if steps are missed how these corrected? What is the layout of the theatre? What is the spatial distribution of experts and equipment? |
| Post-operative recovery room | What instructions are communicated and in what manner (including tone) to the recovery room staff about observation level, warning signs? How frequently are observations carried out? What happens if the patient deteriorates? |
| Post-operative surgical ward | What instructions are communicated and in what manner to the post-op surgical ward staff about observation level, warning signs? How frequently are observations carried out? What happens if the patient deteriorates? |

Inclusion criteria

- Consecutive emergency presentations

Exclusion criteria: none.

*Data analysis*

We will collate the information obtained from documentary and observational sources and triangulated with relevant qualitative data (see qualitative study below) to identify key system bottlenecks. This information will (1) be fed back to the hospital management, (2) used to inform the theory of change map and identification of interventions to improve access to quality surgical care, and (3) used to design research tools for capturing process data, to be used in the piloting and implementation phases of the project.

#### *Study 3: Surgical cohort study*

*Research Questions*

In people who attended Butajira or Buee hospitals and who had a surgical condition needing intervention:

1. What percentage experienced peri- or post-operative complications?
2. What was their level of satisfaction with the care received and what delays did they encounter trying to access care?
3. What was their post-operative level of day-to-day functioning?
4. What costs did they incur when accessing surgical care and in what % of people was this cost catastrophic (>10% of annual expenditure) or impoverishing (pushing household below poverty threshold)?

*Study design*

The study will be a hospital-based cohort study, with adults recruited at the time that they are identified as having a condition requiring surgical intervention and with a follow-up assessment conducted at 30 to 60 days post-operatively in the community.

*Setting*

Butajira and Buee hospitals, Gurage zone.

*Sampling*

We will approach consecutive adults who are identified as having a condition requiring surgical intervention in the study hospitals over a four-month period. On the basis of existing levels of patient flow, we expect the sample size to be approximately 150 people in total (approximately38/month) from Butajira hospital.

*Inclusion criteria*

- Aged 18 years and above
- Able to converse in Amharic
- Able to understand the interview (for example, excluding people with severe intellectual disability or dementia)
- Providing informed consent

*Exclusion criteria*

- Medically too unstable to discuss with research staff.

*Study procedures*

In discussion with the surgical teams in the hospital, and building on the facility observational study, improvements will be made to the paper-and-pen capture of existing indicators relevant to quality surgical care. Data capture will also be supported by project staff during the sub-study to ensure accurate information. The project staff will attend the pre- and post-operative wards on a daily basis to extract relevant information from the clinical records.

Pre-discharge, potential participants will be approached, informed about the study and invited to participate. For patients who are discharged without being approached, we will use contact information in hospital records, cross-checked with the Butajira Health and Demographic Surveillance Site (HDSS) database for people in Meskan/Mareko. The Butajira HDSS enumerators will introduce data collectors to the potential participants, who will then be given an information sheet/ the information sheet will be read out and the person invited to consent to participation. If the person who underwent the surgical operation is no longer alive, the nearest relative or caregiver will be invited to complete the relevant components of the interview.

Face-to-face interviews will be conducted between 30 and 60 days post-discharge from hospital in the person's home or at a location convenient to them.

##### *Measures*

###### Hospital records

Project data collectors will extract the following information from the enhanced clinical records/ project forms at the time of discharge from hospital: indication for operation, type of operation, time from admission to operation, peri-operative complications, post-operative complications (including anaesthesia-related complications, surgical site infections, haemorrhage, re-operation and mortality), length of stay in hospital, planned vs. self-discharge, cost of hospital stay.

###### Community-based interviews

*Sociodemographic characteristics*

Structured questionnaire administered as an interview asking about age, gender, educational level, place of residence and marital status.

*In-Patient Assessment of Health Care (I-PAHC) questionnaire*

We will use 21 items from the I-PAHC questionnaire, which was developed and validated in Ethiopia (67). The participant will be asked to rate their experience of being admitted to hospital for surgical care.

*World Health Organisation Disability Assessment Schedule (WHODAS version 2.0; 12-item)*

The WHODAS is a 12-item scale measuring functional impairment caused by any health condition(24). The WHODAS includes items covering work, interpersonal and social functioning, as well as the number of days out of role. The WHODAS has been used in Ethiopia to measure disability in people with depression (68) and severe mental disorders and found to be valid (25). The reference period for the WHODAS is the past one month.

*Help-seeking*

This is adapted from the Butajira Treatment Questionnaire (BTQ). The BTQ asks participants where they first sought help, their subsequent pathway to care, the time from developing symptoms until receiving surgical intervention and reasons for delay.

*Household economic impact of surgical care*

This will be adapted from a household survey instrument used previously in the WHO SAGE study on health and ageing in six LMICs(69), using the sections on assessing household size, consumption and strategies employed to cope with the costs of accessing surgical care. The SAGE questionnaire has been used in the study site previously, contextualised for relevance, translated into Amharic and found to be acceptable and feasible.

#### *Data analysis*

We will describe the help-seeking and potential barriers for timely surgical care in those who access care, level of satisfaction with care and post-operative functioning using simple descriptive statistics. We will estimate the extent to which the person restored their previous level of functioning by comparing the number of days able to function in a month.

Catastrophic healthcare costs will be calculated by summing all expenditure and presenting the expenditure on surgical care as a % of total household expenditure. In order to compare the level of catastrophic healthcare costs with that experienced by households from population controls, we will use estimates from our previous Emerald study.

#### *Study 3: Community survey*

*Research Questions*

In a rural community sample:

- What is the prevalence of surgical and dental conditions, and the unmet need for surgical and dental care, in a predominantly rural Ethiopian community?
- What is the burden of life-limiting conditions in the community?
- What percentage of deaths were potential avertable with surgical intervention?
- What are the preferences expressed for different types and levels of disability to inform development of utility weights?

In people with a surgical or potentially life-limiting condition:

- What is the difference in level of impairment, pain, overall functioning and days out of role in those receiving a surgical intervention?
- What is the difference in total health care costs in the last two months in people who received a surgical intervention compared to those who did not?
- What are the explanatory models and help-seeking behaviours for surgical, dental and potentially life-limiting conditions?

*Setting*

The study will be conducted in the Butajira Health and Demographic Surveillance Site (HDSS).

*Pre-piloting and development work*

For the investigation of utility weights for disability items, we will do exploratory work with focus groups and cognitive interviewing in the target group (members of the general population in a rural Ethiopian setting). This exploratory work will examine the type of approach to determining utility weights which is feasible in this setting.

*Sampling and sample size*

Sample A (avertable mortality due to surgical conditions): All households with a death recorded in the Butajira HDS database in the preceding four to five years will be included. There are an estimated 180 to 200 deaths per year in the HDS population (Dr Mitike Molla, personal communication). We will aim for a sample size of 700 which will give precision of +/-1.4% around the percentage of deaths which could potentially have been averted with access to surgical care, assuming that 25% of deaths were potentially avertable (4).

Sample B (prevalence of surgical conditions needing intervention): In order to detect a prevalence of 25% of unmet need for surgical care in the community (4), a sample size of 600 households (with 2 persons sample randomly per household, n=1200 in total) is required to achieve a precision of +/-2.5%. We will stratify sampling by sub-district (kebele) and select households randomly, with probability proportionate to size of the total population of the kebele. Within households, we will use random sampling to select two people to be interviewed using the HDSS sampling frame. If the selected household member is not present, we will return to the home on a convenient date up to three times. If the person is still not present, we will go to the next randomly selected household.

*Inclusion criteria for Sample B:*

- Living in the Butajira HDSS for at least one year prior to the survey.
- Able to communicate in Amharic, the working language of the study site.
- For adults (18 years and older), informed consent will be required.
- For persons aged 16 or 17 years and are married, consent will be obtained from the individual.
- For children aged 13 to 17 years, assent and parental/guardian consent will be required.
- For children aged 12 years or younger, consent will be obtained from a parent or guardian and non-refusal from the child.

*Exclusion criteria*

- Medical emergency identified

*Measures*

- In order to investigate deaths that were potentially avertable with a surgical intervention, we will request access to the Verbal Autopsy data collected by the Butajira Health and Demographic Surveillance Site. If the Verbal Autopsy data are not available, we will utilise Section 1 of the SOSAS, as described below.

- The **Surgeons OverSeas Assessment of Surgical Need (SOSAS)** questionnaire has been used in settings similar to Ethiopia (in Sierra Leone, Nepal and Rwanda (70)(71) and validated against surgical assessment in Nepal(72).

The SOSAS is a structured questionnaire which has two sections.

- Section 1 will be administered to all households where a death occurred in the previous year (sample A). The interviewer will establish whether the person had any of the following conditions in the week before their death: abdominal distension or pain; bleeding or illness during childbirth; injury; mass, growth, or swelling; acquired deformity; or a wound not due to injury or congenital deformity. The measure also asks about help-seeking and delays in obtaining care prior to death.

  For deaths occurring in women who were pregnant, in childbirth or in the post-partum period, additional questions will be asked (see Maternal Health Study).

- Section 2 will be administered to two household members in Sample B. The SOSAS questionnaire is a head-to-toe verbal examination of each anatomical region (face, head and neck; chest and breast; abdomen; groin, genitals and buttocks; back; arms, hands, legs and feet), seeking to identify conditions requiring a surgical procedure (wound care, suturing, incision, excision, manipulation of tissue in a safe and painless way). The original SOSAS will be adapted to be administered by surgical residents (rather than lay interviewers) and to be (1) assess surgical conditions experienced in the past 12 months, and (2) assess surgical conditions which are more longstanding but associated with residual difficulties. The surgical residents will be conduct a physical examination as well as verbal investigation, chaperoned by a family member or lay data collector of the same gender. The surgical residents will make the judgement of whether or not there is unmet need for surgical care, which will be compared to the participant's perceptions of needs for care.

If a surgical condition is identified, the person will be given a referral slip to the nearest local provider (and communication of the referral facilitated). If the condition is judged to be urgent, the project will facilitate transport to the facility.

- Where a surgical condition is identified, the lay data collector will then administer a fully structured questionnaire to investigate help-seeking behavior, delays/barriers to obtaining surgical care and costs of help-seeking and having a surgical condition. We will use sections from the **Client Service Receipt Inventory (CSRI)** to examine costs associated with the surgical condition. The CSRI has been adapted for use in the Butajira area previously (68).

*Data collectors*

The fully structured questionnaires will be administered by lay data collectors who will be trained for 5 days in the administration of the questionnaires and ethical conduct.

To examine utility weights for different disability states, we will randomly administer items from the WHODAS, 12-item version (see above) linked to questions about the person's preferences for different states.

*Data collection procedures*

The surgical assessment will take place in the homes of participants. The selected households will be identified with support from Butajira HDSS enumerators. The interview and physical examination will be conducted in a private place.

The lay data collectors will be supervised in the field by supervisors. Non-identifiable data will be collected using electronic tablets operating offline. The data will be uploaded on a daily basis when internet access is available and kept securely. Electronic back-ups of the data will be stored on password-protected project computers. Identifiable data about the person, for example their name and the name of the household head, will be collected on paper forms which link the person to the project identification number. These forms will be kept securely in a separate place to the questionnaire data. The data will be entered into a separate, password-protected database.

*Specific ethical considerations*

Any person identified as having a surgical condition needing urgent intervention will be given an explanation about the need for treatment and referred to Butajira general hospital, with transport arranged by the project. Persons identified to have surgical conditions requiring non-urgent intervention will be given a referral to the appropriate service. The project will undertake all efforts to facilitate the referral and care of these persons.

Respondents will not be paid for their participation in the study.

#### *Study 3: Qualitative study*

*Research questions*

- What are the barriers and facilitators to improving access to quality surgical care, and what are the patterns of help-seeking?
- What are the unmet needs of people with surgical conditions who do, and do not, access surgical care? What is the impact of unmet needs for surgical care?
- What are the care pathways, costs and processes and how do these vary by provider?

- How might surgical care systems be strengthened to increase contact coverage (surgical volume) and improve processes and outcomes of care?

*Sample*

A purposive sample of stakeholders will be selected, as described in Table 3. We estimate that a total of 40 to 50 interviews will be conducted, based on when theoretical saturation is attained.

*Table 3: Sampling approach for surgical qualitative study*

| | |
|---|---|
| People with surgical conditions who have not accessed care | Identify from community survey |
| People with surgical conditions who have accessed care | Identify from facility study |
| Surgical care providers (surgeons, IESOs, obstetrician/gynaecologists, GPs providing surgical care, surgical/OT nurses, anaesthetic providers) | Butajira and Buee hospitals |
| Managerial level: woreda heads, hospital CEOs, quality cluster leads | Butajira and Buee hospitals, Meskan, Mareko and Sodo districts |
| Primary health care providers (health centre staff and HEWs) | Meskan, Mareko and Sodo districts |
| Traditional and religious healing (bone-setters, holy water, tooth extracters etc) | Meskan, Mareko and Sodo districts |
| National level stakeholder for surgical care | Federal Ministry of Health and regional health bureau leads for SaLTs programme. |

*Study design*

Qualitative study using a framework approach and in-depth interviews with key stakeholders.

*Setting*

The Meskan, Mareko and Sodo districts of the Gurage zone, as described previously.

*Eligibility criteria:*

Able to converse in Amharic, aged 16 years of above, not acutely unwell and requiring emergency care, and able to understand the interview.

*Data collection and analysis*

We will use the same methodology as described for study 1. The topic guides are provided in Appendix 3, but will be developed iteratively as data collection proceeds. We will pay 100 Birr ($4) as time compensation for any participant who is required to travel from their place of work or residence in order to participate in the interview.

*Ethical considerations*

Please see the general ethical consideration of the overall ASSET project. For this specific qualitative study, we will follow these general principles, and pay particular attention to the following potential issues. When approaching the families of deceased persons, great sensitivity will be exercised. In all cases we will take care that people do not feel pressurised to participate. All participants will be remunerated for time and transport costs, where relevant. We will ensure anonymity of the interview transcripts and any quotations used in publications or reports.

### Study 3: Pilot phase

The output from the diagnostic phase will be the district level plan for quality surgical and dental care which specifies key system bottlenecks. The potential health system strengthening interventions (HSSIs) presented earlier will then be adapted to target these key bottlenecks, with new HSSIs developed as needed.

The HSSIs will be piloted in the quality cluster district hospitals and health centres, with evaluation that includes the following study designs: quality improvement studies, process indicators (pre-post training evaluations, competence evaluations, qualitative studies.

### Ethical considerations for ASSET studies

Ethical approval will be obtained from the Institutional Review Board of the College of Health Sciences, Addis Ababa University, Ethiopia, and the Psychiatry, Nursing & Midwifery subcommittee of King's College London's College Research Ethics Committee, UK.

Specific ethical considerations are presented in relation to each of the individual ASSET studies. Here we present the overall ethical considerations which are relevant across the studies.

**Informed consent**

All participants will be informed about the study and given the opportunity to ask questions before being invited to consent to participate. Only people who give voluntary informed consent will be included in the study. We will seek written consent, but if the person is not literate, we will ask a literate witness to sign to confirm that the information sheet has been read out correctly. In such cases we will ask the person to give a thumb print to signify their consent to participate.

**Data protection**

All paper-based records collected in this study will be stored in a secure filing cabinet or cupboard. All electronic data resulting from the project will be stored on a secure computer network and/or password-protected computer and external hard drive.All data will be encoded so that the interview and test results cannot be directly linked to contact details. Data relating to personal information of the participants (e.g. their names and contact details) will not be stored on portable devices or media, but only in highly secured systems, and the paper-based records (e.g. the signed informed consent forms) will be stored securely and separate from the questionnaire and interview data. Only members of the research group will have access to the data. Audio files will be encrypted and password-protected and only identifiable by the participant's project identification number (making sure that the names of the participants are not included in the recording).

Personal data collected for the study will be stored until 2 years after the study is completed. Other research data will be stored until 7 years of completion of the study.

**Recompense to participants**

Where appropriate, we will remunerate participants for their time and reimburse their travel costs.

**Deception**

No deception is involved in this study. The nature of the research is explained in the information sheets (see Annex).

**Risk of harm to participants**

We will minimise the risk of harm by ensuring that participants are fully informed about the study prior to voluntary participation. In the event that a participant wishes to withdraw from taking part in ASSET or from contributing their data, they are free to do so.

We recognise that some participants could possibly become distressed when interviewed about their health condition, mental health or substance use problems, or experiences of violence. We will seek to minimise this occurring in the first place by ensuring that questions are worded sensitively. We will ensure that our data collectors are trained to be sensitive to signs of distress and to offer to reschedule or discontinue the interview if distress is noted. If a person remains distressed, the data collector will contact the supervisor and make arrangements for appropriate support to be provided. If needed, referral will be made to local health workers who have been trained in mental health care.

**Benefits**

The contributions of participants will help to improve the healthcare delivered in the study districts.

The study will also build capacity in health workers and healthcare administrators in the study districts.

The findings of this study are likely to apply to other low-income African settings too.

**Confidentiality**

Confidentiality will be maintained throughout. Interviews will be conducted in private. Confidentiality of collected data will be maintained as described in the Methods and Data Protection sections.

**Declaration of conflicts of interest**

None of the collaborators have any potential conflicts of interest with the proposed project.

**Quality assurance**

The project PIs (CH, AB, SS and AA) will visit the study site on a regular basis in order to ensure that the ethical principles outlined above are adhered to, including random checks of the procedure for obtaining informed consent. This will be supported by our established procedures for ethical data management (see Methods and Data protection sections).

### Timetable for diagnostic and pilot phase studies

| **Project activities** | **2018** | | | | | | **2019** | | | | | | |
|---|---|---|---|---|---|---|---|---|---|---|---|---|---|
| | **Jul** | **Aug** | **Sep** | **Oct** | **Nov** | **Dec** | **Jan** | **Feb** | **Mar** | **Apr** | **May** | **Jun** | **Jul** |
| Training of regional and district trainers | ■ | ■ | | | | | | | | | | | |
| Training of facility staff in study districts | ■ | ■ | ■ | ■ | ■ | ■ | | | | | | | |
| Theory of Change workshops: surgical care | | ■ | | ■ | | | | | | | | | |
| ToC workshops: maternal & NCD/mental health care | | | | ■ | ■ | | | | ■ | | | | |
| Hospital assessment tool | | ■ | | | | | | | | | | | |
| Hospital process mapping study | | | ■ | ■ | | | | | | | | | |
| Health centre process mapping/ quality of care | | | | | | ■ | | | | | | | |
| OPD patient survey | | | | | ■ | ■ | ■ | ■ | | | | | |
| OPD observational study | | | | | ■ | ■ | ■ | ■ | | | | | |
| ANC patient survey | | | | ■ | ■ | ■ | ■ | | | | | | |
| ANC observational study | | | | ■ | ■ | ■ | ■ | | | | | | |
| Community survey: surgical burden | | | ■ | ■ | ■ | | | | | | | | |

| |
|---|
| Surgical cohort |
| Qualitative study |
| Development of system strengthening interventions |
| Piloting and process data/qualitative evaluation |

### Community engagement and dissemination plans

We have established community advisory boards in the three study districts as part of existing projects (PRIME and AFFIRM). The community advisory boards include representatives from the woreda health office and other relevant woreda offices, service users, health centre heads, religious leaders, NGO leads and community leaders. The community advisory boards will be continued for the ASSET project. We will meet with the community advisory boards on a regular basis in order to consult about the project plans and activities and to feedback findings from the study.

Members of the research team have active partnerships with the Ministry of Health. Dissemination of study findings through these partnerships will take a high priority. We will support this with production of policy briefs. We will publish our work in peer-reviewed journals and present the findings at local and international conferences. We will build on our previous mapping of stakeholders and ensure timely dissemination of findings.

### Budget

| Summary Totals | Funding per year of project (Ethiopian Birr) | | | | Total funding (Ethiopian Birr) |
|---|---|---|---|---|---|
| | 2018 | 2019 | 2020 | 2021 | |
| Directly Incurred Costs: | | | | | |
| Personnel | 3192500 | 4673000 | 4490000 | 631000 | 12986500 |
| Equipment | 1934000 | 225000 | 0 | 0 | 2159000 |
| Travel | 1701500 | 2114000 | 2044000 | 433500 | 6293000 |
| Consumables | 506200 | 520400 | 475400 | 21600 | 1523600 |
| Fieldwork | 6477500 | 4892400 | 1474100 | 21300 | 12865300 |
| Indirect Costs | 1104936 | 993984 | 678680 | 88592 | 2866192 |
| Total | 14916636 | 13418784 | 9162180 | 1195992 | 38693592 |

### References

1. The Federal Democratic Republic of Ethiopia Ministry of Health. Envisioning Ethiopia's path towards universal health coverage through strengthening primary health care. Addis Ababa, Ethiopia: FMOH; 2015.

11. World Health Organization and Calouste Gulbenkian Foundation. Integrating the response to mental disorders and other chronic diseases in health care systems. Geneva, Switzerland: World Health Organization; 2014.

29. Central Statistical Agency [Ethiopia], ICF. Ethiopia Demographic and Health Survey 2016: Key Indicators Report. Addis Ababa, Ethiopia, and Rockville, Maryland, USA.: CSA and ICF.; 2016.

50. World Health Organization. Responding to intimate partner violence and sexual violence against women. WHO clinical and policy guidelines. Geneva: WHO; 2013.

51. Baron EC, Hanlon C, Mall S, Honikman S, Breuer E, Kathree T, et al. Maternal mental health in primary care in five low- and middle-income countries: a situational analysis. BMC Health Serv Res. 2016;16:53 doi:10.1186/s12913-016-1291-z.

52. Fekadu A, Hanlon C, Medhin G, Alem A, Selamu M, Giorgis TW, et al. Development of a scalable mental healthcare plan for a rural district in Ethiopia. Br J Psychiatry J Ment Sci. 2016 Jan;208 Suppl 56:s4-12.

53. World Health Organization. mhGAP Intervention Guide for mental, neurological disorders in non-specialised health settings, version 2.0. Geneva, Switzerland: WHO; 2016.

54. Kroenke K, Spitzer RL. The PHQ-9: A new depression diagnostic and severity measure. Psychiatr Ann. 2002;32:509–15.

55. Hanlon C, Medhin G, Selamu M, Breuer E, Worku B, Hailemariam M, et al. Validity of brief screening questionnaires to detect depression in primary care in Ethiopia. J Affect Disord. 2015;186:32–9.

56. Girma F. Detecting depression during pregnancy: validation of PHQ-9, Kessler-10, Kessler-6 and SRQ-20 in Butajira area health centers antenatal care clinics, Ethiopia [Certificate in Psychiatry]. [Addis Ababa]: Addis Ababa University; 2013.

57. Weathers FW, Blake, Schnurr PP, Kaloupek, Keane TM. The Life-Event Checklist for DSM-V (LEC-5). National Centre for PTSD, USA; 2013.

58. Weathers FW, Litz BT, Keane TM, Palmieri PA, Marx BP, Schnurr PP. Post-Traumatic Stress Disorder checklist for DSM-V. National Centre for PTSD, USA; 2013.

59. Spitzer RL, Kroenke K, Williams JBW, Löwe B. A brief measure for assessing generalized anxiety disorder: the GAD-7. Arch Intern Med. 2006 May 22;166(10):1092–7.

60. Zink T, Levin L, Putnam F, Beckstrom A. Accuracy of five domestic violence screening questions with nongraphic language. Clin Pediatr (Phila). 2007 Mar;46(2):127–34.

61. Mayston R, Habtamu K, Medhin G, Alem A, Habtamu A, Prince M, et al. Developing a measure of mental health service satisfaction for use in low income countries: a mixed methods study. BMC Health Serv Res. 2017;17:183; DOI 10.1186/s12913-017-2126-2.

62. Lauritsen JM, Bruus M. EpiData Entry (Version 3.1). A comprehensive tool for validated entry and documentation of data. Odense, Denmark: The EpiData Association; 2008.

63. WHO Verbal autopsy standards: ascertaining and attributing causes of death [Internet]. WHO. [cited 2018 Jan 12]. Available from: http://www.who.int/healthinfo/statistics/verbalautopsystandards/en/

64. Nelissen E, Mduma E, Broerse J, Ersdal H, Evjen-Olsen B, van Roosmalen J, et al. Applicability of the WHO maternal near miss criteria in a low-resource setting. PloS One. 2013;8(4):e61248.

65. O'Flynn E, Andrew J, Hutch A, Kelly C, Jani P, Kakande I, et al. The Specialist Surgeon Workforce in East, Central and Southern Africa: A Situation Analysis. World J Surg. 2016;40:2620–2627.

66. Derbew M, Laytin AD, Dicker R. The surgical workforce shortage and successes in retaining surgical trainees in Ethiopia: a professional survey. Hum Resour Health. 2016;14 (Suppl 1):29.

### SOUTH AFRICA: WORK PACKAGE 4

#### Promoting Person-Centered TB Care: A mixed method study

**Study Design:** Implementation research, health systems research, mixed qualitative and quantitative methods

**Research to be conducted by:**

Knowledge Translation Unit, UCT Lung Institute

**In Partnership with:**

Centre for Rural Health, University of KwaZulu Natal

NIHR Global Health Institute, Kings' College London

Norwich Medical School, University of East-Anglia

#### ACRONYMS and ABBREVIATIONS

| | |
|---|---|
| ART | Anti-Retroviral Treatment |
| CoBALT | Comorbid Affective Disorders, AIDS/HIV, and Long Term Health |
| DfID | Department for International Development |
| DHB | District Health Barometer |
| HCW | Health Care Worker |
| HIV | Human Immuno-Deficiency Virus |
| LMICs | Lower to Middle Income Countries |
| MDR TB | Multi-Drug Resistant Tuberculosis |
| MhINT | Mental Health Integration |
| MPAC | Mpumalanga Provincial Aids Council |
| NCD | Non-Communicable Disease |
| NSP | National Strategic Plan |
| OHU | Occupational Health Unit |
| PHC | Primary Health Care |
| PRIME | Programme for Improving Mental Health Care |
| SCRU | Soul City Research Unit |
| SSA | Statistics South Africa |
| TB | Tuberculosis |

| | |
|---|---|
| ToC | Theory of Change |
| WHO | World Health Organisation |
| XDR TB | Extensively Drug Resistant Tuberculosis |

### INTRODUCTION

South Africa's protracted struggle against a significant tuberculosis (TB) burden persists. The South African TB epidemic has been rendered all the more complex given cross-cutting epidemiological, socio-economic and political intersections. Along with a substantial human immunodeficiency virus (HIV) burden, TB often presents comorbid with a range of non-communicable diseases (NCDs) and mental illness. While comorbid mental illness has been frequently pointed out to be an important consideration in TB management, the bulk of research have focused on the linkages between TB and alcohol use disorders. Given the demands that TB treatment place on patients – including (but not limited to) the somatic dimensions of taking medications, the experience of disease symptoms and the side effects of medication regimens, and the manifestation of different kinds of stigma during the disease and treatment experience – depression is increasingly recognised as a prevalent comorbidity and associated with poor TB treatment outcomes. This being noted, the face-to-face dimensions of a depression counselling process raise potential challenges, the most obvious being the occupational risk of TB infection for counsellors.

South Africa has made impressive progress in scaling up the provision of ART services in the past decade, with around 3.5 million people now receiving treatment, leading to a rise in life expectancy from 54 to 62 years (1). However, TB is proving more challenging to address and has been embattled by the emergence of MDR and XDR strains which are diverting clinical, financial and operational resources. Globally, TB receives substantially less funding than HIV or malaria, and recent research has been dominated by the development and testing of new diagnostics, and shorter, alternative medication regimens. Comparatively little health systems research on TB care delivery has been done. This research leverages health systems advances from the area of mental health to address unmet needs in TB, and importantly the significant role that stigma plays in limiting effective implementation of new advances. The output of the research will be an adapted intervention specific to patients with TB that can then be tested in a definitive trial and scaled up for implementation across South Africa. Furthermore, current partnerships with other high burden countries such as Ethiopia and Nigeria offer opportunities to disseminate the findings and intervention from this research to those settings.

The proposed study builds on previous work that aimed to integrate mental health into primary healthcare (PHC) settings in South Africa, during which the verticalized nature of TB services, its distinct occupational risk, and overwhelming illness experience emerged as key challenges. An approach is required that tailors for a patient population on the peripheries of the socioeconomic landscape, who, along with their providers and households, experience a substantial degree of social stigma. This study builds on the scale-up of integrated mental health services as part of a CDC-funded initiative called MhINT (Mental Health Integration Project) in high TB burden areas. It draws from a mixed methods study design, including a scoping review and the development, implementation and assessment of stigma reduction, clinical communication skills and mental health interventions. The ultimate aim is to improve providers', patients' and households' experience of care, identify and treat co-morbid mental illness, improve TB treatment outcomes and strengthen uptake of infection control measures in clinics and homes.

In order to evaluate an adapted counselling intervention, the research team will map relevant contextual features that are considered likely to impact on the adaptation and delivery in each clinic. This will involve identifying:

- Macro: Policy of TB and mental health, funding arrangements, widely circulated cultural discourses of mental health, the importance and consequences of social stigma for people living with TB.

- Meso: local health area/facility policies or service protocols on management and treatment of patients with TB; the main social actors involved in determining intervention delivery (i.e. healthcare staff, patients, managers); and the different sites and scenes which impact on how the intervention is adapted and delivered (e.g. local area management meetings, space for delivering counselling intervention);
- Micro: the main (i.e. counselling sessions, consultations) activities and subsidiary activities (e.g. additional meetings, receptionist screening of patients) of intervention delivery; and interactional arrangements of each activity.

### BACKGROUND

#### TB burden and the need for a public health response

South Africa is among the top ten high burden tuberculosis (TB) countries globally with an estimated incidence rate of 781 cases per 100 000 (2). South Africa's TB epidemic has been driven by HIV, with an increase in TB cases of 400% over the past 15 years (3). An estimated 59% of people with active TB are also infected with HIV (2), and TB remains the leading cause of death in people living with HIV and the single leading cause of death overall contributing to 6.5% of deaths in 2016. MDR-TB has emerged as a growing global problem, and while the percentage of new cases with MDR-TB is estimated at 4.1% (2.8-5.3) and re-treatment cases at 19% (9.8-27), MDR TB now accounts for nearly 19% of the National TB Programme budget (2) and has contributed significantly to worsening stigma surrounding the disease. South Africa has consistently fallen short of the WHO's TB treatment targets to successfully treat 85% of cases. Eighty one percent of the 291 793 new and relapsed cases registered in 2015 were reported as having successfully completed TB treatment (2). However, this figure obscures large variations in outcomes across districts with some districts not even achieving 60% (4). The National Strategic Plan for HIV, STIs and TB (2017-2022) has four broad TB-related goals: (i) to reduce TB incidence by at least 30%, (ii) to implement the 90-90-90 strategy for TB, (iii) to address the social and structural drivers of HIV, TB and STIs and (iv) to reduce stigma and discrimination of people living with HIV and TB by 50% by 2022 (5).

A large challenge to achieving these targets is the role that stigma plays in pathways to and through care during the TB illness episode. TB infection prevention mechanisms, fear of infection and the serious socioeconomic challenges thereof leads to the manifestation of both internal and external stigma, in turn leading to TB diagnostic delays and treatment noncompliance (6-8). Significantly, the experience of social stigma is significantly related to the development of MDR-TB, due to poor treatment adherence (9).

Given the link between infection control knowledge and behaviour, there is a pronounced need to strengthen health communication channels outside the PHC facility sphere to households and communities (10, 11). Some HCWs are unwilling to work in TB programmes, are afraid of TB infection when interacting with patients (intensified after confirmed diagnosis), leading to TB patients being shunned, avoided and segregated (12-14). Stigma can also impede the implementation of measures to protect others in the home and other confined spaces such as public transport (15). TB patients are often discriminated against and stigmatised in communities, due to a fear of infection, links between TB stigma and other types of stigma (for instance HIV, poverty and mental illness), perceptions of links between TB and disreputable behaviour, and supernatural beliefs (16-18). In contexts of high HIV prevalence, TB stigma cannot be disentangled from HIV stigma, two diseases that play out in a "cursed duet" (19). Accordingly, a double stigma emerges, illustrated by studies in South Africa (20-22). Co-infected patients tend to foreground their TB status above their HIV status – HIV is avoided due to fears of acknowledging its burden. There are several inroads to mitigate TB stigma pathways. For example, clear communication about, along with the normalisation of, TB infection control measures; integrating TB literacy and counselling into household contact investigations; and engaging patients in decisions about auxiliary therapies, such as for comorbid depression (23).

#### TB burden in health care providers, stigma and the need for improved TB infection control and anti-stigma interventions for health care providers

Health care workers (HCWs) are at a three to four-fold increased risk for TB infection and disease, compared to the general surrounding population of health facilities (24-26). MDR TB also affects HCWs at a greater frequency than the communities they serve (27-29). In a cross-country analysis, both the prevalence and incidence of latent TB among HCWs have also been calculated to be highest in South Africa (30). Further, the rates of TB among HCWs in South Africa have been suggested to be extremely high irrespective of screening method used, explained to a considerable degree by occupational exposure (31). In a study among 28 public hospitals in South Africa, poor implementation of TB infection prevention measures, particularly the use of personal protective equipment, was significantly associated with TB disease among HCWs, (32). The increased risk of TB affects all healthcare personnel including community health workers, counsellors, administrative and support staff, laboratory workers and health science students (33, 34) Published data demonstrate that clinical staff (nurses and doctors) appear to be at highest risk (35, 36), however there is a dearth of data regarding ancillary support staff and community health workers. The non-profit sector have begun to respond to the growing recognition of the weight of the challenge of TB among HCWs, most notably the establishment of TB Proof, an organisation aiming to creating safer healthcare environments, destigmatising TB, and mobilising resources for TB prevention (37).

Although HCWs from lower socioeconomic backgrounds are also at high risk of TB exposure in their communities, studies that have controlled for living conditions confirm additional risk of TB disease attributable to workplace exposure (38). South Africa's high HIV prevalence rate could be another factor for HCWs' risk to acquire TB disease, though evidence is limited. A 2004 study suggested a 20.3% HIV prevalence rate among non-professional HCWs and 13.7% among healthcare professionals (39). In 2007, an overall prevalence rate of 11.5% among student nurses, nurses and physicians in a Gauteng study was suggested (40). In some countries, the close association of TB and HIV disease aggravates stigma surrounding a TB diagnosis, resulting in diagnostic and treatment delays among HCWs (41). Importantly the threat of occupationally-acquired TB and the adverse consequences for infected HCWs have contributed to attrition of these workers in the very region most challenged by severe shortages in human resources for health (42, 43). South Africa published TB infection control guidelines in 2007 (updated in 2012 and again in 2015) establishing a minimum standards for TB infection control practices for health facilities (44). Despite the availability of legislation, guidelines and policies, implementation of TB infection control is generally poor and adherence of HCWs is sub-optimal (45-47). Barriers to implementation include lack of resources, distrust of TB infection control efforts by HCWs, and disproportionate focus on individual level personal protections (48).

Stigma surrounding TB and/or HIV diagnosis among South African HCWs is also an important determinant of health-seeking behaviour and adherence to TB infection control recommendations (49). While in most high-income countries it is standard practice to isolate people who potentially are infected with TB, in many LMICs this rarely happens and is often perceived to be discriminatory (17, 50). TB diagnostic and treatment technologies (for instance, DOTS as a supervisory tool rather than a support tool) have been implicated to add to social constructions of stigma and to further entrench stigmatising attitudes (51). Nurses providing TB care are often stigmatised (52), while stigma and confidentiality concerns significantly influence HCWs' willingness to undergo TB screening and HIV testing (53). Further, witnessing TB stigma in health facilities render HCWs less likely to use the occupational health unit (OHU) in hospitals for screening and services (54). HCWs are in a unique position to be key change agents to increase awareness, lobby for resource allocation and to assist in implementing TB prevention interventions (55).

#### TB and common mental disorders and the need for TB-proof counselling interventions

Historically mental illness co-morbid with TB has focussed on alcohol use disorders (56) because of the shared hepatotoxicity of TB drugs and alcohol, and less on depression and psychological distress despite a growing number of studies suggesting very high rates of comorbidity (57-60). There is an established relationship between stigma and help-seeking behaviours, very often mediated by depressive symptoms. Perceptions of public stigma contribute to self-stigma experiences which in turn influence help-seeking attitudes and willingness (61). Stigma “gets under the skin” of people experiencing distressing symptoms by playing a causal role in rumination in stigma-distress relationships (62). By becoming internalised, stigma directly affects attitudes towards treatment, and poor treatment adherence is associated with higher levels of stigma and depressive symptoms (63-66). Stigma enhances vulnerability to mental health challenges, reduces treatment self-efficacy and causes concern regarding disclosure (67, 68). Although there is a well-established link between depression and non-adherence (61, 63-71) efforts to improve TB treatment outcomes have not prioritised identification and management of depression, and research in this area remains scarce and mainly among MDR TB patients or those hospitalised for treatment (72). Provision of depression interventions during TB treatment is not without its challenges. Isoniazid, which forms part of the backbone of TB treatment regimens through the intensive and continuation phase of treatment, interacts with commonly used antidepressants such as fluoxetine as well as being linked as causing depression(69); and counselling interventions require significant face-to-face time, imposing significant occupational risk to the counsellor. The need for innovations to existing mental health care programmes that reduce occupational risk to the counsellors is urgently needed.

#### Leveraging existing primary care interventions for health care providers and patients

A large treatment gap of 75% exists for depression, and case detection is highly unlikely to improve without provision of additional mental health resources (73). Since 2011 we have been working to develop, evaluate and scale-up interventions to address common mental disorders, mainly depression, in primary care in South Africa as part of the DfID-funded PRIME (Programme for Improving Mental Health Care) programme in the North West Province of South Africa (74, 75). The focus on depression was guided by burden of disease data which shows that depression accounts for the major proportion of mental illness and is among the top three causes of Years of Life Lived with Disability in 146 countries [1]. We have used a collaborative stepped care model led by nurses (detection and case management of depression), complemented by referral services to primary health care doctors (initiation of antidepressants) and clinic-based counsellors (manualised depression counselling). The clinical component is based on the combination of simplified, integrated care guidelines, educational outreach training to nurses, and enhanced prescribing provisions for nurses. Its origins were in the South African adaptation of the WHO’s Practical Approach to Lung Health which aimed to strengthen passive case detection of TB through integrated case management of people presenting to primary care with cough and difficulty breathing. Three randomised trials in South Africa have shown substantial improvements in TB case detection without compromising TB treatment outcomes (76-78). The programme has subsequently been expanded to include all common conditions in adults presenting to primary care and integrates content on communicable diseases, NCDs, mental illness and women’s health (79). In the Western Cape Province, it has also been expanded to include management of MDR TB as these services have been decentralised to primary care. A global version of this programme now known as PACK (Practical Approach to Care Kit) is available through a partnership with the British Medical Journal Publishing Group (pack.bmj.com) and adaptations are being implemented in Botswana, Malawi, Brazil, Nigeria and Ethiopia.

The PRIME work in the North West province has focussed on strengthening the mental health components of the programme, and the addition of clinic-based counsellors to provide manualised depression counselling originally developed and pilot tested in the KwaZulu-Natal Province (80) and addressing the key triggers for depression in this population including poverty, interpersonal conflict, grief and loss, and stigma. A pair of trials is underway to evaluate the effects of this intervention on mental and physical outcomes depression co-morbid with HIV and hypertension, the two physical conditions which account for the lion’s share of chronic conditions seen in primary care. The HIV trial (CobALT - Comorbid Affective Disorders, AIDS/HIV, and Long Term Health) is funded by the National Institutes for Mental Health (81) and the hypertension trial (PRIME) by DfID (82). Data collection was completed in January 2018 and data cleaning and analysis are now underway. Already we are working in collaboration with the respective provincial Departments of Health in one of the trial districts (Dr Kenneth Kaunda district) in the North West province and a further two rural districts (Amajuba in Northern KwaZulu-Natal, and Ehlanzeni in Mpumalanga) to scale up components of the intervention and embed them into routine services as part of MhINT (Mental health Integration Project). This involves expanding facility-based counselling services at clinics where these previously had been limited to the provision of HIV counselling and testing, as well as strengthening the nurses and doctors' clinical training in mental health as well as their clinical communication and emotional coping skills. The latter were identified as important to better equip nurses to identify and address signs of common mental disorder in the consultation sessions in the formative phase (75); as well as being important non-technical skills for promoting patient centred care. Facility and district-led Continuous Quality Improvement (CQI) cycles are also part of the scaling up programme to optimize embedding of the programme.

Exposure to the counselling intervention among TB patients in the trials and the scale up sites is low. Nurses were asked not to refer TB patients to the counsellors given that we had not put in place sufficient measures to protect counsellors from contracting TB and that in practice counselling required substantial face-to-face time often in confined settings to ensure privacy. This new research project aims to build on this existing programme, through expanding the intervention package to promote person centred TB care through i) engendering greater TB infection control at the facility level, as well as provider self-care that should assist to reduce stigmatizing attitudes through reducing the fear of contagion; ii) Reduce stigma in persons struggling with TB through improving coping strategies, as well as empowering persons struggling with TB to minimize infecting loved ones during the infectious stage; and iii) adapt the existing counselling programme to meet the needs of depressed persons struggling with TB at the appropriate stage during the TB illness episode while safeguarding clinical and counselling staff and managing their occupational risk of TB.

### RESEARCH OBJECTIVES

The Specific Research Objectives of this study are to:

1. To complete a scoping review to describe the prevalence of depression, psychological distress, stigma and alcohol use disorders among patients attending TB services, their relationship to each other and TB treatment outcomes, and linkages to antiretroviral treatment (ART) programmes.
2. Assess the current status of TB control programmes, organisation of TB care and implementation of infection control measures recommended as part of the National Health Department's TB Infection Control Policy in two potential pilot districts.
3. Develop an understanding of TB stigma and understand their concerns and needs that can inform the expansion of the existing provider and patient programme to better meet the needs of TB management in primary care facilities.
4. Elucidate the relative contributions, relationships and sequencing of stigma, infection control issues, symptoms, psychological distress and comorbidities during different stages of TB care with a view to identifying entry points that might be addressed through scalable interventions.
5. Adapt our existing depression and alcohol use counselling intervention for use with TB patients, strengthening the focus on stigma reduction.
6. Adapt our existing clinical and communication skills and self-care intervention for health workers (nurses, doctors, counsellors, community health workers) to reduce stigmatizing attitudes, strengthen individuals' agency to protect themselves appropriately from occupational TB and promote more person-centred TB care.
7. To complete preparatory work needed to finalise a follow-on evaluation of the impact of the modified intervention on stigma among TB patients including: (i) collation of facility-level TB data from the Ehlanzeni district, (ii) review of stigma measures in TB patients and (iii) power calculations to inform a robust yet feasible evaluation design.

### RESEARCH METHODS

#### Phases

The research will be divided into two work phases:

1. *Phase 1 (*Diagnostic Phase; specific objectives 1 -3; months 1-12):
   i. A scoping review will be conducted to establish the epidemiology of depression, alcohol use disorder, psychological distress and stigma among patients attending TB services, their relationship to each other and timing during the TB treatment episode, association with outcomes including successful linkages to antiretroviral treatment (ART) programmes. The enhanced scoping reviewing framework developed by Levac et al. from the original work of Arksey, will be utilised as it increases the clarity and rigor of the review process (83, 84).
   ii. A situational analysis will be done at a sample of PHC facilities in participating in the MhINT programme in the Ehlanzeni and Amajuba districts. This will include a facility profile to assess the current status of TB control programmes, the organisation of TB care and implementation of infection control measures recommended as part of the National Health Department's TB Infection Control Policy,
   iii. Formative individual interviews and focus group qualitative interviews will be conducted as part of the situational analysis with 1) TB patients and service providers in the Ehlanzeni and Amajuba districts; 2) TB Proof members; and 3) service providers who successfully manage to implement infection control measures to better understand the facilitators to doing so and how they communicate these to patients. Preparatory work for the follow-on evaluation of the impact of the modified intervention on stigma and mental health of TB patients in the Ehlanzeni and Amajuba districts. This will include: (a) Collation of facility-level TB data from MhINt clinics; (b) Review of stigma measures in TB patients and providers; and (c) Selection of an appropriate evaluation design adequately powered to detect a policy-important change in stigma among TB patients in line with the National Strategic Plan Objective 4 of reducing stigma among TB patients by 50%.
2. *Phase 2* (Adaptation and Piloting of the Intervention; specific objectives 5-7; months 13-20):

At the end of Phase 1, we will make an informed decision about whether or not to focus subsequent efforts in the Ehlanzeni or Amajuba sub-districts.

   i. A Theory of change workshop will be arranged with relevant district stakeholders to better elucidate the relative contributions, relationships and sequencing of stigma, infection control issues, symptoms, psychological distress and comorbidities during different stages of the TB illness episode with a view to identifying entry points that might be addressed through scalable interventions. The intervention will be co-designed with the district stakeholders underpinned by a Theory of Change Map of the intervention, linking content, mechanisms of action and intervention outcomes.
   ii. The Theory of Change Map will be used to adapt our existing intervention, comprising manualised counselling delivered by mid-level counsellors, and clinical and communication skills training for nurses and doctors of TB. We will expand the stigma reduction elements, and explore whether the introduction of visible infection control measures (e.g. N95 masks) can be used to initiate supportive conversations with patients, to reduce fear, provide containment regarding management of their infection control risk and contact tracing in their households, strengthen identification of co-morbid mental illness, and promote a person-centred approach to encourage retention in care and adherence to treatment. We will pilot the TB-customised intervention in two successive phases of implementation, first in one clinic, and second in three clinics. A case study approach will be used to understand feasibility and acceptability of the intervention and guide refinement of the intervention. A

range of mixed methods will be adopted (semi-structured interviews, focus group discussions, direct observation) to evaluate intervention delivery. Findings will be used to refine the adapted intervention components as well as underpinning Theory of Change map.

#### Scoping Review

Published and unpublished primary studies and reviews will be identified by two reviewers through a three-step search strategy. Scoping questions to be used include:

i. What is the epidemiology of mental conditions (including substance use) among patients attending PHC-level TB services.
ii. How can stigma be measured?
iii. What are the attitudes towards TB?
iv. What are the perceptions, experiences and attitudes regarding TB infection control?

In the initial step, a search of two databases MEDLINE and CINAHL will be completed. This will be followed by an analysis of the text words contained in the title and abstract of retrieved papers, and of the index terms used to describe the articles. A second search using all identified keywords and index terms will then be undertaken across all included databases. Thirdly, the reference list of all identified reports and articles will be searched for additional studies. A logical and descriptive summary of the results that aligns with the objective and question/s of the scoping review will be completed.

#### Situational Analysis

The situational analysis will be conducted in four to six purposively selected primary care facilities in the Ehlanzeni and Amajuba districts. A case study approach will be used with mixed methods to develop a general evaluation framework with three components: the 'contextual' component will investigate the social, demographic and organizational context (85); the 'intervention' component will analyse the counselling intervention's underlying conceptual orientation, organisational processes guiding delivery; and the 'outcomes' component will investigate changes affected and participant's experiences of this complex intervention. Data collection methods will include a facility profile, individual interviews and focus groups, observations and a Theory of Change workshop.

#### Research Setting

The research sites for the first phase are Ehlanzeni (Mpumalanga) and Amajuba districts (KwaZulu Natal) in South Africa. According to the 2013 District Health Barometer (DHB) the incidence of TB smear positive cases was 191.5 per 100 000(86) in Ehlanzeni and 225 per 100 000 in Amajuba district (86). As work is currently being done both of these districts as part of the MhINT project, they were selected as most appropriate for the diagnostic phase. At the end of that phase a decision will be taken on which district to focus subsequent phases including adaptation and pilot implementation of the intervention. Factors that will be taken into account include the scale of the TB burden in the district, progress with embedding the MhINT intervention and capacity to absorb an add-on module for TB clients.

#### Study Population and Data Collection Methods

##### *Study Population*

*Staff*: Purposive sampling will be used in recruiting managers to provide the necessary information for the facility profile, and in the recruitment of nurses, doctors, counsellors and health care workers who are treating patients with TB at each of the primary care facilities in the selected districts. For the piloting of the intervention, staff who are trained in the adapted intervention will be asked for consent to observe and/or video/audio record their consultation and to participate in a focus group about their experience of the intervention. This will only be applicable where the relevant patient has also provided the required consent.

*Patients*: The recruitment of patients will be determined through theoretical sampling and assisted by the nurse in the clinic who will identify who the research team can approach to participate in individual interviews. Recruitment of patients will stop once data saturation is achieved, and no new themes are emerging in interviews. For the piloting of the intervention, patients who receive the adapted intervention will be asked for consent to observe their consultation and video or audio-recording will be used according to the participant's preference. Patient's will also be asked to be interviewed about their experience of the adapted intervention in Phase Two.

*Stakeholders*: The recruitment of key stakeholders for the formative qualitative interviews and Theory of Change workshop will be through purposive sampling.

#### *Data Collection Methods*

##### *Phase 1: Data Collection Instruments*

###### Facility Profile

A facility profile will be conducted with the primary care facility managers using the Facility profile adapted from MhINT situational analysis tool.

###### Individual Interviews and focus groups

The participants who will be individually interviewed will be patients with TB, and focus groups will be conducted with nurses, doctors, managers, health care workers and counsellors working at the selected primary care facilities.

The focus groups and interviews will be semi-structured and carried out in the language most appropriate to each setting, audio recorded, and transcribed verbatim. Once informed consent has been obtained, the researcher will check the participant is willing for the interview to be audio-recorded, explaining the reasons for doing so. If a participant does not wish the discussion to be recorded, the researcher will make written notes. Participants will be reassured that neither the transcript of the interview/focus group nor the handwritten notes will contain any personal identifying information and that nobody will listen to the audio-recording or read the notes, except for members of the research team involved in transcribing and/or analysing the data.

Individual in depth interviews with patients willing to participate will explore perspectives of TB stigma, triggers of psychological distress, experiences and perceptions of infection control measures (see Appendix 5).

Focus groups with nurses, counsellors, community health workers and doctors will be conducted with staff willing to participate to explore: infection control measures currently in use, perspectives of infection control requirements, reasons for level of infection control measures taken (not taken), barriers to identifying patient's mental health needs, perspectives of the existing MhINT intervention, barriers and facilitators of using the counselling intervention within the TB programme, the consequences of the intervention for how staff protect themselves from TB infection, and for how the intervention triggers discussion of stigma, seriousness of condition and infectivity (see Appendix 6).

Interviews with provincial managers and policymakers will explore current services and interventions to support people with TB, interventions to reduce TB infection and address mental health needs of patients with TB with and without and co-morbid mental health disorders, and perspectives on required interventions to improve service delivery at a system wide, organisational and individual clinical level (see Appendix 7).

#### Non-Clinical Observations

There will be two aspects to the observational component of the case study approach, the first will be in Phase One of the study. The research team will carry out direct observation of the clinic flow for TB care and infection control measures implemented within non-clinical areas, such as waiting rooms and TB room' areas (Appendix 8). The researcher will record contemporaneous written or audio-taped field notes of any activities which relate to standard TB care and infection control measures implemented within each of the selected clinics. The researcher will take field notes of their observations and ask staff questions to clarify the researcher's observations. The researcher may document quotes in their field notes, but no staff member or patient will be named in the field notes.

#### *Phase 2: Data Collection Instruments*

##### Theory of Change Workshop

The Theory of Change workshop will include key stakeholders identified during the situational analysis at each primary care facility to elicit the relative contributions, relationships and sequencing of stigma, infection control issues, symptoms, psychological distress and comorbidities during different stages of the TB illness episode with a view to identifying entry points that might be addressed through scalable interventions.

Participants will be asked to determine TB pathways; how each point in the pathway interacts with stigma, psychological distress and symptoms; the determinants of stigma and psychological distress across macro, meso and micro contextual levels; the mechanisms by which different interventions may affect which patient outcomes; and the feasibility of implementing different intervention components.

##### Consultation Observations

The second aspect of the observational component of the case study approach involves consultation observation of the adapted intervention.  This will involve a researcher video-recording or audio-recording (dependent on the participants' preference), observing and taking field notes of clinical consultations and counselling sessions. In practice, this is likely to involve a researcher being present whilst the relevant member of staff consults with a patient (Appendix 13). Video-recordings or audio recordings will be transcribed and analysed to understand how consultations and counselling are implemented and the consequences of the intervention for how patients respond to the use of masks and other infection control measures, and for what types of discussion are facilitated or constrained within the counselling sessions and consultations. These insights will enable the intervention to be refined to better facilitate integration into routine care including the identification of training for staff in optimising intervention delivery.

In order to systematically investigate pilot implementation of the adapted clinical, counselling and systems intervention, and how this is maintained throughout the process of adaptation and implementation, the research team will use the map of contextual features to identify likely tensions, 'moments of vulnerability', to processes of implementation. Such moments are likely to be located within the patient consultations and counselling sessions, in particular, tensions between the protocol of the intervention and how it is integrated into routine practice by staff and then received by patients. Identifying moments of vulnerability will enable us to track the implementation of the intervention across different contextual levels, as observed within qualitative ethnographic, interview and interactional data and provide a framework for identifying intervention mechanisms and components within the Theory of Change workshop.

##### Individual Interviews and focus groups

The participants who will be individually interviewed will be patients with TB who have received the adapted intervention, and focus groups will be conducted with nurses, doctors, managers, health care workers and counsellors who implement the adapted intervention at the selected primary care facilities.

The focus groups and interviews will be semi-structured and carried out in the language most appropriate to each setting, audio recorded, and transcribed verbatim. Once informed consent has been obtained, the researcher will check the participant is willing for the interview or focus group to be audio-recorded, explaining the reasons for doing so. If a participant does not wish the discussion to be recorded, the researcher will make written notes. Participants will be reassured that neither the transcript of the interview/focus group nor the handwritten notes will contain any personal identifying information and that nobody will listen to the audio-recording or read the notes, except for members of the research team involved in transcribing and/or analysing the data.

#### **<u>Data Analysis</u>**

##### *Interviews/focus groups*

All interviews and focus groups will be transcribed verbatim and in the first instance thematically analysed using NVivo software. This will be to provide detailed staff, managers, policymakers and patient perspectives of the process and content of infection control measures within clinics. We will then develop a coding scheme for the identified themes and structure according to different contextual levels and mechanisms of impact. A constant comparison approach will be adopted, working iteratively between data obtained from different interviewees within and between clinics.

##### *Ethnographic Observations*

Video recordings and field notes will be analysed thematically in the first instance to provide a detailed description of process and content involved in infection control implementation within clinics. We will then analyse the ethnographic data to empirically observe how infection control measures are delivered and received by patients, how they trigger discussions of stigma with patients and tensions between the delivery and receipt of measures to protect staff from infection.

##### *Theory of Change workshop*

The workshop will be audio-recorded and transcribed verbatim, then analysed to describe the TB care pathway. Using open and axial coding the transcript will then be analysed for how the pathway interacts with stigma, psychological distress, and the mechanisms by which different intervention components function to achieve which outcomes.

##### *Qualitative data synthesis*

The collection and analysis of qualitative data will be iterative, moving between data collection and data analysis to test emerging theories. It may for example emerge that some professionals have particular perceptions about infection control measures, which shape their experience and use of the intervention, and this may require deeper exploration. The analysis of the observational ethnographic data will therefore require knowledge from professional interviews to compare how reported experience relates to actual implementation of infection control measures. Care will be taken to identify and follow up deviant cases which do not fit into emerging theories.

In drawing case comparisons across clinics, we will develop hypotheses about why the intervention is linked to outcomes. This may lead us to identify factors which are plausibly and/or consistently related to successful or unsuccessful delivery of the components of the intervention. Emerging theories, and the relationship of the data to the conceptual literature underpinning the intervention will be discussed and refined at team meetings throughout the research.

#### Data Management

All personal information obtained about patients or staff for the purposes of recruitment or data collection (e.g. names, addresses, contact details, personal information) will remain confidential and held in accordance with the South African Protection of Personal Information Act No 4 of 2013 (88) and with the United Kingdom (UK) Data Protection Act 1998 (87).

> Each participant included in the study will be assigned a research number and all data will be encrypted and stored without the participant's name or address. Electronic data will be held on a secure database on a password-protected computer at the Knowledge Translation Unit in UCT Lung Institute, and paper-based information held in a locked filing cabinet in the research team office. Access to data will be restricted to the research team. Transcribers will have signed confidentiality agreements.

Names and participant details will not be passed onto any third parties and no named individuals will be included in the write up of the results. The only time personal information would be passed on to a third party would be if we considered there was risk of serious harm to a research participant, and this would only occur after discussion with the person concerned.

#### Data Dissemination

The engagement and dissemination strategy and activities will follow an integrated (as opposed to end-of-grant) knowledge translation approach, with *continuous* stakeholder engagement guided by an explicit dissemination framework with dissemination tools and channels tailored to a range of stakeholders. This approach is informed by the WHO TDR's 2014 Implementation Research Toolkit.

The intervention developed during PRIME is already been scaled up to four other districts and the South African researchers (Fairall, Petersen, Lund) are actively engaged with the National, Provincial and District Health Departments to explore how best to expand and strengthen provision of counselling services at clinic-level, whether through the appointment of additional designated counsellors or extending the scope of existing HIV counsellors. This project would feed directly into that work, and if successful, provide greater impetus to the argument for long-term resourcing for this cadre of staff. It would also inform the current debate as to whether to persist with an integrated training curriculum for Adult Primary Care, or whether to suggest the adoption of the modular approach used in the Western Cape, which would allow greater exposure to the mental health content, and alignment with other health systems strengthening measures.

### ETHICAL CONSIDERATIONS

Research inquiry is dependent not only on the expertise of the researcher, but also on honesty, integrity and good clinical practice (89). Use of ethical principles should pervade the research from the conceptualisation through to the dissemination of findings (90). The protocol complies with the Declaration of Helsinki (91).

#### Institutional and Gatekeeper Permission

Ethical approval from the Ehlanzeni and Amajuba Departments of Health is being sought as part of the MhINT project and ongoing work in these districts.

#### Ethical Principles

The core ethical principles of research for vulnerable populations will be employed in the study and include the following:

##### *To Protect Research Participants from Harm*

This includes both emotional and physical harm that might result from the research. It also means to protect their rights and interests. As this study is a usual clinical care with supplementary counselling and case study approach, no invasive procedures are being implemented and the risk of physical harm is non-existent. The research team will ensure that no interview or observation occurs in cases where urgent care is required or where the research team could interfere with routine care. Furthermore, the research team has experience and the knowledge to discern a suitable time for conducting interviews which will be so as not to place participants at risk. For staff focus groups, the research team will accommodate the schedule of the staff member, so as not to cause disruption to clinical care on undue stress.

##### *To Ensure the Safety of Research Participants*

Research participation in the study is voluntary. "Voluntary participation" denotes that informed consent to take part in the study must be obtained before any research tools are used. "Informed consent" means that a participant has agreed to take part in the study after being informed of and understanding the following: research topic, the purpose, the methods and processes; what the data will be used for; the participant may withdraw from the study at any time and that it is possible for participants who have withdrawn from the study to return to the research at any time. Patients will provide written informed consent to be interviewed (Appendix 1) and provide written consent to have their consultation be observed (Appendix 9). Staff will provide written consent to participate in focus groups (Appendix 2) and provide written informed consent to have consultations video-recorded and observed (Appendix 10). All participants will be provided with a research information sheet (Appendix 1-3; 9-10). A researcher will briefly introduce the study and will allow participants the opportunity to ask questions. A consent form will be provided for participants to sign. The researcher will also sign the consent form. The research participant will be given a copy of their signed consent form to keep, and a further copy will be retained by the research team.

The process of obtaining informed consent will not be hurried and the individuals will be given time to decide on participation. The technique of asking individuals to repeat what the researcher has told them about the research will be used to ensure understanding prior to obtaining informed consent. Written record of informed consent will be kept confidential and stored in a file that will be kept in a locked cupboard at the KTU. A copy of the consent form will be issued to each participant if they want one.

##### *To Respect Cultural Traditions, Norms and Customs*

The researchers are of South African origin and are aware of the diversity of cultures and backgrounds. This will not impede the research as sensitivity to diverse cultures, customs and norms will be respected. The interviews will be conducted in English, SiSwati or isiZulu depending on what each participant is comfortable with.

##### *To Establish as Much Equality as Possible*

Every attempt to minimise the power inequalities between researchers and participants will be made. The research team will use research methods conducive to working with vulnerable groups. This means that the research team will use words that participants can understand. The research team will allow sufficient time to build trust with participants and explain the research study.

##### *To Avoid Raising Unrealistic Expectations*

The research team will be clear and honest about the research and what will be done with the information gathered. The research team will inform the participants what they should expect and that their participation in the study will not be reimbursed. The research will primarily give voice to the participants, which in effect will hopefully benefit future generations.

##### *To be Fair*

The interviews will be scheduled at the convenience of the participants, so as not to conflict with personal time and commitments. Venues in which the interviews will be conducted, are convenient to the participant as they will be at the clinic.

##### *To Respect Privacy*

If the participant did not wish to take part in any part of the research, the participant's request will be respected and upheld. The participants will be clearly informed that their participation or decision not to participate will not affect them in any way.

##### *To Ensure Confidentiality*

All personal information obtained about patients or staff for the purposes of recruitment or data collection (e.g. names, addresses, contact details, personal information) will remain confidential and held in accordance with the South African Protection of Personal Information Act No 4 of 2013 (88) and with the UK Data Protection Act 1998 (87).

> Each participant included in the study will be assigned a research number and all data will be encrypted and stored without subject's name or address. Electronic data will be held on a secure database on a password-protected computer at the Knowledge Translation Unit in UCT Lung Institute, and paper-based information held in a locked filing cabinet in the research team office. Access to data will be restricted to the research team. Transcribers will have signed confidentiality agreements.

Names and participant details will not be passed onto any third parties and no named individuals will be included in the write up of the results. The only time personal information would be passed on to a third party would be if we considered there was risk of serious harm to a research participant, and normally this would only occur after discussion with the person concerned.

4. SCRU. Literature review of TB in South Africa. Soul City Research Unit; 2015.

15. Organization WH. WHO policy on TB infection control in health-care facilities, congregate settings and households. 2009.

44. NDOH. The draft national infection prevention and control policy for TB, MDRTB and XDRTB. Pretoria2012.

84. Arksey H OML. Scoping studies: towards a methodological framework. Int J Soc Res

Methodol. 2005;8(1):19-32.

85. Murdoch J. Process evaluation for complex interventions in health services research: analysing context, text trajectories and disruptions. BMC Health Serv Res. 2016;16(1):407.

86. MPAC. Annual Progress Report 2014/2015. 2016.

87. Data Protection Act 1998, (1998).

88. Protection of Personal Information Act No 4 of 2013, (2013).

89. Burns NG, S.K. The Practice of Nursing Research: Appraisal, Synthesis and Generation of Evidence. Sixth ed: Saunders; 2009. 750 p.

90. Botma YW, S.C.D.; Mulaudzi, F.M.; Greeff, M. Research in Health Sciences. South Africa: Pearsons; 2010.

91. Association WM. Declaration of Helsinki. 2013.

### SOUTH AFRICA: WORK PACKAGE 5

#### Integrated palliative care and primary health care

Professor Richard Harding, Cicely Saunders Institute, King's College London

Dr Liz Gwyther, Dept Palliative Medicine, Faculty of Public Health & Family Medicine, University of Cape Town

Professor Irene Higginson, Cicely Saunders Institute, King's College London

Dr Wei Gao, Cicely Saunders Institute, King's College London

Dr Matthew Maddocks, Cicely Saunders Institute, King's College London

7th March, 2018

#### Summary

The majority of deaths globally are attributable to non-communicable disease (NCD), and four-fifths of these deaths are in low and middle-income countries. COPD is currently the fourth leading cause of death globally and is predicted to be the third leading cause of death by 2030. A review of the evidence of patient and caregiver views in advanced COPD found symptoms and concerns that span the dimensions of palliative care. The majority of these patients will suffer from pain, dyspnoea and other physical symptoms, and may require support with psychosocial or spiritual problems as their diseases progress. After the first hospitalisation for an exacerbation, 50% of patients die within 3.6 years. Research has shown palliative care alongside usual care for people living with COPD can reduce breathlessness and hospitalisations, improve quality of life and distress, satisfaction, dignity and survival. The standard of care of COPD patients in South Africa is described in the Guideline for the management of chronic obstructive pulmonary disease (2011). There is no guidance on palliative care interventions that improve the quality of life of patients with COPD.

**Aim:** To develop a feasible and acceptable primary care-led integrated palliative care intervention for patients and families living with COPD. Objectives: 1) To appraise existing interventions for COPD palliative care; 2) To appraise routine and acute primary care health service usage by COPD patients in the Western Cape; 3) To typologise primary care settings in the W Cape; 4) To measure the prevalence of symptoms and concerns among COPD patients attending primary care; 5) To describe stakeholder views (patients, staff, primary care health care professionals) on symptoms and concerns, management, and staff training needs; and 6) To integrate findings from objectives 1-5 to generate an intervention underpinned by the theory of person-centredness and a model of intended action.

**Target population:** People living with COPD in the City of Cape Town.

**Study setting:** The study will take place in 8 sites: Two primary care district hospitals, three 24hr Community health centres with Emergency units, and three 8hr Community Health Centres with emergency rooms. Eligible patients and families will be identified by their treating clinical teams and the researcher will introduce the study to them. If they wish to participate they will be given 24 hours to consider their decision and then arrange to meet the researcher at the clinic or at their home. Staff will be directly approached by the study researcher and follow the same procedure.

The facility manager/senior doctor will be interviewed at each site will be interviewed to understand the current care for COPD patients. Socio-demographic data and clinical data will be collected from COPD patients (sample size 385) who will also complete a symptom assessment questionnaire (MSAS), the African Palliative

Outcome Scale (POS) and the Center for Epidemiologic Studies Depression Scale (CES-D). Severity of breathlessness on exertion over the previous 24 hours will be measured using the London Chest Activity of Daily Living Questionnaire.

Cross-sectional in-depth qualitative interviews with a purposive sample of stakeholders, adult patients, family members and facility staff. Patients and family members will be asked about primary needs and concerns, what would make care more patient centred and how should that be delivered, current self-management, preferences for advance planning and communication practices, views on potential intervention components, decision making and drivers of emergency unit attendance. Staff will be asked about value of patient-centredness, barriers and training/mentorship needs, leadership requirements, implementation, communication skills needs, anticipated challenges and benefits.

Data transfer between UCT and KCL will be of Epi Data files of anonymised quantitative data, and Word files of anonymised qualitative transcripts. Analysis: For all measures we will conduct descriptive analysis to report prevalent concerns then conduct regression analyses and identify predictors of scores for each measure integrate the prevalence data from this objective with the qualitative data from the subsequent objective to determine the primary and secondary outcomes of the intervention.

### Justification

The majority of deaths globally are attributable to non-communicable disease (NCD), and four-fifths of these deaths are in low and middle-income countries [1]. By 2030 it is projected that deaths due to NCDs will be the most common causes of mortality in developing countries [2], attributable to a combination of increasing and aging populations, the adoption of risk-factor lifestyles such as increasing tobacco consumption, and poor access to diagnostic, preventative and curative treatment services. It is anticipated that by 2020 the largest increases in NCD deaths will occur in Africa (www.who.int/mediacentre/factsheets/fs355/en/). In South Africa, WHO estimates place the burden from NCD at two to three times higher than developed countries [2]. National and Western Cape Province data reveal a high proportion of deaths from diseases of the circulatory and respiratory systems [3] [4].

COPD is currently the fourth leading cause of death globally (http://www.who.int/gard/publications/GARD_Manual/en/), and is predicted to be the third leading cause of death by 2030 [3] A review of the evidence of patient and caregiver views in advanced COPD found symptoms and concerns that span the dimensions of palliative care, i.e. physical, psychological, social and spiritual [4]. The vast majority of these patients will suffer from pain, dyspnoea and other physical symptoms, and may require support with psychosocial or spiritual problems as their diseases progress [5].

After the first hospitalisation for an exacerbation, 50% of patients die within 3.6 years [6]. COPD patients are more likely than lung cancer patients to die in hospital [7], The disease course is often unpredictable, with sudden and life-threatening exacerbations, often leading to ad hoc decisions about how to proceed. However, COPD patients rarely discuss advance care plans or access any palliative care, with doctor attitudes, difficulties in prognostication and poor communication the most commonly reported reasons [8]. A systematic review of resource use and health care costs of COPD patients at the end of life found increased hospitalisations, intensive care unit stays, primary care consultations and medication prescription, as well as a lack of palliative care services [9]. Evidence also suggests that COPD can lead to greater financial hardship [10], which carries great implications for patients and families in LMIC.

#### Policy

Despite the relative historical neglect of NCDs within palliative care [11], there have been a number of important related recent initiatives: in September 2011 the United Nations held the first High-Level Meeting on NCDs, at which Resolution 66/2 on the Prevention and Control of Non-Communicable Diseases was adopted by the General Assembly. The resolution included the recognition of palliative care as an integral part of the response to such illnesses – using access to palliative care assessed by morphine- equivalent consumption of strong opioid analgesics (excluding methadone) per death from cancer – and supplemented the WHO's 2008-2013 Action Plan for the Global Strategy for the Prevention and Control of Non-Communicable Diseases, which include a commitment to increase the availability of palliative care. In May 2014, the World Health Assembly Resolution 67.19 urging member states to implement and monitor palliative care actions that were included in the agreed World Health Organization's (WHO) global action plan for the prevention and control of NCDs.

#### The effectiveness of palliative care

Palliative care is a global human right [12], enabling patients and families to live well with progressive illness, improving their outcomes, and to save costs by reducing unplanned admissions and futile treatments [13; 14; 15; 16; 17]. The World Health Assembly (WHA) resolution on palliative care (Resolution 67.19, 2014) calls for palliative care "integrated throughout the life course" and for novel data on effective interventions [18]. RCTs from high income countries have found that early palliative care improves quality of life and care, symptom control, patient and caregiver satisfaction, survival, family depression, and costs [19; 20; 21; 22; 23].

#### Current evidence in COPD palliative care

Palliative care for hospitalised organ failure patients in South Africa increases rates of home death and reduces length of stay [17]. However, despite evidence of high palliative care needs among those with non-malignant disease, their access to palliative care is generally poorer than for those with a malignancy [5; 24].

An integrated service for patients with progressive illness experiencing breathlessness [25] (54% sample had COPD diagnosis) provided the following service components: joint 1st appointment between respiratory and palliative care, information, self-management guidance including a "pacing" guide, a hand-held fan or water spray, a poem, and a crisis plan. It found an intervention advantage for both the primary endpoint (mastery of breathlessness) and survival. A further trial of a breathlessness intervention found cost savings, and qualitative benefit particularly in relation to anxiety and professional guidance on symptom control and supportive information [26].

Systematic reviews have found self-management can reduce breathlessness and hospitalisation, improve quality of life and distress, satisfaction, dignity and survival [27; 28; 29]. Reported strategies to avoid emergency department admission include goal setting, monitoring and risk management, and building partnerships between primary care and respiratory medicine [30]. A systematic review of advance care planning preferences for COPD patients found that while the majority of patients would wish to engage in ACP, it was rarely offered, the main reason being complex and unpredictable disease trajectories, professionals' desire for hope among patients, and poor communication skills regarding sensitive topics [31]. When discussions regarding palliative and end-of-life care are conducted with COPD patients, they are usually in far advanced disease stages and in acute settings [32]. This is contrary to evidence from an RCT of ACP among patients with severe lung disease, which found a higher than anticipated preference to receive the intervention [33].

#### Methodological issues in evaluating palliative care in COPD

A clinical COPD palliative care trial found no quantitative benefit for the intervention despite qualitative description of intervention advantage [26]. The authors suspected a therapeutic effect from the interviews, which may be greater given the relative isolation of COPD patients and the lack of attention to their palliative care symptoms and concerns. We will similarly be informed by our recommendations from an RCT conducted among a marginalised population in sub-Saharan Africa, which found an effect from researcher contact and the participation expenses paid [34]. Given the potential challenges to recruiting and retaining patients in palliative care, the specific nature of relevant outcomes in progressive illness, and the ethical issues of managing and measuring symptoms and concerns towards end of life, we will adhere to the MOREcare guidance on developing and testing complex interventions in palliative care [35; 36; 37; 38; 39; 40].

### Study design

We present here the protocol and ethics application for the 1st Phase of the study. The larger study will aim to Phase 1: develop, Phase 2: pilot and Phase 3: test an integrated palliative care intervention for COPD patients attending palliative care. Phases 2 and 3 will be the subject of new ethical review applications.

The study will apply the MRC Framework for the development and testing of complex interventions [41].

### Aims & objectives

**Aim:** To develop a feasible and acceptable primary care-led integrated palliative care intervention for patients and families living with COPD

The study objectives are as follows:

PHASE 1 (MRC "DEVELOPMENT" PHASE) Months 1-12

i) OBJECTIVE: To appraise existing interventions for COPD palliative care identifying core components, processes and outcomes through review of the literature.

OUTPUT: Summary of key intervention features to inform qualitative semi-structured topic guide, and covariates for main trial

ii) OBJECTIVE: To appraise the availability of routine acute & primary care health service usage by COPD patients in the Western Cape

*OUTPUT:* Analysis of service usage data *(Publication # 1)* and appraisal of potential for use of routine data for longer term trial outcomes.

iii) OBJECTIVE: To typologise primary care settings in the Western Cape.

OUTPUT: Tabulated list of primary care settings by key characteristics to informing sampling for the development qualitative phase and stratification for the evaluation.

iv) OBJECTIVE: To measure the prevalence of symptoms and concerns among COPD patients attending primary care.

OUTPUT: Prevalence study to inform the clinical intervention development and trial sample size (Publication # 2).

v) OBJECTIVE: To describe stakeholder views (patients, staff, primary care health care professionals) on symptoms and concerns, their current and preferred methods of management, components and delivery of intervention, and staff training and mentorship needs.

OUTPUT: Primary data on stakeholder views to inform feasibility and acceptability, and ensure the theoretically-driven person-centred intervention is generated from patient preferences and priorities (Publication # 3).

vi) OBJECTIVE: To integrate findings from objectives i-iv to generate a novel intervention underpinned by the theory of person-centredness and a model of intended action.

OUTPUT: a theoretically-driven evidence-based person-centred intervention with optimal potential for effectiveness across diverse primary care settings (Publication # 4).

#### Phase 1 Target population

People living with COPD in the City of Cape Town. We support the conclusion of a review of palliative care for people with COPD that "early and longitudinal palliative care takes place in the outpatient setting and is initiated by the patient's primary care physician" [42].

#### Setting

The study will take place in 8 sites: TWO primary care district hospitals: False Bay Hospital and Wesfleur Hospital, THREE 24hr Community health centres with Emergency units from different sub-structures in the Cape Metro, proposed CHCs are Delft, Gugulethu and Vanguard CHCs; and THREE 8hr Community Health Centres with emergency rooms from different sub-structures in the Cape Metro, proposed CHCs are Durbanville or Goodwood, Nyanga or Heideveld, Du Noon or Grassy Park. We will be guided by the Department of Health in finalising these sites.

#### Inclusion and exclusion criteria

##### *Inclusion criteria*

*Patients*: Adults (aged at least 18 years) attending primary care with diagnosed COPD

Able to communicate in English, Afrikaans or Xhosa

Able to give informed consent

*Caregivers:* Primary caregiver to be identified by the patient, in line with the definition of caregiver "unpaid, informal providers of one or more physical, social, practical and emotional tasks. In terms of their relationship to the patient, they may be a friend, partner, ex-partner, sibling, parent, child or other blood or non-blood relative."[43]

Adult

Able to communicate in English, Afrikaans or Xhosa

Able to give informed consent

*Staff:* Health care professionals (doctors, nurses, social workers, occupational therapists, physical therapist) who see COPD patients in the primary care setting as part of their routine practice.

#### Exclusion criteria

Patients: Housebound and unable to attend primary care

### Phase 1 recruitment procedure

Recruitment will take place by dedicated study research staff. Eligible patients and families will be identified by their treating clinical teams and have the study introduced to them. If they are interesting in potential participation, then they will be introduced to the study researcher who will provide further study details. If they wish to participate they will be given 24 hours to consider their decision and then arrange to meet the researcher at the clinic or at their home (taking research staff safety into consideration).

Staff will be directly approached by the study researcher and follow the same procedure.

Questionnaires, information sheets and consent forms will be translated into Afrikaans and Xhosa using forward/backward translation to ensure consistency. Research staff will be bi/trilingual and able to collect data in any of the three study languages within the team.

##### Consent

Participants will have the information sheet read aloud to them and once they have had the opportunity to answer any questions, they will be asked to sign (or give an ink thumb print) on the consent form. They will be given a copy of the information sheet to keep.

#### Potential harms and benefits

##### Potential harms

This phase of the study is cross-sectional and has no intervention. Potential harms are that the interviews will ask that the participant discuss their symptoms and concerns and the care they receive. This has the potential for distress.

##### Potential benefits

Studies have shown that research interviews with palliative care patients are very rarely distressing and that in fact participants report benefit from 1) being able to discuss their illness and concerns and 2) the sense that they are making a contribution to others beyond their own life and care who will have need for advanced care in the future[44].

##### Mitigation of potential harms

We are two clinical academic departments of palliative care (King's College London and University of Cape Town) with a long track record of conducting palliative care research. Firstly, none of our study materials (topic guides or questionnaire items) disclose diagnosis or prognosis to patients or families. All participants will be previously diagnosed with COPD and we will not share any information regarding future disease development.

Our distress protocol is as follows.

Before data collection begins, the participant will be informed that the information they provide will not be shared in any identifiable way, however, if they share any information which causes the researcher to be concerned for the patient's welfare or the welfare of another, they will share that information with the participant's clinical team, and will tell the participant that they are doing so.

Any participant who appears to become physically or psychologically will have their interview paused. The interviewee can then choose to resume if appropriate (e.g. coughing episode has passed or they have repositioned to be more comfortable), to take a rest and resume, to make a further appointment to continue the interview, or to terminate and end participation. At the end of the interview, participants will be offered time with the researcher to debrief, with no data being collected during this debrief. If any concerns (pain, psychological distress, lack of information) arise during the interview, the researcher will offer to refer the patient back to their clinical team.

### Data storage and management

Data will be stored as follows. All paper-based data will be saved in a locked filing cabinet in a locked research office within the Dept of Public Health & Family Medicine at UCT. This Dept is only accessible by swipe card/ door entry following verbal request by the continually present dept administrator during office hours.

Consent forms, patient name/contact details and completed data collections tools will all be stored separately. Questionnaires will only carry the unique Study ID number (no patient name, date of birth, contact details). Electronic data will be stored on a study-specific PC, again in the locked research office within the Dept of Public Health & Family Medicine at UCT. The PC will be password protected. Qualitative digital data will be stored on the PC and then deleted following transcription. Transcription will remove any identifying information (names of patients, families, staff, clinics/providers)

Paper based data will be stored for a period of 7 years.

Data transfer between UCT and KCL will be of Epi Data files of anonymised quantitative data, and Word files of anonymised qualitative transcripts following Material Transfer Agreement.

Reporting of qualitative data will remove identifying data while meeting the aims of the research, with the understanding that some patient stories may be identifiable by staff.

Participants will have the right to remove all their data from the study within 2 weeks of the data collection.

We will directly observe recruitment and data collection procedures monthly. All data will be double entered and discrepancies resolved through reference to the original forms.

### Procedure

#### **OBJECTIVE ii): To appraise the availability of routine acute & primary care health service usage by COPD patients in the Western Cape.**

*Design*: analysis of routine anonymised data.

*Procedure*: The anonymised acute health service use database is held by UCT for analysis. We will liaise with the PI of this dataset (based within our Dept of Public Health & Family Medicine) to extract data on the demographics, clinical characteristics and admissions, length of stay for the past 5 years. Data analysis will be at KCL following transfer of anonymised dataset.

#### **OBJECTIVE iii): To typologise primary care settings in the Western Cape.**

*Design*: self-report cross-sectional interview with facility manager using pro-forma

*Procedure*: The facility manager at each site (Community Health Clinics and District Hospitals) will be approached by the study dedicated researcher to participate in a single interview. The interview will be conducted at the Facility at a time of convenience to the manager. The researcher will work through the pro-forma with the manager and complete each descriptive variable to understand the current care for COPD patients (see Appendices).

*Analysis*: Data will be extracted into a Notedpurpose-designed Excel sheet to allow comparison across facilities.

#### OBJECTIVE iv): To measure the prevalence of symptoms and concerns among COPD patients and family members attending primary care.

*Design:* Cross sectional survey with self-report, file extraction and FEV1

*Procedure:* We will recruit a convenience sample of COPD patients within the sample of clinic, with the sample weighted according to facility size. The sample size has been calculated to estimate prevalence of problems in an unknown population size with 5% precision and 95% confidence. Sample n=385. We will record the response rate.

*Measures*: are as follows, with each read aloud and the study researcher recording answers. Patients will be interviewed alone but may choose to have a family member present is needed.

Socio-demographic data (age, sex, ethnicity, first language, education level, living arrangements and socioeconomic status) will be collected from the patients. Clinical data will be collected from patient records including diagnosis, years since diagnosis, stage of the illness, current treatment, medication adherence, referral patterns and referral to other services performance rating and co-morbidity. We will also extract from file the patient health status/disease stage at the point of referral from primary care, number of re-referrals, number of hospitalizations in previous 12 months and number of healthcare contacts in previous 2 months (outpatient visits, telephone contacts etc), date/result of last FEV1 on record, and then the researcher will measure the patient's FEV1(see Appendices).

Patients will be requested to fill in the Memorial Symptom Assessment Scale- Short Form [45; 46] (MSAS-SF) every month. The MSAS-SF is an abbreviated version of the MSAS, a questionnaire validated in several different populations including patient with heart failure [47; 48]. The MSAS-SF enables a multidimensional assessment of symptoms. Patients are asked to rate symptom presence and symptom burden for 32 symptoms. Answers are given on a five-point Likert scale ('not at all', 'a little bit', 'somewhat', 'quite a bit', or 'very much').

Patients and family members will also complete the 10 item African Palliative Care Association African Palliative Outcome Scale [49]. This instrument measures the multidimensional wellbeing of patients. The Palliative Outcome Scale (POS) was originally developed by researchers in the Department of Palliative care and Policy, King's College London[50]. The POS was then adapted for use in Africa by the African Palliative Care Association (APCA) in 2007[51]. It has been validated for use in South Africa, in English, Afrikaans and Xhosa, Zulu and Sotho[49].

Patients will be requested to complete the Center for Epidemiologic Studies Depression Scale (CES-D). The CES-D is a common screening scale measuring feelings and behaviours characteristic of symptoms of depression during the past week. Scores range from 0 to 60, and 16+ used as a cut-off for probable caseness[52]. It has been used in previous community depression studies in South Africa[53; 54; 55]. It has a Chronbach's Alpha of 0.90 for women and 0.91 for men.

Patients will also complete The Medical Outcomes Study (MOS) Social Support Survey[56]. The questionnaire measures the participants' levels and type of social support. The survey has 19 items that are further divided into 4 subscales, namely emotional/informational supports, tangible support; affectionate support and positive social support, and one additional global item. The instrument has been used in several heart failure studies and also in studies in (South) Africa [57; 58; 59]. The reliability has been established ($\alpha > 0.91$) and has been stable over time.

Severity of breathlessness on exertion over the previous 24 hours will be measured using the London Chest Activity of Daily Living Questionnaire (15 items)[61].

The COPD Assessment Test (CAT) is a simple, eight item, health status instrument for patients with COPD [62], which is highly practical and has good psychometric properties including responsiveness[63]. A decrease in CAT score represents an improvement in health status, whereas an increase in CAT score represents a worsening in health status. The most reliable estimate of the minimum important difference of the CAT is 2 points[63].

The Client Services Receipt Inventory is a measure of health service and informal care usage[64], which we have used in the African context[65; 66].

*Analysis:* For all measures we will conduct descriptive analysis to report prevalent concerns

determine the normality of data and then conduct regression analyses (linear, ordinal as appropriate) and identify predictors of scores for each measure integrate the prevalence data from this objective with the qualitative data from the subsequent objective to determine the primary and secondary outcomes of the intervention.

#### OBJECTIVE v): To describe stakeholder views (patients, staff, primary care health care professionals) on symptoms and concerns, management and training needs

*Design:* Cross-sectional in-depth qualitative interviews with a purposive sample of stakeholders.

*Procedure*: A purposive sampling frame with be used for each stakeholder group.

For patients: ≤25 adult patients who meet the inclusion/exclusion criteria sampled according to age, gender, ethnicity, family size, disease severity, frequency of admissions.

Topic guide: primary needs and concerns, what would make care more patient centred and how should that be delivered, current self-management, preferences for advance planning and communication practices, views on potential intervention components, decision making and drivers of emergency unit (EU) attendance.

For family members: ≤20 family primary caregivers as identified by the patient who meet the inclusion/exclusion criteria sampled according to age, gender, ethnicity, relationship to patient, family size, patient disease severity, frequency of admissions.

Topic guide: primary needs and concerns, what would make care more patient centred and how should that be delivered, current management strategies for patient, preferences for discussion regarding advance planning and communication practices and involvement in that process, views on potential intervention components, decision making and drivers of EU attendance.

For staff: ≤ 20 Health care professionals purposively sampled according to designation (medicine, nursing, allied health professionals), age, gender.

Topic guide: value of patient-centredness, barriers and training/mentorship needs, leadership requirements, implementation, communication skills needs, anticipated challenges and benefits

*Analysis:* interviews will be transcribed verbatim and then translated into English. Translated transcripts will be imported into NVIVO for thematic analysis.

#### OBJECTIVE vi): To integrate findings form objectives i-iv to generate a novel intervention underpinned by the theory of person-centredness and a model of intended action.

Procedure: we will convene an expert meeting pf patients, family members, primary care, respiratory, and palliative care, academics. The meeting will review the findings from objective i-vi to develop and feasible and evidence-based intervention.

We will utilise the primary data from objectives i-iv, and draw on the following:

The theory of person-centredness to achieve the goal concluded by a review of family and patient preferences in advanced COPD- that "*care interventions need to be based on patient preferences rather than professionally driven*"[4].

A systematic review of interventions (pharma & non-pharma) to manage breathlessness [29]

Our Decision Support Tool recommendations of how to manage self-report breathlessness scores (based on international Delphi and systematic review)

Prior trial evidence that non-malignant patients with breathlessness value professional reassurance of their long-standing strategies [26].

### Dissemination of research findings

A report will be written describing research findings and disseminated to facility managers, the department of health in the W Cape and City of Cape Town. A summary of the findings will be developed in leaflet format for distribution to participants and a poster format will be developed for display at facilities.

Publications will be written for submission to peer-reviewed journals and abstracts submitted to relevant conferences for oral and/or poster presentation.

### SOUTH AFRICA: WORK PACKAGE 6

#### Maternal Mental Health and Violence against Women in South Africa

#### Purpose of the study

The purpose of this study is to improve the coverage of maternal mental health services in the Cape.

This purpose will be served through the following specific objectives:

1. To support the Western Cape Department of Health (WCDoH) in the scaling up of a screening and counseling service for PCMD, linked to the roll out of the Practical Approach to Care Kit (PACK) guidelines for routine antenatal care.
2. To collaborate with local stakeholders to develop and pilot an intervention for care and support for victims of violence against women (VAW) during the perinatal period, integrated into routine perinatal healthcare, and linked to the scaled up counseling service in Objective 1.

#### Background

**Perinatal common mental disorders (PCMD)** such as antenatal and postnatal depression and anxiety are highly prevalent in the Western Cape, South Africa. Studies in Khayelitsha, a large peri-urban township settlement in Cape Town have reported a point prevalence of depression of 39% in pregnant women,[1] and 34.7% in postpartum women.[2] Perinatal depression in this setting is associated with lack of partner support, intimate partner violence, low household income, and younger age.[1,2] Perinatal anxiety disorder has been reported in 23% of antenatal women in one deprived urban setting in Cape Town, and was associated with a history of pervious mental health problems, major depressive disorder, multigravidity, food insecurity, unplanned or unwanted pregnancy, pregnancy loss and experience of threatening life events.[3]

Perinatal common mental disorders have been associated with a number of adverse consequences, for both mother and child. These include preterm birth, low birthweight, diminished mother-infant bonding, infant under-nutrition, stunting and increased prevalence of diarrhoea.[2,4]

While maternal and child health have been identified as key priorities for intervention by the South African Department of Health,[5] no programmes for the treatment of PCMD have as yet been introduced in a systematic manner within the public health sector.[6]

Several models have been developed to provide screening and mental health care for the perinatal period in the Western Cape. The Perinatal Mental Health Project (PMHP), until recently provided services in 3 Midwife Obstetrics Units (MOUs) in the Cape Town metro.[6] The Africa Focus on Intervention Research for Mental health (AFFIRM) randomized controlled trial developed and evaluated a structured manualised task sharing counseling intervention for antenatal depression at 2 MOUs in Khayelitsha, Cape Town in 2014-2016, delivered by community health workers (CHWs).[7] The trial showed that there was a small but significant effect of the counselling intervention on depression at 3 months postnatal, compared to enhanced usual care (3 supportive monthly phone calls). The effect became more significant at 12 months postnatal, suggesting that the counseling intervention has longer term benefits on managing depressive symptoms compared to the phone calls. Women in both intervention and control arms showed a substantial reduction in depression symptoms into the postnatal period.

Following presentation of the preliminary AFFIRM trial results in January 2017, senior Managers in the WCDoH were supportive of scaling up a counseling service for antenatal depression in all 12 MOUs in the Cape Town metropolitan area. The WCDoH is willing to work with the UCT ASSET study, who would take responsibility for the development of training materials and the evaluation of the scale up.

**Interpersonal violence** is ranked 2nd in its contribution to the burden of disease in South Africa, and among women, intimate partner violence (IPV) accounts for 62.4% of the burden of interpersonal violence.[8] The Western Cape gender based violence indicators study reported in 2014 that 39% of women had experienced some form of violence in their lifetime, and approximately the same proportion of men report perpetrating violence against women.[9] In the same study, 13% of women in the Western Cape reported physical or sexual abuse during at least one of their pregnancies. These are likely to be under-estimates, as many women do not disclose their experience of violence.

Currently there is no routine screening for gender-based violence among women during antenatal or postnatal care visits, either in WCDoH services or in PACK 2016-2017. Recognition of victims of intimate partner violence (IPV) in primary care in the Western Cape is very low – in one study only 9.6% of victims of IPV were recognized in primary care.[10] This study did not recommend universal screening for VAW in the perinatal period, but recommended indicated screening, particularly for women who present with the following signs: sexually transmitted infections including HIV, assault, chronic pain syndromes and symptoms suggestive of a mental health problem e.g. sleep disturbance, tiredness, feeling depressed or anxious; history of a mental illness or psychiatric medication. Models developed to date have recommended indicated screening, conducted by a primary care provider, with referral to an 'IPV champion' who will provide a more comprehensive assessment and management plan.[11]

### Methods

#### Study design

The study will include three phases, with different study designs for each phase.

Phase 1 (Diagnostic phase): Situation analysis, Formative qualitative research, Theory of Change workshops and Literature review.

Phase 2 (Piloting phase): Pilot testing intervention and assessment instruments.

Phase 3 (Evaluation phase): Controlled before-after study, supplemented by qualitative and quantitative process evaluation.

### Phase 1: Diagnostic Phase

#### Characteristics of the study population

The following study participants will be recruited:

1. Approximately 40 pregnant women attending their first antenatal clinic booking at Midwife Obstetric Units and mothers of young babies attending well baby clinics in Cape Town. Inclusion criteria will be: screen positive for either depression/anxiety (score >13 on EPDS) or experience of violence in the past 3 months, resident in Cape Town, age 18 years and older, and capacity to provide informed consent.
2. Approximately 20 Community Health workers (CHWs) and Lay Counselors who work in the catchment area of MOUs or Community Health Centres in Cape Town. This will include CHWs and lay counsellors

within Department of Health structures, and employed in NGOs subsidized by the Department of Health.
3. Approximately 10 Registered Nurse Midwives and nurses who work at MOUs in Cape Town.
4. Approximately 8 Service managers at MOUs and Community Health Centres and District Health Management staff in Cape Town.
5. Approximately 10 social workers linked to Department of Health services and in the Department of Social Development.
6. Approximately 10 management staff involved in the management of non-governmental organisations (NGOs) providing support for mental health needs or gender based violence for perinatal women in Cape Town.

Inclusion criteria for groups 2-6 will include professional roles as described, and capacity to provide informed consent.

Participants from Department of Health facilities will be recruited from one MOU in each of the four substructures. Specific facilities will be nominated by our Department of Health senior management colleagues, in order to represent each of the four substructures in the Cape Town metro (See Appendix for list of all MOUs in the Cape Town metro). Sample sizes are approximate as recruitment will continue until saturation is achieved for each of the major themes to be explored.

#### Recruitment and enrolment

For phase 1, the following recruitment and enrolment procedures will be followed:

1. For pregnant women attending their first antenatal booking at Midwife Obstetric Units and mothers of young babies (below 1 year of age) attending well baby clinics in Cape Town: we will use purposive sampling to get a roughly equal representation of women who meet criteria for depression/anxiety and experiences of violence, and an equal representation from antenatal and well baby clinics. These women will be approached in the queues at the clinics and asked if they would be willing to participate in the study (See Appendix for Informed Consent form for perinatal women). If consent is provided the Edinburgh Postnatal Depression Screening (EPDS) tool and the violence screening tool will be administered (See Appendix). Women who score 13 or above on the EPDS or indicate experience of physical or sexual violence during the past 3 months will then be invited to participate in a semi-structured interview, which will last between 45 minutes and 1 hour (See Appendix for SSI Schedule for perinatal women). In addition, we will also ask these women about their experience of care and needs for person centred healthcare (See Appendix).
2. For Community Health workers and Lay Counselors who work in the catchment area of MOUs or Community Health Centres in Cape Town: we will approach these individuals at their work place and ask them about their willingness to participate in the study (See Appendix for Informed consent form for Health Workers). Those who consent to participate will take part in a semi-structured interview that will last between 45 minutes and 1 hour (See Appendix for SSI schedule for counselors).
3. For the Registered Nurse Midwives who work at MOUs in Cape Town: we will approach these individuals at their work place and ask them about their willingness to participate in the study (See Appendix for Informed consent form for Health Workers). Those who consent to participate will take part in a semi-structured interview that will last between 45 minutes and 1 hour (See Appendix for SSI schedule for Midwives).
4. For Service managers at MOUs and Community Health Centres and District Health management staff in Cape Town: we will approach these individuals at their work place and ask them about their willingness to participate in the study (See Appendix for Informed consent form for Health Workers). Those who consent to participate will take part in a semi-structured interview that will last between 45 minutes and 1 hour (See Appendix for SSI schedule for Service managers).
5. For Social workers in the Department of Social Development: we will approach these individuals at their work place and ask them about their willingness to participate in the study (See Appendix for

Informed consent form for Social Workers). Those who consent to participate will take part in a semi-structured interview that will last between 45 minutes and 1 hour (See Appendix for SSI schedule for Service managers).

6. For staff involved in the management of non-governmental organisations (NGOs) providing support for mental health needs or gender based violence for perinatal women in Cape Town: we will approach these individuals at their work place and ask them about their willingness to participate in the study (See Appendix for Informed consent form for NGOs). Those who consent to participate will take part in a semi-structured interview that will last between 45 minutes and 1 hour (See Appendix for SSI schedule for NGOs).

In addition, after the completion of the initial situation analysis and semi-structured interviews, we will recruit participants for Theory of Change workshops in each of four MOUs, comprising staff involved in the provision of care for perinatal women (see below).

#### Research procedures and data collection methods

##### *Situation analysis*

We will develop a situation analysis tool that compiles data from four selected MOUs (one per substructure in the Cape Town metro, as stated above). Data will be collected on healthcare worker numbers and roles in each facility; skills and competencies; governance, organization and leadership of care processes; patient flow through the care system; availability and functionality of essential equipment; availability of essential drugs; functioning of HMIS; and feasibility of using patient records as a data source.

We will review existing clinic records of 4 selected MOUs (one per substructure), to collect data on numbers of antenatal women presenting for their first clinic assessment per month. Data will include mean numbers of antenatal and postnatal clinic visits; records of depression or violence (if any). We have estimates of numbers of depression screen positive women from our own previous research (AFFIRM trial) as well as data from the Perinatal Mental Health Project (in 3 MOUs) and other studies. Data will also be obtained from previous studies of gender-based violence during the perinatal period to determine prevalence estimates and risk factors.

The situation analysis tool will be based on a prior situation analysis tool that we developed for the Programme for Improving Mental health carE (PRIME).[12] The tool will elicit data that is currently available in routine health management information systems or in publically available databases. Quantitative data for the situation analysis will be collected by fieldworkers using Computer Assisted Personal Interviews (CAPI).

Data from the situation analysis will be used to map current resources and systems of care for perinatal women with depression, anxiety and experiences of violence in MOUs in the Cape Town metro. This will be important for considering the possible design of a referral and counseling service integrated into existing healthcare systems, in the piloting and evaluation phase.

##### *Formative qualitative research*

The situation analysis will be supplemented with qualitative interviews with facility management, care providers, and perinatal women, as described above, exploring these issues. Qualitative semi-structured interviews will be conducted regarding experiences of depression, anxiety and violence, coping strategies and feasibility and acceptability of the proposed intervention. The focus of the qualitative research is therefore on the acceptability and feasibility of a service, rather than the development of a new psychological intervention, as the intervention will be an evidence-based problem-solving therapy intervention, adapted from the AFFIRM RCT.[13] In relation to CHWs and lay counsellors, one of the main questions of the diagnostic phase is who would be best placed to deliver the counseling (CHWs or HIV/lay counselors), and where is the best place to deliver it. We will also conduct semi-structured interviews with District health management staff regarding their perceptions of current perinatal health services, perinatal mental health services, needs of women who are depressed, anxious or are victims of violence, and feasibility and acceptability of the proposed intervention.

Interviews will be conducted in English, Afrikaans or isiXhosa, depending on the preference of the study participant.

The ASSET Fieldwork coordinator and trainer/supervisor will also conduct observations of care processes in the four selected facilities (one in each substructure). This will include observations of the stages along the care pathway from antenatal booking/well baby clinic to referral to treatment to outcomes, exploring a variety of scenarios. It will also include observation of liaison with other sectors such as Social Development, statutory services and NGOs, as relevant.

##### *Theory of Change workshops*

We plan to conduct Theory of Change workshops in the diagnostic phase, as we believe that they are an essential means of collecting data, obtaining buy-in and informing both the content and evaluation of the intervention. These ToC workshops, as described above will happen at 3 levels:

1. Senior management (1 ToC workshop)
2. Facility management and staff (4 ToC workshops, one in each facility)
3. NGOs and Department of Social Development (1 ToC workshop)
4. Service users: perinatal women with depression/anxiety and experiences of violence (2 ToC workshops)

The objectives of the ToC workshops will be to obtain the suggestions of these key stakeholder groups regarding the design, content, feasibility and acceptability of the proposed intervention, and to promote their buy-in.

During the diagnostic phase we will also establish a Community Advisory Board, which will include local community organisations working in the field of perinatal health, the Department of Health's "First 1000 days" initiative, and NGOs working with women who are victims of violence in the community.

We will use the findings of the diagnostic phase to develop a draft proposal and theory of change for the intervention. We will then present the proposal and obtain feedback from provincial policy makers/senior management; facility managers and staff; and patients to obtain their suggestions regarding the design, content, feasibility and acceptability of the proposed intervention, and to promote their buy-in.

##### *Literature review*

We will conduct a structured literature review to identify optimal screening tools to be used by midwives or relevant MOU personnel to routinely detect women with depression, anxiety and experiences of violence in antenatal clinic visits.

#### **Data safety and monitoring**

Qualitative data collected from in-depth interviews will be recorded and transcribed verbatim into MS Word files before being uploaded into NVivo 11 for analysis. Quantitative data will be collected through fieldworkers by administered Computer Assisted Personal Interviews (CAPI). We will have verbal and written permission from participants to collect all types of data.

Data quality and standards assurance will occur during data collection and during data management. We will initiate a project-specific data transfer system and protocol to regularly transmit data which will be organized into a single database with unique project-generated identification numbers. A link file approach will be used for maintaining the confidentiality of data files. In a link file system, confidential information is separated into different files without common links. A third link file that contains identifying number pairs is used to link the data records. The advantage of this system is that either of the database files can be used directly for report generation without the use of decrypting subroutines or access to the link file. The study database will be stored within the firewall- protected UCT server and nightly backups will be performed. The study database will be password-protected following standard password safety procedures. It is UCT's policy that the fewer the number of individuals handling sensitive information, the greater the protection. Therefore, project files and databases associated with this study will only be available to research personnel through the authorization of the PI, and all staff with access to the project data must sign confidentiality agreements. These agreements will cover information on what is and is not allowed to be shared, what data can be shared, and the specific permissions required before data or information may be shared. In addition, the study reports (e.g., aggregated data in progress reports) generated by the research team will provide total anonymity to participants since no names or identifying data will be part of such reports. Paper copies of locator information, consent forms, and participant contact forms linked to the data through the project-generated identification will be stored in locked filing cabinets in a locked office at UCT. Participant locator information will be destroyed within 1 year of the conclusion of the study.

#### Data analysis

For the initial diagnostic phase data will be largely qualitative. Interviews conducted in Afrikaans and isiXhosa will be translated into English before analysis is conducted. Analysis of transcribed interview recordings will be conducted in English using the framework approach with NVivo qualitative data analysis software.[14] The data will be indexed and initial codes will be guided by the semi-structured interview topics. The analysis will focus specifically on discovering themes related to the main topics of enquiry for each stakeholder group (see Semi-structured interview schedules for the main themes). Further emergent themes not captured by the initial coding will be identified through extensive reading of the transcripts. Once a coding framework is agreed by the investigating team, two independent coders will commence with coding. They will consult regularly to compare coding and resolve discrepancies. Cohen's Kappa will be calculated to assess inter-rater reliability, and a standard 80% agreement will be regarded as acceptable.

Quantitative data collected from the situation analysis tool will be analysed descriptively. The quality of routine health management information systems (HMIS) data will also be assessed for its face validity and reliability, specifically with a view to potentially using routine HMIS for the process evaluation during the evaluation phase.

#### Description of risks and benefits

Some study participants, specifically perinatal women with experiences of depression, anxiety or violence, are a highly vulnerable group who may require specific support or interventions. For example women with severe depression or anxiety may be at risk of suicide, and women who report being victims of violence may well be in ongoing danger from the perpetrator. In order to assess this risk, specific questions regarding suicidal thoughts and risk of harm by others will be included in the initial interview. In the event that a participant reports intent of self harm, she will be referred to the psychiatric nurse in the facility for further assessment. In the event that a participant reports that she is in danger from a perpetrator of violence, she will be referred to a social worker and will follow statutory protocols for her protection. Depending on the severity of the situation this may include provision of safe housing. We will check that referral services are adequately equipped to manage these referrals, and will facilitate support where necessary. A referral form will be created, and a list of organizations working in the area, together with their contact details will be provided to all women who are at risk of interpersonal violence or self-harm.

During training for the conduct of semi-structured interviews, fieldworkers will be briefed on standard protocols for these referral procedures, and will be trained in the sensitive and supportive enquiry of the study participant's needs and wishes.

Although some study participants are a vulnerable group, the risks of harm associated with phase 1 of this study are minimal. This is because the study will not involve any intervention, and interviews will be conducted strictly for the purpose of eliciting participants' views on their experiences and service needs. No new interventions will be introduced or evaluated in this phase. Our experience from previous studies has shown that if interviews are conducted in a sensitive manner, women in these circumstances report finding the interviews supportive and helpful.

#### Informed consent process

Informed consent will follow standard procedures, as set out in the attached Informed Consent form (see Appendix). Fieldworkers will be trained to assess the capacity of the participant to provide informed consent, and those who are found to be lacking such capacity will be excluded from the study.

The process will be that the field workers will individually approach potential participants in the waiting room for their antenatal first bookings at the MOU, and individually request their consent to participate. The fieldworker will inform potential participants that they may refuse to participate at any stage and will not be disadvantaged in any way if they do, and their normal standard of health care will not be affected if they refuse. Participants will be allowed to withdraw from the interview process at any stage. Participants will be asked if they have any questions and if they need more time to consider their options. The midwives will have the fieldworker's phone number at hand if they need to ask any questions themselves.

Health workers, service managers, social workers and NGO management staff will be approached individually either face to face in the facility, or telephonically, to ask if they would be willing to participate in the study.

If participants agree to participate, they will sign an informed consent form available in English, Afrikaans and isiXhosa. A printed information sheet (available in English, Afrikaans and isiXhosa) about the study will also be given to all the participants.

#### Privacy and confidentiality

Strict measures will be put in place to ensure that the privacy of all study participants is protected and confidentiality of all information is maintained, as set out in the above data management protocol. In addition to the removal of personal identifying information, special care will be taken to remove information identifying specific facilities, which may be linked back to health care or health management staff.

#### Reimbursement for participation

Perinatal women will be reimbursed with a grocery voucher to the value of R100 for participating in the study. Health service, social development and NGO staff will not receive any reimbursement for their time as participating in the study is aligned with their professional duties.

#### Emergency care and insurance for research-related injuries

Although the study poses minimal risk of injury as it will largely involve semi-structured interviews and workshops, UCT No-fault insurance will be taken out to protect participants in the event of research related injuries.

#### What happens at the end of the study?

At the end of this diagnostic phase of the study (end of Year 1), the findings will be analysed and written up for publication in peer reviewed journal articles. Findings will be used to inform the design of the main health system strengthening intervention, to be piloted in Phase 2, and evaluated in Phase 3. At that stage we will apply to the UCT Faculty of Health Sciences Human Research Ethics Committee for an amendment to the protocol, and submit the detailed ethics protocols for Phases 2 (piloting) and 3 (evaluation).

Currently the content of these protocols cannot be determined as they will be contingent on the findings of the diagnostic phase. Nevertheless they are likely to include the following aspects:

Study design: Controlled before and after evaluation, supplemented by qualitative and quantitative process evaluation.

Intervention: enhanced PACK, comprising further training of midwives in awareness and detection of depression/anxiety and violence, structured record keeping and referral pathways, manualised telephone or digital platform counseling service plus referral to face-to-face counseling for severely depressed/anxious women at risk of self-harm or referral of at-risk women who are victims of violence for statutory protection.

Primary outcome will be the proportion of women showing a clinically significant improvement on the PHQ9, at 6 months follow-up (defined as a 50% reduction in PHQ9 score from baseline).[15] Data source: individual follow-up by ASSET employed fieldworkers.

Secondary outcomes:

1. Change in treatment contact coverage (counseling) for antenatally depressed/anxious women. Data source: repeat facility detection surveys.
2. Change in detection of antenatal depression/anxiety at MOUs. Data source: repeat facility detection surveys.
3. Change in referral of women with antenatal depression/anxiety at MOUs. Data source: Routine HMIS (depending on feasibility assessment in diagnostic phase)
4. Individual functioning outcomes for women, measured using WHODAS and a previously validated locally developed functioning assessment instrument for perinatal depression.[16] Data source: individual follow-up using ASSET employed fieldworkers.
5. Change in detection of victims of VAW. Data source: repeat facility detection surveys.
6. Change in referral of victims of VAW. Data source: Routine HMIS (depending on feasibility assessment in diagnostic phase)
7. Proportion of women self-reporting as safe. Data source: individual follow-up using ASSET employed fieldworkers.

The following care processes will be important to evaluate:

1. Treatment contact coverage (counseling) for antenatally depressed/anxious women.
2. Detection of antenatal depression/anxiety at MOUs.
3. Referral of women with antenatal depression/anxiety at MOUs.
4. Detection of victims of VAW.
5. Referral of victims of VAW.

The following indicators will be linked to each of the above care processes:

1. Change in treatment contact coverage (counseling) for antenatally depressed/anxious women.
2. Change in detection of antenatal depression/anxiety at MOUs.
3. Change in referral of women with antenatal depression/anxiety at MOUs.
4. Individual functioning outcomes for women, measured using WHODAS.
5. Change in detection of victims of VAW.
6. Change in referral of victims of VAW.
7. Proportion of women self-reporting as safe.

Sample size calculation: to determine the number of observations required per cluster, for a two sample comparison of proportions (using normal approximations) without continuity correction. Assumption: that introduction of screening, referral pathways and counseling will lead to a clinically significant improvement on the PHQ9 (defined as 50% reduction in PHQ9 scores compared to baseline); that intervention clinics will show a 55% clinically significant improvement, compared to 30% in control clinics; significance of 0.05; 80% power; clusters available per arm: 6; cluster coefficient of variation: 0.25. Average cluster required 30 (180 per arm). To allow for loss to follow-up: 40 per cluster, 240 per arm, total N=480.

#### Timeline

Phase 1: Diagnostic phase (April-December 2018):

1. Situation analysis, qualitative formative research and consultation with local stakeholders on design of the intervention, including Theory of Change[17] workshops.
2. Adaptation of existing counseling manuals and training and supervision materials (including translation into Xhosa and Afrikaans).
3. Planning with DoH and NGO partners for Scale up and development of Standard Operating Procedures (SOPs).
4.

Phase 2: Piloting (January – June 2019):

1. Finalising of counseling manuals and instrumentation.
2. Piloting of intervention in one facility.

Phase 3: Evaluation (July 2019 – March 2021)

1. Baseline data collection for primary and secondary outcomes in all 12 MOUs in Cape Town metro (including enrolment of cohort).
3. Roll out of train the trainer programme and subsequent training and ongoing supervision of counselors in NGOs, linked to 6 MOUs (intervention sites).
4. Follow-up data collection for primary and secondary outcomes in all 12 MOUs. This would include enrolment and follow-up of a cohort of women from the 6 intervention MOUs.
5. Qualitative process evaluation, comprising individual key informant interviews and focus group discussions with a range of local stakeholders.
6. Quantitative process evaluation of training and supervision.
7. Subsequent roll out of enhanced PACK intervention, including train the trainer programme to remaining 6 control MOUs.
8. Data analysis and write up.
9. Dissemination, including publication in peer reviewed journals, dissemination to local health forums and community health advocacy groups and engagement with provincial and national policy makers regarding uptake and implementation of study findings and recommendations.

### SIERRA LEONE: WORK PACKAGE 7

#### Surgical Care in Sierra Leone

**Principal investigators**

Dr Andy Leather, Director, King's Centre for Global Health & Health Partnerships, King's College London


Professor Justine Davies, King's Centre for Global Health & Health Partnerships, King's College London


**Lead co-PI (Cross-ASSET Project)**

Professor Martin Prince, Global Health Unit for Health System Strengthening, King's College London


**Co-investigators**

Dr Haja Wurie, College of Medicine & Allied Health Sciences, Sierra Leone


Dr TB Kamara, College of Medicine & Allied Health Sciences, Sierra Leone


Dr Fenella Beynon, King's Sierra Leone Partnership, King's Centre for Global Health & Health Partnerships, King's College London


Dr Daniel Youkee, King's Sierra Leone Partnership, King's Centre for Global Health & Health Partnerships, King's College London


Professor Jennifer Gallagher, Dental Institute, King's College London, UK


Dr Ann H. Kelly, Department of Global Health & Social Medicine, King's College London, UK


Professor Nick Sevdalis, Institute of Psychiatry, King's College London, UK


Dr Chris Willott, King's Centre for Global Health & Health Partnerships, King's College London


Dr Abebe Bekele, Department of Surgery, School of Medicine, College of Health Sciences, AAU, Ethiopia


Dr Charlotte Hanlon, CDT-Africa, College of Health Sciences, Addis Ababa University (AAU), Ethiopia


### Executive summary

#### Background

Health system strengthening is required in order to achieve universal access to safe, affordable and timely surgical and anaesthetic care. In Sierra Leone, the Ministry of Health & Sanitation is formulating a national surgical plan to increase the volume and quality of surgical care. This is in response to the Lancet Commission on Global Surgery and to the World Health Assembly Resolution 68.15 'Strengthening emergency and essential surgical care and anaesthesia as a component of universal health coverage.' To fully realise the potential of these important initiatives and to inform the implementation of the national surgical plan, we need high quality evidence on effective interventions to strengthen the health system.

##### Overall aims

The overall aim of ASSET (health system strengthening in sub-Saharan Africa) in Sierra Leone, is to develop effective health system strengthening interventions to support the translation of clinical evidence into delivery of integrated continuing care at scale across healthcare platforms for surgical (including obstetric surgery) and dental care in Sierra Leone.

#### Specific aims

For surgical/dental care:

(1) To assess the volume of surgical interventions and the quality of care provided.
(2) To identify the health system bottlenecks to delivery of integrated, high-quality continuing care
(3) To adapt, pilot and refine health system strengthening interventions based on (A) defining care pathways, (B) electronic information systems, (C) quality improvement, (D) person-centred care and (E) workforce strengthening.
(4) To implement and evaluate the impact of these health system strengthening interventions upon patient outcomes.

#### Methods

In this protocol, we describe the diagnostic phase of ASSET Sierra Leone.

The diagnostic phase of ASSET will include the following:

(1) Study launch and introductory meetings
(2) Situational analysis
(3) Assessment of readiness to provide care at Peripheral Health Units (primary healthcare level) and at district and teaching hospitals (secondary and tertiary healthcare level)
(4) Sierra Leone referral process mapping and geographical origin (residence) of admitted patients
(5) Hospital patient safety culture survey (across representative secondary and tertiary hospitals)
(6) Hospital clinical pathway process mapping
(7) Hospital documentation study
(8) Hospital data capture studies at tertiary sites (bed occupancy, theatre utilisation, time to emergency theatre)
(9) Patient cohort studies at tertiary sites: perioperative mortality rate (POMR), defined as the all-cause in-hospital mortality rate; cost of care for patients undergoing operative procedures (including direct and in-direct costs of care); patient experiences; and patient reported outcomes
(10) Provider qualitative study

(11) Patient qualitative study (demand, help-seeking, preferences, choices, and perceived barriers)
(12) Diagnostic phase synopsis and Theory of Change workshop/s

During the subsequent piloting phase, health system strengthening interventions will be adapted, piloted and assessed using process data and qualitative studies. In the implementation phase, the impact of adapted health system strengthening interventions upon patient level outcomes and the success of their implementation will be evaluated.

##### Expected results

The diagnostic phase will identify health system bottlenecks to the implementation of timely and good quality surgical care. The output will be context-specific health system strengthening intervention(s) that is(are) ready to pilot across six PHUs, two secondary and one tertiary site in the Western Area. The overall ASSET study will provide rigorous and relevant evidence on effective and feasible health system strengthening interventions to support quality surgical care. This will inform the Sierra Leone national surgical plan implementation.

##### Funding

Funded through a grant from the UK National Institute of Health Research (NIHR) to establish an NIHR Global Health Research Unit on Health System Strengthening in Sub-Saharan Africa at King's College Hospital

##### Keywords

Health system strengthening; implementation research; quality improvement research; mhealth; global surgery

#### Overall ASSET background

##### Introduction

ASSET-Sierra Leone is a collaborative project between College of Medicine & Allied Health Sciences, King's College London (KCL) and the King's Sierra Leone Partnership. The focus of ASSET-Sierra Leone is on the development and evaluation of interventions to strengthen the Sierra Leone health system to support surgical (including obstetric surgery), anaesthesia and dental healthcare.

ASSET Sierra Leone is a component part of a larger ASSET research programme. The overall aim of ASSET is to develop effective health system strengthening interventions to support the translation of clinical evidence into delivery of integrated continuing care at scale across healthcare platforms for NCDs, mental and substance use problems, surgical and dental care, and maternal healthcare in Ethiopia, Zimbabwe, South Africa and Sierra Leone. The general methodological approach will involve (1) a diagnostic phase to identify care pathways, the main health system bottlenecks, acceptability and feasibility of potential system strengthening interventions and development of a district level integrated healthcare plan, (2) a piloting phase, and (3) an implementation and scale-up phase.

In this protocol, we describe the diagnostic phase of ASSET-Sierra Leone.

##### Global surgical system challenges and opportunities in Sierra Leone

The global burden of surgical disease is considerable and response to surgically treatable conditions is considered to be one of the most neglected areas in global health. In a recent Lancet Commission on Global Surgery (LCoGS), an estimated 32.9% of all lives lost in the year 2010 were estimated to have been due to health conditions needing surgical care, resulting from a 90% treatment gap for basic surgical care in lower-income countries. Inequity is high, with the biggest unmet need for surgical care being borne by the poorest sector of society, with an estimated 5 billion people lacking access to quality surgical care, when needed. The prioritisation of surgical care will save lives, promote the welfare of communities and the economic growth of nations.

Trauma is a major global health problem, with injuries accounting for 5 million deaths, which is more deaths than TB, Malaria and HIV combined, with 90% of these deaths occurring in LMICs. In addition, about 270000 women die from complications of pregnancy annually. Many of these injury-related and obstetric-related deaths, as well as deaths from other causes (e.g. abdominal emergencies and congenital anomalies), could be prevented by improved access to surgical care. Beyond mortality, untreated surgical diseases are among the top 15 causes of physical disability worldwide. In the case of disability, surgery is not merely curative, but also prevents the social and economic disparities that accompany untreated disabilities. Like other public health solutions, surgery improves the economic output due to improved population health.

Despite this large surgical burden, surgical services are not being delivered to many of the individuals who need them most despite evidence that surgery is cost effective when compared to other public health interventions, and despite a strong economic argument for global scale up of surgical services. Indeed, until the 1990s, health policy in resource-constrained settings focused sharply on infectious diseases and under-nutrition, especially in children. Surgical capacity was developing in urban areas but was often viewed as a secondary priority that mainly served socioeconomically advantaged people. A framework for global health priority setting has been applied to the global surgical community showing that the community still needs to overcome some challenges, before surgical care can be widely acknowledged and priorities for investment in health systems' surgical capacities can be supported.

In 2015, the Lancet Commission on Global Surgery outlined recommendations to improve access to surgery, globally. To measure progress, six indicators were developed: 1) access to timely essential surgery, 2) specialist surgical workforce density, 3) surgical volume, 4) perioperative mortality, and 5 & 6) protection against impoverishing and catastrophic expenditure. Progress has been made with data collection. In addition, the commission urged all countries to develop national surgical plans. A number of countries have made progress including Zambia and Ethiopia.

In Sierra Lone, a population study done in 2012, using the Surgeons Overseas Surgical Assessment Tool (SOSAS tool), estimated that 25% of the population had a condition which required a surgical consultation and 25% of verbal autopsy deaths were likely due to conditions that required surgery. The need to improve the quality of surgical care in Sierra Leone is thus acute, but there are many challenges to improving access to quality surgical care in the country. The current profile of specialist surgical, anaesthetic and obstetric providers in Sierra Leone shows that there are 0.15 specialist surgeons, anaesthetists and obstetricians (SAOs) per 100,000. This is far below the LCoGS recommendation of between 20-40 trained SAOs per 100,000. There are currently estimated to be 400 operations per 100,000 as compared with the LCoGS recommended number of 5000 per 100,000. Historically, surgical care has not been prioritized in Sierra Leone when compared to other LMICs. However recent commitments to improving access to surgical services for the population include: surgery is covered in the ministry of health Basic package of Essential Health Services (2015-2019); and there has been government support for a National Surgical Planning Process that was launched in 2016. In addition, postgraduate training in surgery, obstetrics and anaesthesia has been strengthened by national events including the passage of the Acts of Parliament to establish the Teaching Hospitals Complex and the Council for local postgraduate training, as well as the recruitment of foreign consultants to support clinical care and training. The economic argument for prioritisation of surgical services in the country is strong: surgical disease affects those that are economically active; a recent modelling study has shown that the macro-economic impact of untreated surgical conditions in the country is large (Grimes et al in print); and surgical care is cost-effective at the hospital level. However, more information is needed about the state of surgical services and barriers to access of care, to enable planning of services that are commensurate to need.

#### ASSET health system strengthening approach

In the ASSET project, we will use implementation and quality improvement (QI) methods to address barriers to providing safe, affordable and timely surgical care when required. We will work across the surgical platform (including perspectives from the community to tertiary care delivery) to develop, adapt and evaluate health system strengthening interventions (HSSI) to enhance delivery of guideline-based care. Core domains of HSSI and quality improvement for continuing care are well established, from research, and consensus. We will operationalise these into 5 HSSI elements:

1. evidence-based care pathways for high volume surgical procedures, with practice-based training, ongoing mentoring and support for implementation
2. Health Management Information System (HMIS) data to monitor and improve the quality of continuing care and patient outcomes, with specific focus on measuring detection, engagement on care pathways, adherence, retention and 'treatment to target'
3. a peer-driven QI culture, with motivated healthcare workers taking responsibility for improving care processes and outcomes (a 'learning health systems approach')
4. person-centred care reflecting patient values and preferences, with agreed care plans and patient empowerment for self-management
5. workforce strengthening to prepare for necessary organisational change, and foster non-technical skills (e.g. leadership, teamwork and communication, counselling).

There is evidence that these approaches can work, but, in sub-Saharan Africa, existing evidence is limited mainly to demonstration projects of single health system strengthening interventions rather than a basket of interventions across the whole health system; and interventions are rarely documented and evaluated with rigour. In ASSET Sierra Leone, a HSSI will be adapted to the Sierra Leone context, piloted and refined, and then applied as a package of measures to improve care delivery.

### ASSET Sierra Leone Introduction

#### Aims

The **aims** of the ASSET Sierra Leone research project are 1) to map care pathways, practices and patient experiences within primary, secondary and tertiary healthcare facility levels in the Western Area of Sierra Leone; 2) to assess the barriers to access to surgical care at the community level; and 3) to use implementation and improvement science methodologies to study the effects of interventions to improve achievement of increased access to care and improved quality of care.

Our aim addresses the following hypothesis: the volume of surgery can be increased, and the quality of surgical and anaesthesia care can be improved, by the introduction of a broad health system strengthening intervention that addresses challenges in the community, at peripheral health unit level and across secondary and tertiary care.

The study is divided into diagnostic and intervention phases. This protocol pertains to the diagnostic phase only (numbers 1 and 2 above); a separate protocol will be produced for the intervention phase of the research. The goal of the diagnostic phase is to comprehensively assess care and care-pathways for patients with surgical diseases from the community to the tertiary referral centre. In doing so, we will identify key steps where intervention to improve access to care and improve the quality of care is necessary and the methodologies by which to intervene to enable success.

#### Study sites

This is an exploratory study that will be done in the Western Area of Sierra Leone. This will allow capture of the country's only government tertiary referral centres that provide surgical care, their related district hospitals (DH), representative peripheral healthcare units (PHUs), and their communities. In addition, to enable ascertainment of some wider whole-country barriers in access to care, we will capture referral patterns from the entire country.

The Western Rural area has a population of 442,951 (6.3% of population of Sierra Leone), with 98.3 males per 100 females. Western Urban has a population of 1,050,301 (14.8% of population of Sierra Leone), with 98.8 males per 100 females.

In terms of healthcare facilities, the Western Area has 39 Maternal and Child Health Posts (MCHP), 28 Community Health Posts and 39 Community Health Centres (CHC). In addition, there are 11 government hospitals, 22 private clinics and 10 private hospitals (UNICEF 2015 survey report). In 2017, the WHO Service Availability and Readiness Assessment SARA survey (a health facility assessment tool designed to assess and monitor the service availability and readiness of the health sector and to generate evidence to support the planning and managing of a health system) was conducted in the Western Area. The survey reported that the Western Area had the lowest density of health facilities per head of population in Sierra Leone (1.2 and 0.6 per 10,000 for Rural and Urban, respectively) with inpatient bed density being lower in Rural (4 per 10,000) than Urban (11 per 10,000) wards. Maternal bed density in the Western Area is also the lowest in the country.

The study will be conducted at:

- 2 government tertiary hospitals: Connaught providing adult and paediatric surgery and PCMH providing obstetric surgery
- 2 secondary hospitals: Lumley and Rokupa
- 6 MCHP and CHC sites: Kissy, Blessed Mokada, Hamilton, Waterloo, Signel Hill and Tokeh

All studies will be conducted in one or more of these sites based on the level of care and appropriateness of data collection. Both tertiary sites were chosen to gain a better understanding of this system. It should be noted that PCMH involvement in the ASSET project is limited based on a number of logistical restraints of the larger project. Secondary sites were chosen through convenience sampling. The primary care sites were chosen through focus group discussions to identify a representative sample of sites based on a number of factors, including geography, patient population, patient numbers etc.

#### General permissions

We have verbal permission from Dr Deen, Deputy Chief Medical Director of the University of Sierra Leone Teaching Hospitals Complex to do this study and Dr AP Koroma M/S of PCMH Hospital and we will secure permission from the Minister of Health prior to doing our study. Details on individuals' consent are given under each described study section.

#### Ethical considerations

Ethical approval was granted from the Sierra Leonean Scientific Ethics and Research Committee and from the King's College London.

#### General sample size considerations

This is an exploratory study and, as such a sample size calculation in order to choose the number of facilities or patients to study does not apply. In terms of the site selection, we have chosen to site our study in the country's two government run tertiary referral centres providing surgical care and two government secondary hospitals in the Western Area providing surgical care. While it is not feasible to study all the Western Area PHUs, we will use stratified random sampling to select which type of PHU (MCHP or CHC) to study based on the distance to the nearest appropriate tertiary referral unit.  If we find in our study of referral data that some areas or MCHPs/CHCs are referring significantly more surgical patients than others, we will also stratify based on quartiles of patients referred. Regarding sampling for PHUs to include in the study, we estimate that studying 10 of the 78 PHUs will allow us to ascertain the major barriers in, and facilitators to, accessing quality care at these units and allow for generalizability across all PHUs.

Specific sample size considerations are given under each study-section. Details on sample size considerations for individual study elements are included within each section.

### Study plans

#### Study launch and Introduction meetings

At the start of the diagnostic phase, we will invite relevant stakeholders to an ASSET launch event and we will also hold introductory meetings where we outline the project and respond to any questions. We will not formally be studying the outcomes from the launch event or the introductory meetings, but they will serve to disseminate knowledge of the study amongst relevant parties in the Western Area and hence improve acceptability of any resultant planned intervention. Relevant stakeholders for the ASSET launch will include Ministry of Health & Sanitation colleagues and senior academic and clinical leaders. Relevant stakeholders for the introductory workshops will include: district health managers; community leaders, patient representatives, primary health unit managers and clinical providers; secondary care hospital managers and clinical providers; and, tertiary care hospital managers and clinical providers.

#### Situation Analysis

A situation analysis will be undertaken in order to provide a baseline understanding of (a) the potential structural challenges to improving access to high quality surgical, anaesthetics and dental care, and (b) of the adequacy of routinely available information about surgical, anaesthetic and dental care. The situation analysis will collate data that is available in the public domain using a modified version of the global ASSET situation analysis tool.

#### Hospital-level studies

##### *Hospital readiness to provide care*

We have created a succinct tool to assess readiness to provide care at tertiary or secondary (one tool for both levels of care), and primary health care level, based on SARA survey methodology, expert opinion, and the Harvard (PGSSC)-Ethiopian Facility Assessment Tool, to ascertain facility preparedness to care for the patient requiring  surgical, anaesthetic, obstetric, and dental treatment. Into this tool we have embedded data collector observations to allow validation of the responses.

The *Hospital Assessment Tool* will be administered to the hospital director/medical superintendent, surgical/anaesthetic/obstetric providers and operating theatre nurses. Data collection will take approximately one full day.

The *PHU Assessment Tool* will be administered to the PHU manager, and their identified healthcare workers Data will be collected by trained data collectors and physicians. Data collection will take approximately ½ a day. The data will be collected by trained data collectors and physicians. Data collection will take approximately ½ a day. Tertiary sites: Connaught. Secondary sites: Lumley, and Rokupa. Primary healthcare sites: Kissy, Blessed Mokada, Hamilton, Waterloo, Signel Hill and Tokeh. Statistical considerations and data analysis. This is a descriptive study, and no formal power calculation has been done.

To present results, a summary score will be created from the results from each section of the questionnaire. Additionally, we will summarize the scores for each section across all facilities assessed and describe these using measures of central tendency and spread. We will compare differences in score between methods using cross-tabulation. We will also compare data collected with our methodology and that of WHO SARA using summary scores and cross-tabulation. These results will then be descriptively compared to the results from the 2017 WHO SARA survey.

#### *Hospital patient safety culture survey*

We will assess patient safety culture using the Hospital Patient Safety Questionnaire. Note, in 2017, the survey was distributed to staff at Connaught for self-completion. A total of 50 responses were returned (74% of those sent out). This study aims to build upon this data.

For this current study we will repeat the survey in a pilot study at Connaught by administering the questionnaire to staff by a trained research assistant. We will compare the quality (response rate and missing-ness of data) of the responses and the response rate between the self-administered 2017 survey and data-collector administered questionnaire. If there is no substantial difference in the quality of the data collected, we will deliver the questionnaire to the staff at Connaught by the methodology with the best quality data.

Tertiary sites: Connaught. Secondary sites: Lumley, and Rokupa.

#### *Statistical considerations and data analysis*

The survey tool website (https://www.ahrq.gov/sites/default/files/wysiwyg/professionals/qualit patientsafety/patientsafetyculture/hospital/userguide/hospcult.pdf) provides guidance on sampling size. For facilities with 500 or fewer staff, the recommendation is to include all staff anticipating a 50% response rate. For facilities with 501-999 staff, the recommendation is to sample 500 staff anticipating 250 responses (50% response rate). As the total staff of the 2 tertiary hospitals and 2 secondary sites is between 501-999 staff, we will sample 500 staff from across these sites (based on proportion of total number of staff providing surgical care at each sites and proportions of cadres making up total staff. Thus, the numbers from each site will be:
*A (n value) = Y (consultants) + Z (theatre staff) + B (nurses) + C (Admissions staff)*

Site A, total number sampled = X of which Y are consultants, Z are theatre staff, B are nurses, C are admissions staff, and etc). The target sample size is based on three assumptions: simple random or systematic random sampling, a response rate of 50 percent, and a confidence interval of +/− 5 percent.

Results for each section of the questionnaire will be described using measures of central tendency and spread, and the overall response summed and described for the cohort. Response rate, results for each section, and overall scores will be compared between the self-administered and data-collector administered questionnaires and compared using cross-tabulation. Results will be disaggregated by provider type, and differences between provider responses will be explored, whilst acknowledging that we have not powered the study for this analysis.

#### *National referral process mapping and geographical origin*

Data will be prospectively collected over a two-month period from that captured by Sierra Leone wide Referral Coordinator scheme. Data are collected in the following domains: patient origin (geographical area and health-facility type and name), time of referral, reason for referral, referral centre's perceived urgency of referral, time of arrival at receiving facility, discharge diagnosis, and outcome. Tertiary sites: Connaught

Analysis of the data to enable description of distance travelled between referral facilities, time taken to travel between facilities, and outcomes of patients referred has been done. This has disaggregated results by type of referral facility, urgency of conditions, reason for referral, and final diagnoses. Further, general prospective analysis – as had been planned - will not add substantively to this analysis.

#### *Clinical pathways process mapping*

We aim to ascertain clinical pathways and undertake process mapping for surgical patients at Connaught hospital – from admission to discharge - within the hospital environment.

We will map the possible routes of, and processes experienced by, patients as they transit through the hospital, and describe the reasons behind possible pathways. Thus, for patients who require surgical care, we aim to capture information on the following themes:

1. Local guidelines for clinical pathways and processes used in various areas in the hospital
2. Local/staff deviations from these guidelines

This will be done through a combination of focus groups, observations, and ethnographic work. Focus groups with care providers (physicians and nurses who work within the surgical and/or obstetric care pathways) and managers will be conducted to ascertain the pathways and processes for surgical patients in their area of care. Questions will be open and around the above themes. Based on these responses a preliminary map will be created, which will subsequently be presented to local stakeholders and research participants for validation.

We will do ethnographic work to ascertain patient experiences, as they transit through the hospital and also observe the way they are treated during these processes. Additionally, we will observe the interactions between different cadres of staff and between staff and patients. Following the mapping process further in-depth qualitative work will be undertaken with hospital care workers of all cadres to identify the wider barriers and facilitators to accessing and providing quality surgery care. We aim to do around 20 Key Informant Interviews (or until saturation of responses is reached) across all cadres of healthcare workers in each of the hospitals (tertiary and district) participating in the study.

The process mapping study element will be done in tertiary hospitals only. See the table for examples of potential clinical areas and processes to be observed.

*Table 1 Patient pathways and processes*

| | |
|---|---|
| Hospital main gate | Any restrictions or delays on entering the hospital compound?<br><br>Where do ambulances get sent? |
| Triage room/ emergency department | Are there any delays between presentation to hospital and initial triage? What are the reasons for delays (e.g. needing to purchase a card, presenting during lunchtime, etc).<br>Who carries out the triage? Do they measure vital signs? Are they documented? What happens next if the person is considered to have a surgical condition? To whom and how is this information communicated? |
| Surgical assessment | At what stage would the patient be reviewed by a surgical, obstetric, or anaesthetic care provider (medical officer level, general practitioner, surgeon or anaesthetist or obstetrician/ gynaecologist)? Who else is involved in the diagnostic process (patient, family members?) and what information is provided by the patient that might facilitate referral? E.g. about connections or payments. What are the reasons for delays before surgical review? When the decision to operate is made, what happens next? What observations are conducted while the person is waiting for surgery? What else needs to happen before the surgery can take place? What happens if the person needs surgical referral? How long do they wait in the hospital before transfer? |

| | |
|---|---|
| Surgical ward | SSIs. How are surgical wound infections documented? How is information on wound infection collected? Do surgeons know their wound infection rates? Where do patients go if they develop wound infection after hospital discharge? What antibiotics are available? How are antibiotics obtained? How and when are antibiotics given? Who administers antibiotics? How long are antibiotics continued? What protocols exist for peri-operative administration of antibiotics?<br><br>Sierra Leone Early Warning Scores (SLEWS) and the deteriorating patient. Are vital signs collected on every nursing shift? Are SLEWS measured and recorded accurately? Are vital sign measurements increased when SLEWS scores increase? Are nursing staff calling medical staff when SLEWS scores increase? Do medical staff respond to calls from nursing colleagues?<br><br>Preparation for theatre.<br>Are there guidelines for preoperative fasting for patients? Are there preoperative anaesthetic assessments? How do theatre and ward staff communicate prior to a patient going to theatre? |
| Operating theatre | WHO checklist.<br>Is the WHO or other locally appropriate safe surgery checklist implemented? What are the interactions between staff involved in implementing the checklist? Who is in charge and if steps are missed how are these corrected?<br><br>Theatre sterile procedures.<br>What is the process for skin antisepsis and the availability and use of antiseptic agents? How are hands washed and scrubbed? How are gowns and drapes washed, dried and sterilised? How are gowns and drapes mended? How sterile gloves procured? How are sterilisers maintained? How are instruments cleaned? How are instruments sterilised? How is sterilisation confirmed? How are swabs tracked during surgery? How often are swabs counted? What happens when there is a discrepancy in swab numbers? |
| Post-operative recovery room | What instructions are communicated and in what manner (including tone) to the recovery room staff about observation level and warning signs? How frequently are observations carried out? What happens if the patient deteriorates? |
| Post-operative surgical ward | What instructions are communicated and in what manner to the post-op surgical ward staff about observation level, warning signs? How frequently are observations carried out? What happens if the patient deteriorates?<br>What information is recorded at the point of discharge from hospital? Is there a discharge summary or letter? Who gets a copy of it? What instructions are given to patients in terms of what to do/who to seek advice from and where post-discharge? |

Statistical considerations and data analysis. This is a descriptive study, including a largely qualitative element. Information from the mapping process will be presented visually. The number of interviews will be informed by the saturation criterion, such that we will not seek to interview more participants once the themes that emerge from the interview start repeating themselves.; from previous work, we estimate that 3-4 iterations of the focus groups with 4-5 clinical providers will provide high quality information to enable construction and validation of the care process map. Interviews will be digitally recorded and transcribed verbatim for subsequent thematic analysis, using NVivo. Primary and secondary themes that emerge from the interviews will be tabulated and reviewed by the research team, as well as cross-checked with a small sample of participants for accuracy (member-checking technique). The final themes will be tabulated and supported by verbatim quotes from the participants. Information from the focus groups which has informed the development of the maps and the ethnographic study will be analysed using NViVO. The results of the process-observational study and document review will be summarized using descriptive statistics where appropriate and thematic analysis.

*Hospital documentation study*

This study at Connaught will assess clinical documentation at baseline, identifying key documents and records and gaps in documentation that need to be filled, to develop and implement a hospital performance measurement tool.

Documents included: initial triage forms, clinical records, nursing records (if separate), surgical checklists, and any additional forms or registrations books that are completed. The documentation will be identified from all relevant clinical settings; for example, triage room, emergency department, obstetrics ward, surgical ward and operating theatre.

To do this study we will:

1. Create a list of the types of documents routinely used (eg medical notes, anaesthetic charts, theatre registry, ward registry).
2. Create a list of recorded parameter (clinical and process) entered into each document
3. Create a list of parameters which are both are feasible to collect and can act as sentinels or markers of the state of note keeping in each document type. Examples of parameters include:
    a. Medical notes: vital signs, state of surgical wound, preoperative blood results
    b. Anaesthetic charts: American Society of Anaesthesiologists (ASA) category of physical status classification score
    c. Theatre registry: urgency of case
    d. Ward register: Documentation of patient outcome – discharge or death
4. For each parameter-type selected, we will create a list of the frequency with which data should be collected
5. Create a scoring system for completeness of data collection for each parameter.

Data collection for this study will run concurrently with the cohort/note review study using the same data collection tool.

#### *Hospital data capture studies*

This study will build on the hospital documentation study, assessing specific surgical indicators.

The clinical documentation of aspects of care for a person presenting with a surgical condition will be investigated, including initial triage forms, clinical records, nursing records (if separate), surgical checklists, and any additional forms or registrations books that are completed. The documentation will be identified from all relevant clinical settings; for example, triage room, emergency department, obstetrics ward, surgical ward and operating theatre.

Three major areas will be explored through this study:

1. Theatre productivity

2. Time to emergency theatre
3. Bed occupancy

Data will be captured from Connaught prospectively to determine the number of hours in each standard working day in which the theatre is in use during the working day is defined as 08.30am-17.30 [or other pre-defined end of working day]. Measurement points are: (1) date, (2) theatre name, (3) time the list was planned to start, (4) participant ID, (4) procedure, (5) OR time in and out, (6) number of patients cancelled, and (7) reason for not in use. Statistical considerations and data analysis. These are pilot studies, there has been no formal power calculation for this exploratory study. Measures of central tendency and spread will be described and - where appropriate - trends will be presented graphically.

The length of time for surgical patients to transit from emergency admission to theatre. This data will be captured prospectively using review of notes as part of the note documentation/patient cohort study proforma. Data will be captured for this study over a 3-month period (2 months where all new patients admitted are entered into the study and one further month where patients entered within the first two months are followed up). Statistical considerations and data analysis. Data will be described for each facility as central tendency and measure of spread.

Bed occupancy rates at Connaught hospital. This aspect of the study will assess the patient load over the course of the period of study based on bed occupancy. This will be captured prospectively over a four-month period, using information supplied daily by the Referral Coordinators. Statistical considerations and data analysis. Data will be described per facility as average (and spread) occupancy and daily trends presented graphically.

#### *Patient cohort studies (costs, experiences, outcomes)*

*Financial implications of care*

We will assess the direct and indirect costs of accessing surgical care and the % of people suffering catastrophic or impoverishing (pushing household below poverty threshold) as a result of accessing care. We will do this element of the study in people who have undergone major surgery at Connaught and in people who have had C-sections in PCMH.

We will do face-to-face interviews at time of discharge to ascertain costs incurred in accessing care (both direct – for example, costs of paying for medical care as well as costs of travel to medical care – and indirect – for example, loss of earnings due to accessing care). We will also assess household expenditure to enable assessment of impoverishing and catastrophic expenditure (both of which are calculated based on normal household outgoings). Methods to ascertain costs of accessing care methodology and household expenditure will be based on an adapted Client Services Receipt Inventory and WHO SAGE questions on household income and expenditure; this methodology has been previously adapted for surgery and used in Uganda. We will question both patients and their relatives to enable documentation of full costs of care (for example, this may involve relatives purchasing supplies from outside of the hospital). Interviews will take between 30 and 60 minutes. We will use the World Bank definition of poverty of spending less than $3.10 per person per day and extreme poverty of spending less than $1.90 per person per day. We will use the definition of impoverishing expenditure as one which results in the household being pushed into poverty and catastrophic expenditure as one which results in i) the household spending more than 40% of its non-subsistence expenditure or ii) the household spending more than 10% of its total annual expenditure.

Statistical considerations and data analysis. A recent study in Uganda has shown that although the Lancet Commission on Global Surgery estimated 80% of patients undergoing surgery were at risk of financial catastrophe, actual proportion of admissions experiencing catastrophic expenditure was 60%. To thus capture a proportion of catastrophic expenditure of 60% with a CI of 50-70% will require a sample size of 386 patients. Allowing for drop-outs at 10%, we will require 426 patients. We will present data as number (%) of patients who have experienced catastrophic or impoverishing expenditure.

*Experiences and outcomes.*

We will identify patients who are willing to be followed up in the community 30 days after discharge either from patients who are admitted on the ward, or at the end of the interviews on costs of accessing care. For patients who agree, we will capture their contact details and address and confirm an approximate time when we will approach them in the community. We will study patients who have had a laparotomy, hernia, appendicectomy, or amputation.  Patient experiences will be captured using a modified version of the I-PAHC questionnaire, which was developed and validated in Ethiopia. PROMS will be collected using a modified WHODAS questionnaire which focuses on the ability to carry out standard activities of daily living (walking across a room, getting out of bed, eating, dressing, washing/showering/bathing, going to the toilet) and whether or not care needs are met. The period of focus for these questions will be in the past week. Patients will be followed up 4 weeks after surgery. The study will be done using trained research assistants. Statistical considerations and data analysis. This is a pilot study and we have not done a formal power calculation. Sampling will therefore be pragmatic, and we estimate that to give an indication of patient experiences of care and PROMs with which to power future studies will require follow up of 20 patients

##### *Health care provider study*

This study will explore barriers and facilitators to accessing surgical care from the provider perspective. Health workers across cadres at the tertiary and district level will be included in this study. For this study element we will undertake a series of key informant interviews with healthcare workers at the hospitals PHUs. Sampling will be purposive. The study will also include focus groups the purpose of these focus-groups will be first, to contextualise the experiences of HCWs within the broader social landscape of health provision and second, to elucidate some of the local concerns and therapeutic pathways for surgical care.

We will address the following questions:

1. What are the healthcare providers perceptions of their ability to recognize and provide care for the surgical patient?
2. What are the barriers and facilitators to enable the provision of that care?
3. How they understand and differentiate the quality of service delivery?

Statistical considerations and data analysis. The number of interviews will be informed by the saturation criterion, such that we will not seek to interview more participants once the themes that emerge from the interview start repeating themselves, We expect to interview 15-20 providers at each site. Interviews will be done in Krio or English by research assistants who have been trained in qualitative methodologies. They will be recorded, transcribed, translated, and triangulated using a framework approach and coded and thematically analysed with the support of NViVo software. Follow up focus-group discussions will be undertaken in select communities where HCWs live.

#### Community studies

*Community exploration of barriers and facilitators to accessing surgical care.*

Three groups of interest: those who underwent a surgical procedure (from the cohort study above), those who sought care but did not undergo surgery, and those who did not seek care (snowball sampling/ KI).

This study will explore barriers and facilitators to accessing surgical care from the patient perspective. Three groups of interest will be interviewed: those who underwent a surgical procedure (from the cohort study above), those who sought care but did not undergo surgery, and those who did not seek care.

We will address the following questions:

1. What are the healthcare providers perceptions of their ability to recognize and provide care for the surgical patient?
2. What are the barriers and facilitators to enable the provision of that care?
3. How they understand and differentiate the quality of service delivery?

Statistical considerations and data analysis. The number of interviews will be informed by the saturation criterion, such that we will not seek to interview more participants once the themes that emerge from the interview start repeating themselves, We expect to interview 15-20 patients. Interviews will be done in Krio or English by research assistants who have been trained in qualitative methodologies. They will be recorded, transcribed, translated, and triangulated using a framework approach and coded and thematically analysed with the support of NViVo software.

#### Diagnostic phase results synopsis

We will collate the information obtained from documentary and observational sources and triangulate with quantitative and qualitative data to identify key system bottlenecks. This information will (1) be fed back to the all stakeholders at Ministry of Health, clinicians, hospital management and patient groups, (2) used to inform the development of the intervention/s in the subsequent ASSET programme phase, through use of a 'theory of change' workshop-delivered methodology (see below) with local stakeholders aimed at improving access to quality surgical care, and (3) used to design research tools for capturing process data, to be used in the piloting and implementation phases of the project.

#### Theory of Change workshop

Theory of Change (ToC) is a participatory approach that is employed to develop a contextually valid roadmap to achieve a shared program outcome. This can be achieved by bringing stakeholders together in workshops. The first task is to build consensus on the most important outcome of the program. The group of stakeholders will then work backwards to map out the intermediate outcomes that need to be achieved on the way to the final outcome. The interventions needed at each step in the pathway are determined by reviews of the existing evidence base combined with local experience. Assumptions underlying the steps on the pathway are made explicit and investigated using research studies. Indicators of success are also specified. The output of ToC is a descriptive 'map' which specifies the necessary elements of complex interventions in a logical manner and time sequence as required, the approach to implementation and the indicators that need to be evaluated. In this way, the ToC process enables stakeholders to come to a common understanding of what the interventions will achieve, and how they will achieve their goal. The ToC map also provides a framework for evaluation of the complex interventions.

At the end of the diagnostic phase we will analyse results and prepare the results for presentation as part of a 2 day Theory of Change workshop, facilitated by the research team. Participants who will be invited to the ToC workshop include the Ministry Staff (including from Directorates of Hospitals; HR; Policy, Planning and Information), District Management Teams (DMO and representation from secondary care and PHUs), Teaching Hospital Complex (leaders and care providers and managers) and NGOs providing surgical care. The purpose of the ToC workshop is to elucidate the route to improving access to quality surgical, anaesthetic and dental care in the Western Area. Informed consent will be sought from ToC participants and the workshops will be audio-recorded. The outputs of the ToC workshops will be a full ToC map and a report of how key decisions were made in order to agree on the ToC map.

The output from the ToC workshop(s) will be a context-specific health system strengthening intervention that is ready to pilot across PHUs, secondary and tertiary sites in the Western Area.

### Ethical considerations for ASSET Sierra Leone

Ethical approval will be obtained from the Sierra Leone Ethics & Scientific Review Committee, Ministry of Health & Sanitation, and from King's College London's College Research Ethics Committee, UK.

Specific ethical considerations are presented in relation to each of the individual ASSET studies. Here we present the overall ethical considerations that are relevant across the studies.

#### Informed consent

All participants will be informed about the study and given the opportunity to ask questions before being invited to consent to participate. Only people who give voluntary informed consent will be included in the study. We will seek written consent wherever possible, but if the person is not literate, we will ask a literate witness to sign to confirm that the information sheet has been read out correctly and verbal consent provided. In such cases we will ask the person to give a thumb print to signify their consent to participate.

#### Data protection

All paper-based records collected in this study will be stored in a secure filing cabinet or cupboard. All electronic data resulting from the project will be stored on a secure and encrypted computer network and/or password-protected computer and external hard drive. All data will be encoded so that the interview and test results cannot be directly linked to contact details. Data relating to personal information of the participants (e.g. their names and contact details) will not be stored on portable devices or media, but only in highly secured systems, and the paper-based records (e.g. the signed informed consent forms) will be stored securely and separate from the questionnaire and interview data. Only members of the research group will have access to the data. Audio files will be encrypted and password-protected and only identifiable by the participant's project identification number (making sure that the names of the participants are not included in the recording). Personal data collected for the study will be stored until 2 years after the study is completed. Other research data will be stored until 7 years after completion of the study.

#### Recompense to participants

Where appropriate, we will reimburse participants travel costs and provide refreshments. We will also provide a bar of soap or a packet of biscuits and a drink to all participants in the qualitative studies.

#### Deception

No deception is involved in this study. The nature of the research is explained in the information sheets (see Annex).

#### Risk of harm to participants

We will minimise the risk of harm by ensuring that participants are fully informed about the study prior to voluntary participation. In the event that a participant wishes to withdraw from taking part in ASSET or from contributing their data, they are free to do so.

We recognise that some participants could possibly become distressed when interviewed about their health condition, or ability to pay the costs of care. We will seek to minimise this occurring in the first place by ensuring that questions are worded sensitively. We will ensure that our data collectors are trained to be sensitive to signs of distress and to offer to reschedule or discontinue the interview if distress is noted.

*Benefits*
The contributions of participants will help to improve the healthcare delivered in the study districts. The study will also build capacity in health workers and healthcare administrators in the study districts. The findings of this study are likely to apply to other low-income African settings too.

*Confidentiality*
Confidentiality will be maintained throughout. Interviews will be conducted in private. Confidentiality of collected data will be maintained as described in the Methods and Data Protection sections.

*Declaration of conflicts of interest*
None of the collaborators have any potential conflicts of interest with the proposed project.

*Quality assurance*
The project PIs (AL and JD) will visit the study site on a regular basis in order to ensure that the ethical principles outlined above are adhered to, including random checks of the procedure for obtaining informed consent. This will be supported by our established procedures for ethical data management (see Methods and Data protection sections).

### Timetable for diagnostic phase in SL

| Project activities | 2018 | | | | | | | | | | | | 2019 |
|---|---|---|---|---|---|---|---|---|---|---|---|---|---|
| | Jan | Feb | Mar | Apr | May | Jun | Jul | Aug | Sept | Oct | Nov | Dec | |
| Recruitment of research assistants | | | | | | | | | | | | | |
| Training research assistants | | | | | | | | | | | | | |
| Study introductory meeting | | | | | ■ | | | | | | | | |
| Situational Analysis | | | | | | ■ | ■ | | | | | | |
| Hospital readiness to provide care assessment | | | | | | ■ | ■ | ■ | ■ | | | | |
| Hospital patient safety culture | | | | | | | | ■ | ■ | | | | |
| Sierra Leone referral pathways | | | | | ■ | ■ | ■ | ■ | ■ | | | | |
| Hospital pathway and process mapping | | | | | ■ | ■ | ■ | ■ | | | | | |
| Hospital documentation analysis | | | | | | | | | ■ | ■ | ■ | | |
| Other processese of care (eg POMR and WHO checklist) | | | | | ■ | ■ | ■ | | | | | | |
| Surgical cohort study | | | | | ■ | ■ | ■ | ■ | ■ | | | | |
| Qualitative study (providers) | | | | | ■ | ■ | ■ | ■ | ■ | | | | |
| Community readiness to provide care | | | | | | | | | ■ | ■ | ■ | | |
| Diagnostic phase synopsis | | | | | | | | | | | ■ | ■ | |

### ZIMBABWE: WORK PACKAGE 8

#### Integrated Care for Hypertension, Diabetes and Depression in Zimbabwe

**PRINCIPAL INVESTIGATOR**

**Dr. Dixon Chibanda, PhD**

+263 712 204 107;
University of Zimbabwe,
College Of Health Sciences, Research Support Centre
Parirenyatwa Hospital Grounds

**CO-PRINCIPAL INVESTIGATOR**

**Dr. Ruth Verhey, PhD**

#### List of Acronyms

GBC-Guideline-Based Care

CMD-Common Mental Disorder

FB-Friendship Bench

QI-Quality Improvement

ToC-Theory of Change

NCD-Non-Communicable Diseases

PHC-Primary Health Care

DHPO-District Health Promotion Officer

RCT-Randomized Control Trial

LHW-Lay Health Worker

SSQ14-Shona Symptom Questionnaire

SOPs-Standard Operating Procedures

ACASI-Audio Computer Assisted Interview

ODK-Open Data Kit

### Background

There is a high burden of common mental disorders (CMD) globally [1]. CMD comprising depression, generalized anxiety and stress-related disorders are rarely recognized or treated in low and middle-income countries (LMIC) [2].

Comorbidity of CMD and non-communicable diseases (NCDs) such as hypertension or diabetes is contributing to the severity of the disease manifestation [3].Depression is associated with hyperglycemia and increased risk for diabetic complications. People who suffer from depression usually have challenges with adherence, hence conditions like hypertension and diabetes conditions could worsen[6].More than 50% of people aged 15-59 years suffer from NCDs in low and middle income countries (LMIC)[7]. Growing evidence also suggests high level of comorbidity between hypertension and depression. In South Africa, 16.7 % of adults reported a previous medical diagnosis of hypertension and 8.1% were found to have anxiety and depression [8]. Studies have demonstrated that a comorbid state of depression incrementally worsens health conditions such as diabetes and hypertension [8]. The impact of depression on quality of life and its potential negative effect on diabetes management warrant recognition and treatment of the disorder in diabetic individuals [9].

Depression and diabetes contribute enormously to the public health burden. Identification of risk factors for these disorders is thus a necessary and important strategy[7].

Recently, a rigorous randomized control trial (RCT) of a low intensity psychological intervention called the Friendship Bench Program (FB) which is delivered on primary health care (PHC) level by trained lay health workers (LHWs) was completed in Zimbabwe [2]. This task-shifting intervention is based on cognitive behavioral therapy (CBT) and offers problem solving therapy (PST) in a series of up to six sessions followed by a voluntary participation in a peer support group. Results of the trial showed that the FB was highly effective at treating CMD in adults. The FB has been scaled up to 72 clinics in 3 cities in Zimbabwe and has reached more than 40.000 beneficiaries in the last 2 years. The scale up provides the ideal framework to introduce and evaluate a health system strengthening intervention that focuses on 3 critical NCDs (depression, hypertension and diabetes). Addressing these 3 conditions through an integrated approach could have major public health benefits.

### Rationale

The results of the Friendship Bench (FB) trial showed that the FB is highly effective at reducing common mental disorder (CMD) symptoms in the subgroup with comorbid CMD and HIV, and highlighted that CMDs are a major cause of poor adherence to antiretroviral therapy and hence virological failure. Available data from the FB shows that a significant number of people living with hypertension, diabetes and other chronic conditions have benefited from the program. We would like to further explore how the FB may continue to contribute towards health systems strengthening through this study. Transcripts of FB sessions indicate that the FB may help participants to improve their adherence to treatment. We propose to use thistask-shifted care of the FB as a key universal strategy to strengthen health systems linked to the Friendship Bench through ongoing training, support, and guideline-based care, patient centered care, through understanding care pathways, and peer driven QI in all clinics that the FB has been scaled up to.

### Aims

The study aims to develop and evaluate an integrated care package for hypertension, diabetes and depression (Friendship Bench Plus) in PHC which will:

1. Improve care processes and outcomes such as quality of life and disability

2. Strengthen the use of guideline-based care which is patient centered.

**<u>This will be achieved through the following objectives:</u>**

1. Review of the existing Guideline based care packages for all three conditions.
2. A description of local care pathways of the three conditions.
3. This study is going to use the mixed method approach. The qualitative study will be done through Semi structured interviews to have a situational analysis in the three conditions (diabetes, hypertension, depression). Quantitative methods will include collection of existing data from clinics on the three conditions, prevalence of the conditions, quality of life, care pathways and disability of participants.
4. Develop a package that emphasizes patient centered care and peer driven quality improvement (QI) as components of a health systems strengthening intervention based on findings from objectives.
5. Pilot and evaluate the final package with all the above 3 components for patients using PHC with the 3 NCDs.

This proposal focuses on the diagnostic phase. Objectives4-5 will be informed by the first three objectives; therefore, an amendment will be submitted to this proposal once these are clarified.

### Methodology

#### <u>Overview of study design</u>

The study will use a mixed method approach i.e. qualitative and quantitative methods to collect data from healthcare workers and patients in the study PHCs. We will also rigorously assess existing data held at health care level including patient held treatment cards/books etc.The study will be divided into three main stages which are a) Diagnostic phase, b) Health System Strengthening Intervention (HSSI) Package development and c) evaluation. The stages and the respective research methods to be used are described below.

**Setting:** The study will be conducted in nine(9) local government operated PHCs across Harare where Friendship Bench has been introduced. The PHC facilities in Zimbabwe operate daily providing extensive care for both communicable and non-communicable diseases. The PHC are categorized according to size, staff compliment into; poly-clinics n=20(catering for approximately 150 patients a day), satellite clinics n=32(60-80 patients daily) and Family Health Clinics n=20 (40-60 patients a day). All the PHC are equipped to provide basic care for the 3 conditions (depression, hypertension and diabetes) with depression included as part of the Friendship Bench care package delivered by LHWs. For this study the 9 PHCs will be purposively selected from the 3 categories to ensure a balance in distribution of PHC variables.

### Research Methods

#### <u>Stage 1: Diagnostic Phase</u>

<u>A quick review</u> will be carried out in 9PHCs in Harare to establish the current situation with regards to the 3 NCDs (depression, hypertension, Diabetes) and how treatment is accessed together with follow up care (To inform current guideline based care and pathways to care). This will be done by collecting existing data at the clinics through the clinic register, patient held data (see iii below). Specific data collecting tools attached in the appendix (i) will be used to collect this data using trained research assistants. Consent will be sought to collect the data (see annexed Informed consent form) Results from the quick review will be used to inform on the existing i) referral pathways, ii) diagnosis &medication use/availability, iii) frequency of PHC visits iv) comorbidity and v) clinical outcomes. We will triangulate this data using semi-structured interviews with staff

(nurses, nurse aides, doctors, LHWs, receptionist, clinic managers and policy makers and alternative health providers (AHP) as described below. We will through this situational appraisal seek to learn more about existing resources, the nature of consultations, duration, frequency and quality. We will also assess patient records and the quality of all clinic/patient related documents.

For instance, how care is practiced in these PHCs will be assessed by going through the clinic held records and interviewing patients and staff. We will use checklists to assess continuity of care, drug supply chain. For instance, we will want to know if people being treated are reaching the clinic's intended target in terms of recovery. For this we will also use an attendee survey of those utilizing the PHC to assess management of their clinical conditions and determine the prevalence of the 3 NCDs.

We will test blood glucose using glucometers, and record time of last meal, and we will measure blood pressure as described below. For this initial phase we will allocate 3 research assistants to each of the 9 PHCs to systematically review data held at the clinics using a checklist developed by our team in consultation with the city health department. Data collected during this first step (i) will be triangulated with subsequent data collected as described below.

Quantitative study- As part of the review described above, a cross sectional survey of patients utilizing the 9 PHCs included in the study will be carried out over a 2 month period .We will initially group all PHCs (n=72) according to similarities by size/ staff compliment, conditions treated, and catchment are. These clinics will then be grouped into 3 categories (High, Medium, and Low). From these 3 categories we will select 3 clinics per category and from each PHC in each category we will recruit patients (n=240) with a total sample size of 2160 participants. A socio-demographic questionnaire including screening tools SSQ14/PHQ9/WHO-DAS, questions for cardio-vascular risk factors (CVRF) will be administered all 2160 participants. This will be followed by blood glucose through glucometer and blood pressure with a digital blood pressure measuring device. In addition a sample of urine will be collected from every participant for urinalysis using urine-dipstix.

#### *Sampling*

In the identified 9 clinics all those aged above 18 attending the PHC during the 2 month period will be eligible for inclusion. Every clinic visitor during this period will be allocated a sequential number (based on time of arrival) commencing from number 001. Each numbered card will have a unique code( from A001) which will be randomly selected using a computer generated random number sequence to determine which participants are approached to join the study.

To ensure a balance in allocation we will over sample those aged 35 and above. The final sampling procedure will be determined after a 1-week pilot of the 9 PHCs.

#### *Inclusion criteria*

Those aged 18 and above and are able to understand the consenting process will be enrolled provided they give written informed consent.

#### *Exclusion criteria*

All those who are below 18, or are unable to comprehend the nature of the study or are physically unwell including those who are psychotic or severely depressed and suicidal will not be included in the study.

#### *Procedure*

Three trained research assistants will be based at each of the 9 facilities to sensitize patients as they wait to receive care. The research assistants (RA) will administer a socio-demographic questionnaire to all participants (who give written informed consent) in a quiet room reserved for the study. The socio-demographic questionnaire will also include questions on SSQ/PHQ screening for CMD/depression and presence of cardio vascular risk factors (CVRF) which will include alcohol use, smoking, underactivity and obesity (weight and height to be measured during the blood pressure measurement). After the socio-demographic questionnaire is administered a study nurse will measure blood pressure 3 times with 2-3 minute intervals (while participant is seated) with the mean of the 3 recordings taken as the final participants BP. After the blood pressure is taken the nurse will collect a small amount of blood using a pin-prick and measure blood glucose using glucometer. Both BP measurement and pin pricks for thumbs, glucostix, and point of care will be guided by existing clinic protocols. Those immediately found to either meet criteria for any of the 3 NCDs will receive further questions where we will seek to establish if the participants were ever told about their existing condition, if yes by whom and how? We will seek to establish what help they are getting for their condition, medication, counseling, educational, dietary and exercising input. We will use the data to inform on the real prevalence of the 3 NCDs (known cases + unknown (new cases)) for each clinic. In this group (those with confirmed NCD) we will also enquire about care pathways/patient centered care to augment the qualitative component described below.

#### *Data collection*

Data will be collected using computerized "Samsung" tablets. Our team has experiencing using this kind of data collection method where data is then electronically stored using cloud computing.

**Qualitative:** All those who meet criteria for the 3 NCDs in the quantitative component above will be eligible for inclusion in the qualitative component. The rest of the participants (nursing staff, doctors, LHWs, mangers, policymakers and clinic support stuff) will be approached separately at each of the 9 clinics. Semi-structured interviews will be carried out by trained interviewers. The study design will take anthropological methodology (in-depth personal interviews) to produce qualitative data. These methods were chosen because they allow for exploration of local perceptions of treatment/care provided at clinic level. Experiences with illness and care are complex phenomena that are best explored through long-form, open-ended interviews that allow participants to share experiences. Qualitative interviews with patients, health workers and policymakers will allow for the generation of new ideas about strengthening health systems in the local context that may not emerge from a quantitative survey alone.

#### *Sample selection*

From each of the 9 clinics described above all those meeting NCD criteria of depression, hypertension and diabetes will be approached and informed of their findings then asked if they would want to proceed with the qualitative component of the study. Although we will purposively select participants for this we will ensure a balance between gender and NCD condition.

For the nursing staff, LHWs, alternative health providers (AHP) receptionists, policy makers and doctors' purposive sampling will be used. In a given clinic the nurse in-charge will be approached first for the first interview and after this she will be asked to recommend other key informants based on the different roles required to be interviewed.

The sample size (n=34) was determined based on the number that we expect will be needed to reach theoretical saturation, the point at which no additional information emerges from subsequent interviews. Theoretical saturation is a typical and rigorous form of determining sample size in qualitative research. The experience and expertise in qualitative methods of the research team will be used to determine the necessary number of participants. Fewer will be interviewed if theoretical saturation is reached prior to meeting the sample size indicated above. We anticipate that the sample size will be approximately 34 (individual interviews), but this may be adjusted based on the number needed to reach theoretical saturation.

#### *Specific recruitment strategies by participant type:*

**Patients:** Will be recruited from the cross-sectional study described above based on the presence of the NCDs. Clinical and study records will also be reviewed as described above to triangulate information obtained from the patients during the interviews.

**LHWs:** Initially, the supervisor of the LHWs will be contacted and asked to suggest a number of LHWs who display diversity across the following characteristics: age, gender, community, and attitude toward the PHC/Friendship Bench/ care services. Those LHWs will be contacted and asked to participate in an interview. At the end of each individual interview, the LHW will be ask to suggest other LHWs who may be able to provide new or important perspectives to the study, then those individuals will be contacted by the interviewer and invited to participate.

**Nurses:** Initially, the health center lead/nurse supervisor will be contacted and ask to suggest nurses who display diversity across the following characteristics: age, gender, community, experience, and involvement in hypertension, depression and diabetes care. Those nurses will be contacted and asked to participate in an interview. At the end of each individual interview, the nurses will be ask to suggest other nurses who may be able to provide new or important perspectives to the study, then those individuals will be contacted by the interviewer and invited to participate. This approach will also be used for the other health care workers which include doctors, managers, and support staff.

**Alternative health providers (AHP):** Using the LHWs who interact regularly with the community we will obtain information on the AHPs in each catchment area of the 9 clinics. We will also use information obtained during the quantitative phase to identify patients who have previously used AHP for their conditions. These patients will be asked to provide contact details of their AHP if they feel comfortable to do so. In the interviews with the AHP we will seek to elicit the complimentary use of AHP. In the interviews with AHP we will seek to understand how AHP recognize symptoms of depression diabetes, hypertension and how they decide to refer and where they refer their clients.

#### *Data collection*

##### *Individual interviews*

Individual semi-structured interviews for in-depth exploration of personal experiences and perspectives will be carried out.

All participants will have approximately 60 minutes, in-depth, mostly open-ended personal interviews that seek to examine their experiences with the PHC. The individuals will be asked about their experiences with and perspectives on being referred and managed at PHC for their condition. The interview questions are attached to this proposal; these questions will be used to guide the conversation between the interviewer and participant. The trained interviewer will introduce additional questions to elicit additional information on key themes based on participant responses.

Interviews will be conducted in English or Shona, based on the preference of the interviewee. The interviewers will be bilingual in Shona and English and trained in qualitative methods. Interviews will be conducted in a private space near the health center or in the participants' communities, in a private space near their home. Interviews will be digitally recorded and subsequently transcribed verbatim.

Where relevant, these transcripts will be translated from Shona to English. The interview instruments are attached; these questions will be used to guide the discussion, and the interviewer will introduce additional prompts to elicit additional information on key themes based on participant responses. This is a qualitative research best practice and will be most helpful in generative useful, rich data.

*Plan for interpreting results: outcome measures*

The constant comparative method will be used to guide analysis. After each interview is completed, the recording will be transcribed verbatim in Shona then translated into English for analysis. Four researchers of different backgrounds (including researchers from the region where the study is taking place) will separately read four randomly selected transcripts and assign "codes," labels given to each new idea that appears in the transcript. After the four researchers separately code the transcripts, they will come together and compare their code lists, coming to a consensus through discussion about a final code list that captures all of the ideas that they identified in their initial coding. After the development of a single code list, two researchers will separately code each of the transcripts, assigning codes from the code list to each instance of a key idea.

They will then reconcile their assigned codes through discussion. In order to ensure that codes are being assigned consistently, coders will refer back to instances of the same code in prior transcripts (the constant comparative method).

After all transcripts are coded, the research team will come together to identify themes and cross-cutting ideas from the codes, using an iterative analysis process. These themes will be grouped in a modelling process that seeks to understand the care pathways, use of guidelines, acceptability of current management approach; Existing theory will not be used to guide analysis of results, as this may dilute the voices of the participants.

#### Stage 2 - Package development

I. Data collected in stage one (Diagnostic stage) will be compiled and analysis will be done to gain more insight on key areas in the different communities. Data from the qualitative approach will be analyzed through standard methods described above. Quantitative data will be captured and analyzed using SPSS. Knowledge gained from stage one will be used to develop intervention as follows:

i) A report with key findings will be shared with relevant stakeholders (City Health department, DHPOs, Nurse in charge, diabetic association and others involved).

Ii) Four to five manuscript will be submitted based on results from stage 1.

ii) Theory of change workshop (T.o.C.) with relevant stakeholders to build a consensus on the most appropriate intervention for NCDs will be carried out as part of giving feedback to stakeholders of the formative stage. It will include discussions on lessons learnt, barriers and assumptions to finalize the integrated package.

iii) Intervention aimed at addressing the NCDs will be developed and delivered by LHWs the format of this intervention and how it will be evaluated will be determined during stage 2.

#### Stage 3: Pilot intervention

A pilot will be carried out in a few selected clinics in Harare. It is anticipated that pilot intervention package will seek to maximize the interaction and care pathways for all 3 conditions through the provision of an integrated package of health promotion,

Addressing psychological and physical conditions through introducing simple interventions at an early stage, such as diet, weight loss, exercising and addressing kufungisisa.

The final package will seek to provide an integrated pathway by focusing on patient centered care-patient prioritization such as self-care management. Based on the current existing role of the LHWs involved with the Friendship Bench, through FB-Plus they will have a case manager role which will link closely with activities related to the 3 NCDs providing – counseling, adherence support, life style changes and other positive health benefits identified through the study.

#### Stage 4 Evaluation

An appropriate evaluation design for effectiveness of the package will be carried out over a 6 month period. The MRCZ will be informed of this stage of the study through an amendment of this protocol.

*Table 1. Four stages of Friendship Bench Plus*

| **Stage** | **Year 1** | | | | **Year 2** | | | | **Year 3** | | | | **Year 4** | | | |
|---|---|---|---|---|---|---|---|---|---|---|---|---|---|---|---|---|
| | Q1 | Q2 | Q3 | Q4 | Q1 | Q2 | Q3 | Q4 | Q1 | Q2 | Q3 | Q4 | Q1 | Q2 | Q3 | Q4 |
| **Preparatory work** | XX | XX | XX | | | | | | | | | | | | | |
| **Stage 1 Formative work** | | | | XX | XX | XX | XX | XX | | | | | | | | |
| **Stage 2 Develop package** | | | | | | | | XX | XX | | | | | | | |
| **Stage 3 Pilot intervention** | | | | | | | | | XX | XX | | | | | | |
| **Stage 4 Evaluation** | | | | | | | | | | | XX | XX | XX | XX | XX | |
| **Analysis/report writing** | | | | | | | | | | | | | | | XX | XX |

#### *Research Team*

The research team will include the PI, who is the lead researcher for the entire study, project coordinator who will be responsible for planning for all study activities, arrange necessary meetings with relevant stakeholders and also assist in training in relation to the study and research assistants who will help in data collection. The research assistants shall be holders of a first degree in Psychology or a Social Science and must have training on research ethics from a recognized institution. Interns will be students currently studying towards a degree in Psychology from a recognized university and will assist in data collection.

#### *Ethical Considerations*

*Confidentiality*

Following MRCZ protocols all consent forms will be written in such a way that each participant will understand, in a language that they can comprehend, that is English and Shona

Each participant will be provided with two consent forms to sign. They will leave one signed copy with the researcher and the other one they will take home. They will be asked to read and understand the consent, if they cannot read or write, an impartial witness will be called in to assist. All completed consent forms will be kept in a locked room and cupboard with only recognized research team members having access. All analysis will be performed and saved on a password-protected laptop. Any other materials used and with participant details will also be under lock and key.

##### *PID Number*

No names will be included on the data collection tools. Each participant will be assigned a number that will be used on their documents.

##### *Vulnerable populations*

Potential participants who are assessed by study personnel to have a low literacy level will be asked to invite an impartial witness to join in the informed consent process.

##### *Risks*

There are no major risks for the health and wellbeing of the participant that are anticipated.Medical adverse effects (AEs) and possible serious adverse effects (SAEs) are not expected in this study as there are no drugs to be administered. Social AEs and deaths will be reported to the PI and all IRBs within 72 hours of site awareness as per local guidance, regardless of relatedness to study participation.

##### *Benefits*

We cannot and do not guarantee or promise that there are benefits from the study except the acquired knowledge on how to deal with depression, hypertension and diabetes.

##### *Reimbursement*

All participants attending the study will be reimbursed for public transport to the study sites and will be provided with light refreshments at the venue.
