## Appendix 2 ethics approvals for "HeAlth System StrEngThening in four sub_Saharan African countries (ASSET) to achieve high-quality, evidence-informed surgical, maternal and newborn, and primary care: protocol for pre-implementation phase studies"

**Appendix 2: ethics approval numbers from Kings College London and relevant institutional and local government review committees**

| **Country** | **WP** | **Study title** | **KCL approval reference** | **Local ethics committee and associated approval reference number** |
| --- | --- | --- | --- | --- |
| **Ethiopia** | **WP1** | Integrated primary care for mental health, substance use and non-communicable diseases | HR-17/18-7846 | Addis Ababa University-Institution Review Board (Ref 026/18/Psy) |
|  | **WP2** | Integrated maternal health care | HR-17/18-7850 | Addis Ababa University -Institution Review Board (Ref 026/18/Psy) |
|  | **WP3** | Improving access to quality surgical  and dental care | HR-17/18-6144 | Addis Ababa University -Institution Review Board (Ref 026/18/Psy) |
| **South Africa-UCT** | **WP4** | Promoting Person Centred TB Care: A Mixed Methods Study | HR-17/18-7740 | University of Cape Town, Faculty of Health Sciences Human Research Ethics Committee (Ref 286/2018) |
|  | **WP5** | Integrated palliative care and primary health care for chronic obstructive pulmonary disease | HR-17/18-5766 | University of Cape Town, Faculty of Health Sciences Human Research Ethics Committee (Ref 211/2018) |
|  | **WP6** | Reducing the burden of perinatal  common mental disorders (PCMD) and violence against women (VAW) | HR-17/18-7807 | University of Cape Town, Faculty of Health Sciences Human Research Ethics Committee (Ref 286/2018)  Western Cape Government Strategy and Health Support (Ref WC 2-1807_008) |
| **Sierra Leone** | **WP7** | A study of the financial burden on patients undergoing surgical care in Sierra Leone | LRU-17/18-6455 | Sierra Leone Ethics and Scientific Review Committee (Reference number not given - Appendix 2b provides approval letter) |
|  |  | Sierra Leone qualitative study | LRS-17/18-6674 | Sierra Leone Ethics and Scientific Review Committee (Reference number not given) |
|  |  | Sierra Leone quantitative study | LRS-17/18-6219 | Sierra Leone Ethics and Scientific Review Committee (Reference number not given) |
|  |  | Guiding surgical health system strengthening; process mapping of surgical patients in Freetown, Sierra Leone. | LRU-17/18-6537 | Sierra Leone Ethics and Scientific Review Committee (Reference number not given) |
|  |  | Qualitative study-Modification | MOD-19/20-6674 | Sierra Leone Ethics and Scientific Review Committee (Reference number and letter not provided ) |
| **Zimbabwe** | **WP8** | Diagnostic Phase Protocol for Integrated Primary Health Care (Hypertension, Diabetes and Depression) | HR-18/19-10355 | Joint Research Ethics Committee for the University of Zimbabwe, College of Health Sciences and Parirenyatwa Group of Hospitals(Ref 80/19)  Medical Research Council of Zimbabwe (Ref MRCZ/A/2427)  Research Council of Zimbabwe certificate of registration Ref 03553) |
